# Describing variability of intensively collected longitudinal ordinal data with latent spline models

**DOI:** 10.1101/2025.01.14.25320507

**Authors:** Mark Lunt, David A. Selby, William G. Dixon

## Abstract

Population health studies increasingly collect longitudinal, patient-reported symptom data via mobile devices, offering unique insights into experiences outside clinical settings, such as pain, fatigue or mood. However, such data present challenges due to ordinal measurement scales, irregular sampling and temporal autocorrelation.

This paper introduces two novel summary measures for analysing ordinal outcomes: (1) *the mean absolute deviation from the median* (Madm) for cross-sectional analyses and (2) the *mean absolute deviation from expectation* (Made) for longitudinal data. The latter is based on a latent cumulative model with penalized splines, enabling smooth transitions between irregular time points while accounting for the ordinal nature of the data. Unlike black-box machine learning approaches, this method is interpretable, computationally efficient and easy to implement in standard statistical software.

Through simulations, we demonstrate that the proposed measures outperform standard methods when the assumptions of normality or stationarity are violated. Application to real-world data from a national smartphone study, *Cloudy with a Chance of Pain*, highlights the utility of these measures in characterising symptom variability and trends over time.

The methods developed here provide intuitive tools for analysing patient-reported outcomes in longitudinal studies, with potential applications in prediction modelling, causal discovery and evaluation of interventions.

## 1 Introduction

### 1.1 Background

Advances in digital health have enabled the collection of intensive longitudinal data from patient-reported outcomes, particularly through smartphones and wearable devices. These developments have promised to transform population health research for over a decade [1] through ‘mobile health’ or mHealth, which can reach more patients than conventional methods [2], and across different contexts, via ecological momentary assessment [3]. Such data offer unique opportunities to study subjective phenomena such as pain, fatigue and mood outside clinical settings.

Symptoms can be logged daily, as seen successfully in national smartphone studies including Cloudy with a Chance of Pain [4] and the CoViD Zoe Symptom Study [5]. Such insights are invaluable for understanding day-to-day variability, predicting disease flares and assessing the effectiveness of interventions in real-world conditions.

Despite these benefits, analysing such data poses unique challenges. Patient-reported outcomes are often measured on ordinal scales (e.g. ‘none’, ‘mild’, ‘moderate’, ‘severe’) and are collected at irregular intervals, introducing complexity in handling variability, trends and temporal dependencies. Traditional methods, designed for continuous, regularly sampled, complete data, often fail to respect the ordinal nature of measurements or capture non-linear trends over time.

### 1.2 Challenges in analysing longitudinal ordinal data

Analyses of patient-reported outcomes can address descriptive, ætiological and predictive questions. Descriptive analysis includes quantifying how symptoms fluctuate and identifying ‘typical’ levels and the variability around them, which may not be common across patients. The prediction of time-varying patterns has the potential to identify moments of interest, such as a disease flare, in order to enable a just-in-time adaptive intervention [6]. Ætiological questions need to correlate more than one stream of data to another, allowing us to understand how a time-varying exposure can influence a time-varying outcome. However, limited guidance is available on how best to process longitudinal, subjective symptom ratings.

Recording patient symptom data outside the clinical setting throws up several important challenges:(i) subjectivity in reporting symptom severity; (ii) irregular measurement timing (even with dedicated participants and automated reminders) [7], [8]; (iii) temporal autocorrelation, with symptoms exhibiting smooth, gradual changes of time, poorly modelled by methods assuming independence between observations; and (iv) non-alignment in time, wherein patients describe their day-to-day experience, not necessarily following a common intervention.

To address these challenges, robust statistical methods are needed that respect the ordinal structure of the data, account for temporal autocorrelation and irregular sampling, and provide interpretable results that are accessible both to epidemiologists and clinicians.

The challenges described above can be addressed using generalized linear mixed models (GLMMs) for ordinal data [see e.g. 9], which can account for irregular measurement timing and subject-specific variation through the inclusion of random effects. However, GLMMs typically require the analyst to prespecify the functional form of time’s effect on the outcome (e.g. as a linear or polynomial term). This assumption can be difficult to justify for complex symptom data, where trajectories can be highly non-linear. The penalized spline approach we adopt is a special case of a generalized additive mixed model, offering a more automated, data-driven method for modelling these complex trends without strong *a priori* assumptions. Furthermore, our focus is not just on modelling but on deriving simple, interpretable summary statistics to quantify variability around flexible, subject-specific trajectories. This provides a clinically intuitive tool to complement existing models.

### 1.3 Contributions of this paper

Though it may sometimes be reasonable to treat ordinal data as continuous (e.g. if the number of levels is large) [10], it is often unjustified, leading to reduced statistical power and erroneous conclusions [10], [11]. In this paper, we propose summary measures of variation for ordinal outcomes with a small number of measurement levels. The first, the *mean absolute deviation from the median* (Madm), is an ordinal analogue to standard deviation, applicable for cross-sectional analyses or when the average (median) reported symptom scores do not appear with vary with time.

Using a latent cumulative model allows for the possibility of ordinal levels that are unequally distributed in the latent space. For dynamic longitudinal data, a penalized spline approach enables smooth transitions between irregular time points, with minimal input from the analyst on choice of hyperparameters or arbitrary thresholds, and while remaining more interpretable than a black box machine learning framework. Moreover, the latent spline model can be easily fit in existing standard statistical software with minimal lines of code, and is readily visualized on either the latent symptom or observed measurement scales. A derived summary measure the *mean absolute deviation from expectation* (Made) is proposed for intensive longitudinal ordinal data where symptom reports appear to vary over time and where the mean scores are estimated using such a latent spline model.

In an empirical analysis, the derived indices will be applied to eight simulated individuals, chosen to illustrate different aspects of the indices. This is followed by a real-world application to data from a national smartphone study of people living with chronic pain: *Cloudy with a Chance of Pain* [4].

## 2 Methods

### 2.1 Motivating example

Consider eight simulated^1^ example subjects, who each provide up to 100 consecutive daily reports of their self-assessed pain severity on a five-level ordinal scale: 1 (no pain), 2 (mild pain), 3 (moderate pain), 4 (severe pain) and 5 (very severe pain), shown in Figure 1. Subjects a, b and c all have a constant median pain level, with different levels of variability in the observations. Subjects d and e have pain that is, respectively, increasing or decreasing over time. The remaining three subjects experience sinusoidal patterns with different amplitudes and frequencies.

**Figure 1:**
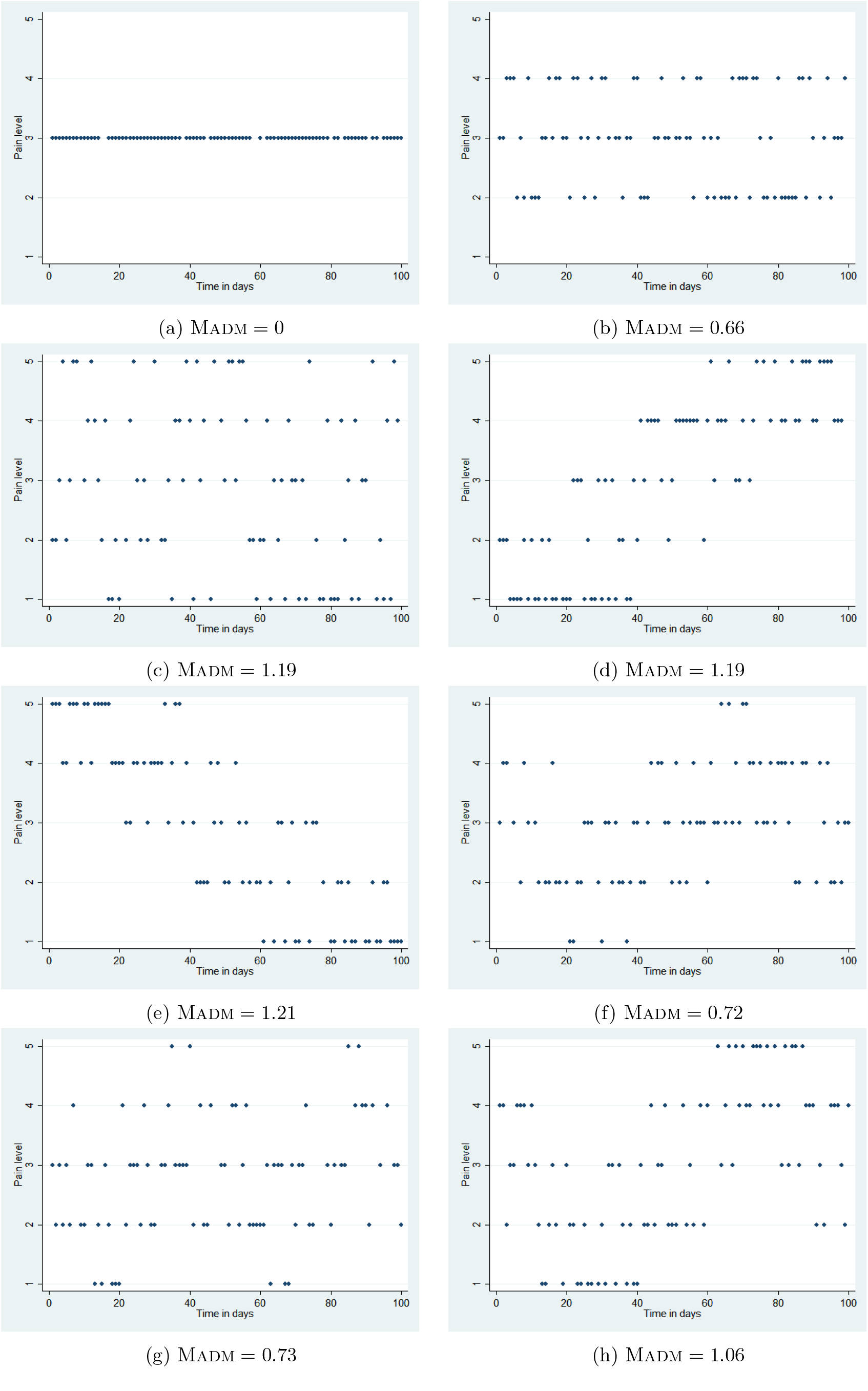
Reported pain severity over time for eight simulated example patients

If marginalising over time, any summary of the distribution of such data is based on estimating the ‘average’ measurement and the variability around it. Frequently, intra-individual pain variability is measured using the intra-individual standard deviation (ISD) [12]. However, estimating a standard deviation also requires estimating a sample mean, which is defined for continuous interval measurements but not for ordered categorical data: the pain severity level ‘2’ is surely greater than ‘1’ and less than ‘3’ but not necessarily halfway between them. Alternatives to the mean include the mode and the median, with the latter possibly a more robust useful measure of central tendency (see Appendix B).

This motivates the derivation of an index similar to ISD, but based on the median rather than the mean scores. The useful mathematical properties of mean squared differences from the mean (i.e.variance) do not hold for mean squared differences from the median, however. Hence we suggest a simpler and more interpretable index, the *mean absolute deviation from the median* (Madm), defined

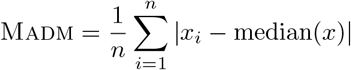

where *x*_*i*_ is the pain level reported by a given subject on the *i*^th^ measurement occasion, *i* = 1, …, *n*. The Madm may be interpreted as the mean ‘distance’ (number of ordinal levels) from the median value. The larger the Madm, the more variable a subject’s pain reports. Figure 1 gives the Madms for each of the eight simulated subjects, all of whom have a median pain score of 3. Using the median absolute deviation, rather than the mean, would offer far less discrimination, since it would need to take the value of one of the pain levels for each subject, so in our example could only take 5 different values in any sample.

However, neither the Madm nor the ISD can describe or offer evidence of systematic variation over time, even if trends were readily apparent from a simple graphical summary. Moreover, a sequence of ordered categorical values may not be satisfactorily imputed with an overall sample median or mode, as it may introduce sharp discontinuities in the temporal trajectory. Hence, we build a statistical model to estimate the level of pain (or other reported symptoms) over time, based on patient reports.

The methods proposed here involve assuming a smooth underlying trajectory of pain (equivalently: fatigue, mood or other symptoms), based on the assumption that pain is a latent phenomenon in continuous space that varies between discrete measurement times, and that discrete ordinal levels are a property of the measurement device rather than the biological process.

### 2.2 Latent ordinal spline models

A simple ordinal logit or probit regression model of pain against time, while easy to fit, would assume a linear trend with time: an heroic assumption that (as we will see) is unlikely to hold for real data from patients with musculoskeletal conditions. Using a spline basis for time allows for an arbitrary amount of variation in the predicted pain level over time, though typically requires an analyst to decide how much variation would be allowed by choosing how many segments (often referred to as control points, knots or bins) in which to to divide the follow-up time: more bins allows for more variability at the expense of model complexity.

One proposed method to make this process more robust is *penalized* splines or *P*-splines [13], [14]. This approach divides the follow-up time into a very large number of bins, more than should be required to model the changes in reported pain over time. Then a penalty term is calculated from the magnitudes of the changes of the coefficients from one bin to the next, and this penalised likelihood is maximised to estimate the coefficients, effectively determining the number of knots (and implicitly the amount of variation) automatically from the data.

Fitting a simple spline model will produce a different coefficient for each bin, giving the model the same number of degrees of freedom as the number of bins. With a penalised spline model, coefficients for adjacent bins are constrained to be close to each other, reducing the number of degrees of freedom in the model. The model will report an “effective degrees of freedom”: the larger this number, the wigglier the association between pain and time and the more complex the spline model [15]. An effective degrees of freedom of 1 corresponds to a straight line, or a linear association with time.

Coefficients of a spline basis may be difficult for clinicians to interpret, but a graphical representation can illustrate the model clearly. As well as a smooth curve in the latent space, the output of an ordered categorical model includes a set of estimated probabilities, one for each ordinal pain level, representing the probability of reporting pain at that level on that day. These may be visualised as a heatmap, where the shading of tiles corresponds to the probability of reporting each pain level at each time point.

This set of probabilistic predictions may be complemented by a single “expected” pain level at each time point. Conventionally, we could report the pain level with the highest probability, i.e. the mode, as the expected level. Alternatively, we could use the “median” pain level based on the cumulative probabilities: the first level such that the probability of being at that level or lower is at least 50%.

To compare our latent ordinal spline model with the implied baseline (a constant median model), we derive an alternative to the Madm, the *mean absolute deviation from the expectation* (Made), which measures observed deviations from a dynamic model making variable predictions over time,

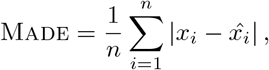

where 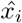 is the median pain severity level predicted by a (spline) model at time point *i*.

The interpretation of the Made is the same as for the Madm: how many steps from the expected value is the observed value on average. If the Made is not much less than the Madm, the model has not improved prediction much: observed values are no closer to the predicted value than they are to the median. The Made can even be larger than the Madm for a poor model. However, the relative reduction in absolute deviation 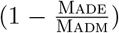 may make a reasonable statistic for the explanatory power of a model.

The results of fitting penalised spline ordinal regression models to the 8 example subjects are shown in Table 1 and Figure 2. It is not possible to fit a regression model to subject 1, whose reported pain was the same for every time point.

**Table 1:**
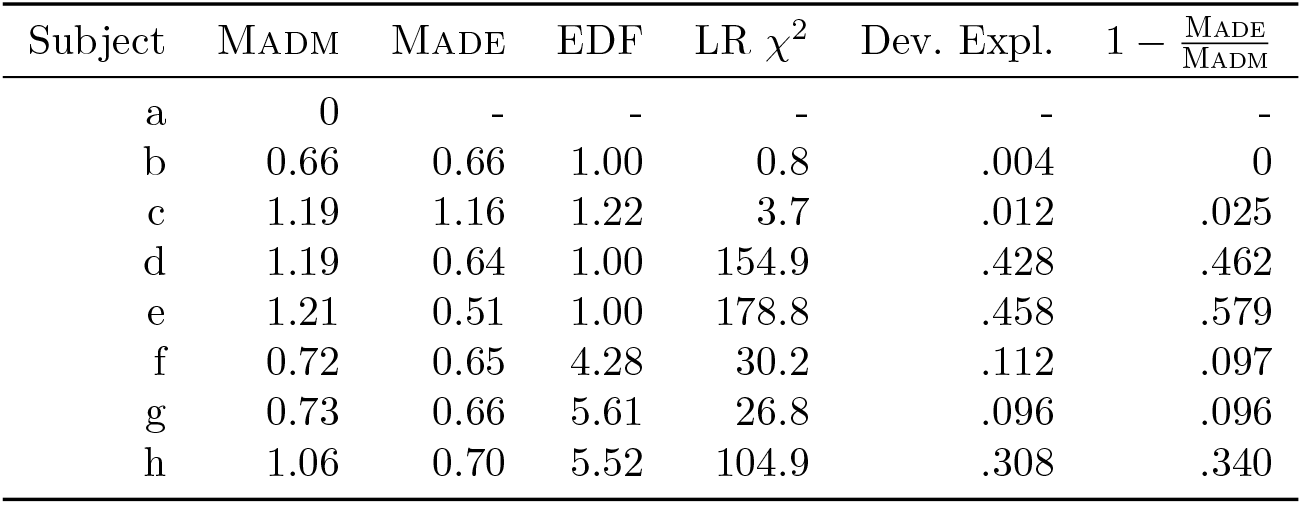
Summaries of ordinal regression models fitted to simulated subjects.

**Figure 2:**
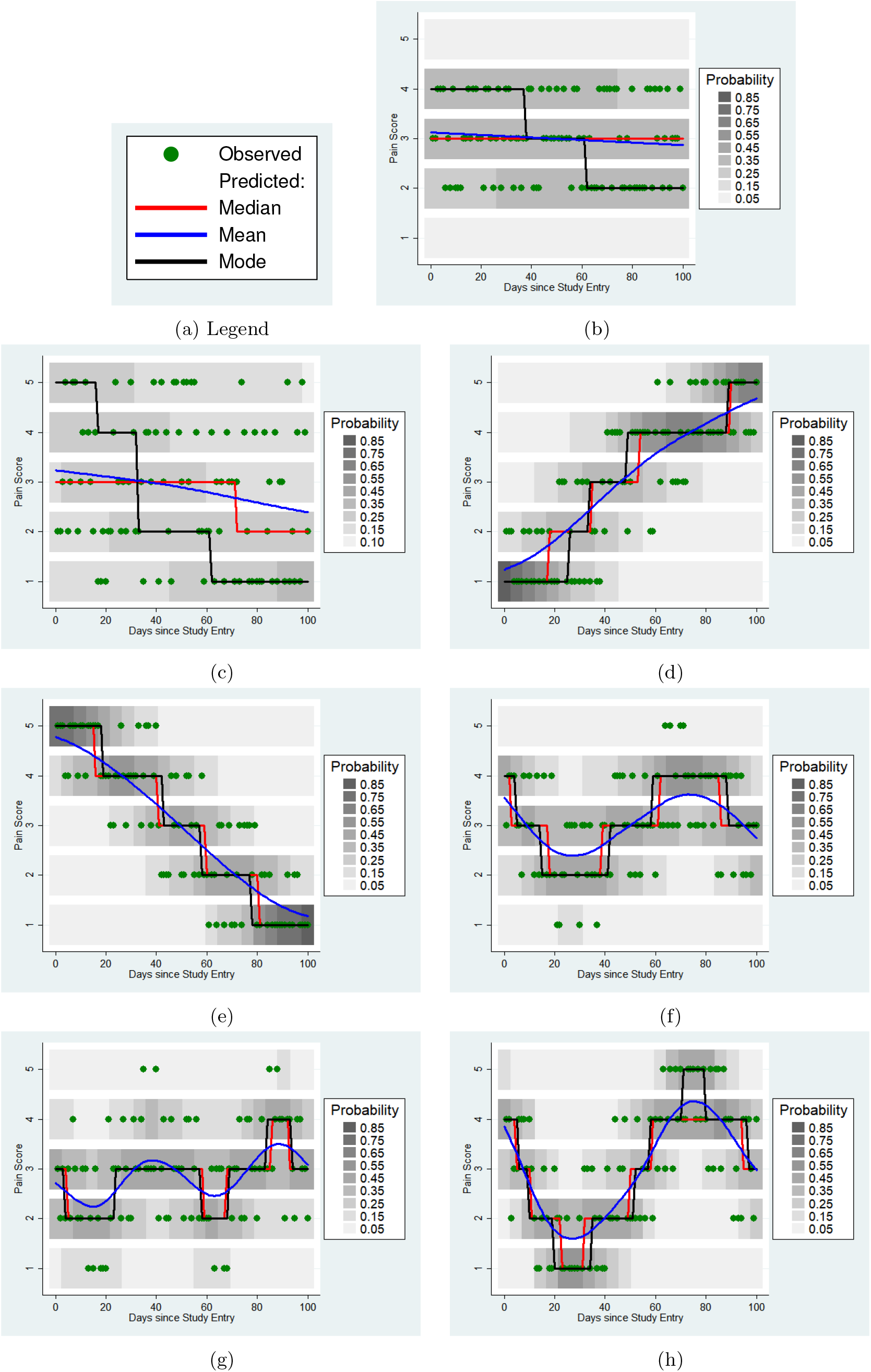
Results of ordinal regression models in example subjects. Blue curves represent the smooth function in the latent space; the grey scale shows the estimated probability of each of the five ordinal levels at each time point, with red lines indicating the most probable such level

As expected, a time-varying model offers little improvement for subjects (b and c) who exhibit no systematic changes in symptoms over time: the Made is not noticeably smaller than the Madm, and this is further quantified with the *χ*^2^ likelihood ratio test statistic. Subjects d and e show a steady change in pain over time, upwards and downwards respectively. This is clear in Figure 2, but can also be read from Table 1. The likelihood ratio statistic is highly significant, but the effective degrees of freedom are 1 in both cases, suggesting a linear trend. Subjects f, g and h all show non-linear changes in pain levels over time, for which simple linear models would offer a poor fit.

The median, Madm, latent ordinal spline model and the derived summary statistics now provide a strategy for describing and grouping individuals according to how their pain changes with time. Statistically non-significant coefficients in the penalised spline model suggest symptoms that do not vary with time, whilst effective degrees of freedom less than 2 indicate a linear trend, and effective degrees of freedom more than 2 imply non-linear fluctuations in reported symptoms over time, which can be visualised with heat maps or predicted curves.

## 3 Results

We illustrate this approach to investigating intraindividual variability in pain by applying it to data taken from the *Cloudy with a Chance of Pain* study [16]. A total of 10,430 subjects contributed to this study, but many were ‘tourists’ who only provided a few days of data [16]. This analysis was restricted to the 2,197 subjects who contributed at least 60 days of data, and reported at least two different pain levels within that data. These subjects contributed between 60 and 449 observations each, with 75% contributing at least 89, 50% contributing at least 235 and 25% contributing at least 196.

Subjects were asked to report a number of potentially painful conditions; in this analysis we are concerned with four: rheumatoid arthritis, osteoarthritis, ‘other arthritis’ and chronic widespread pain. Subjects were divided into six groups according to the self-reported presence or absence of these conditions, as follows:

1. rheumatoid arthritis and no other condition
2. osteoarthritis and no other condition
3. ‘other arthritis’ and no other condition
4. chronic widespread pain and no other condition
5. chronic widespread pain plus at least one other condition
6. none of the four conditions

The numbers of subjects with each condition are given in Table 2. A total of 323 subjects did not fit into any of these groups, i.e. reported two or more types of arthritis, but all groups with particular combinations of types were small and excluded from the analysis.

**Table 2:**
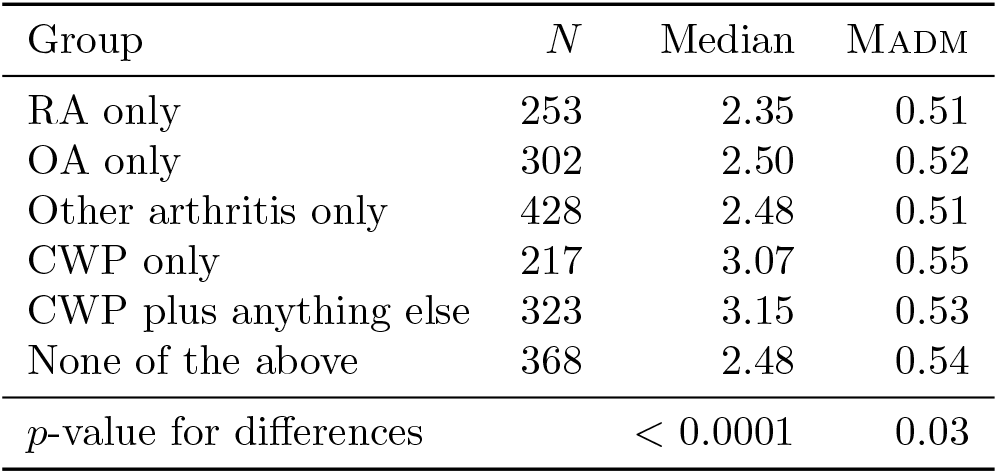
Time-insensitive parameters for subjects with different conditions in the Cloudy data, with results of Kruskall–Wallis test for differences between groups.

The median pain reported was higher for the two ‘chronic widespread pain’ groups than the arthritis groups or the no condition group (see Table 2). The Madm was lower in the arthritis groups than the other conditions, but the between-group differences were not nearly as statistically significant as the between group differences in median.

Overall, 20% of subjects showed no significant changes of pain over time, but this varied significantly between the condition groups (*p <* 0.001), being lower in the RA and other arthritis groups (see Table 3. There was no difference between the condition groups in the proportion of subjects showing a linear trend in pain, which was markedly less (7.5%).

**Table 3:** Percentage of subjects with each of the three types of variation of pain with time (none, linear, non-linear) for subjects with different conditions in the Cloudy data.

| Group | Time Effect |  |  |
| --- | --- | --- | --- |
|  | None | Linear | Non-linear |
| No Condition Reported | 24 | 7 | 69 |
| RA Only | 12 | 9 | 79 |
| OA Only | 25 | 8 | 67 |
| Other Arthritis Only | 16 | 7 | 77 |
| CWP Only | 25 | 7 | 68 |
| CWP plus anything else | 21 | 7 | 72 |

Penalized spline models were fitted using the function gam() from R package mgcv [17]; with a *P* spline term and ordered categorical response. By default, the number of knots is *k* = 10, with positions equally spaced across an interval 0.1% wider than the range of the data.

For all of the condition groups, the Made was markedly lower than the Madm, suggesting that the penalised spline models capture some of the variability in pain (see Table 4). However, the amount of variability that could be explained by the models varied between the conditions, with more being explained in RA only and other arthritis. This is not entirely due the greater proportion of subjects in these groups showing significantly non-linear trajectories, since the deviance explained in these groups is still higher when restricting attention to those with non-linear trajectories (see Table 5).

**Table 4:** Time sensitive parameters for subjects with different conditions in the Cloudy data.

| Condition | MADM | MADE | Dev. Expl. | $1 - \frac{\text{MADE}}{\text{MADM}}$ |
| --- | --- | --- | --- | --- |
| No Condition Reported | 0.54 | 0.46 | 0.12 | 0.14 |
| RA Only | 0.51 | 0.41 | 0.16 | 0.18 |
| OA Only | 0.52 | 0.45 | 0.10 | 0.12 |
| Other Arthritis Only | 0.51 | 0.42 | 0.14 | 0.16 |
| CWP Only | 0.55 | 0.48 | 0.10 | 0.11 |
| CWP plus anything else | 0.53 | 0.45 | 0.11 | 0.12 |

**Table 5:**
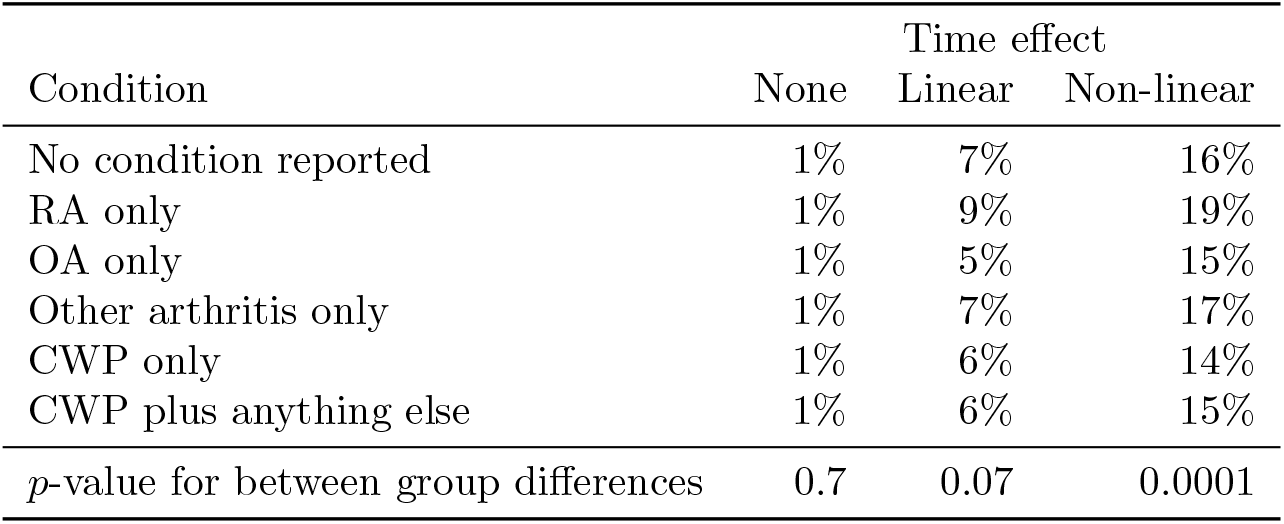
Variation in deviance explained by model type and reported condition in the Cloudy data.

The relative reduction in mean absolute deviation correlated well with the deviance explained (Spearman correlation coefficient = 0.75, *p <* 0.00001). The values are also numerically similar, as shown in Table 6.

**Table 6:** Variation in relative reduction in mean absolute deviation by model type and reported condition in the Cloudy data.

| Condition | Time-Effect |  |  |
| --- | --- | --- | --- |
|  | None | Linear | Non-linear |
| RA only | 1% | 9% | 21% |
| OA only | 1% | 4% | 17% |
| Other arthritis only | 1% | 9% | 20% |
| CWP only | 1% | 7% | 16% |
| CWP plus anything else | 2% | 6% | 16% |
| None of the above | 1% | 9% | 19% |
| *p*-value for between group differences | 0.7 | 0.07 | 0.0001 |

For subjects showing no significant pattern of changes in pain over time, the only graphic needed is to show the proportion of time spent at each pain level. Figure 3 show stacked bar charts for 9 typical subjects from the Cloudy data. The left hand three all have a median pain level of 1, the middle three have a median pain level of 3 and the right hand three have a median pain level of 5. In each group of 3 subjects, the Madm increases from left to right.

**Figure 3:**
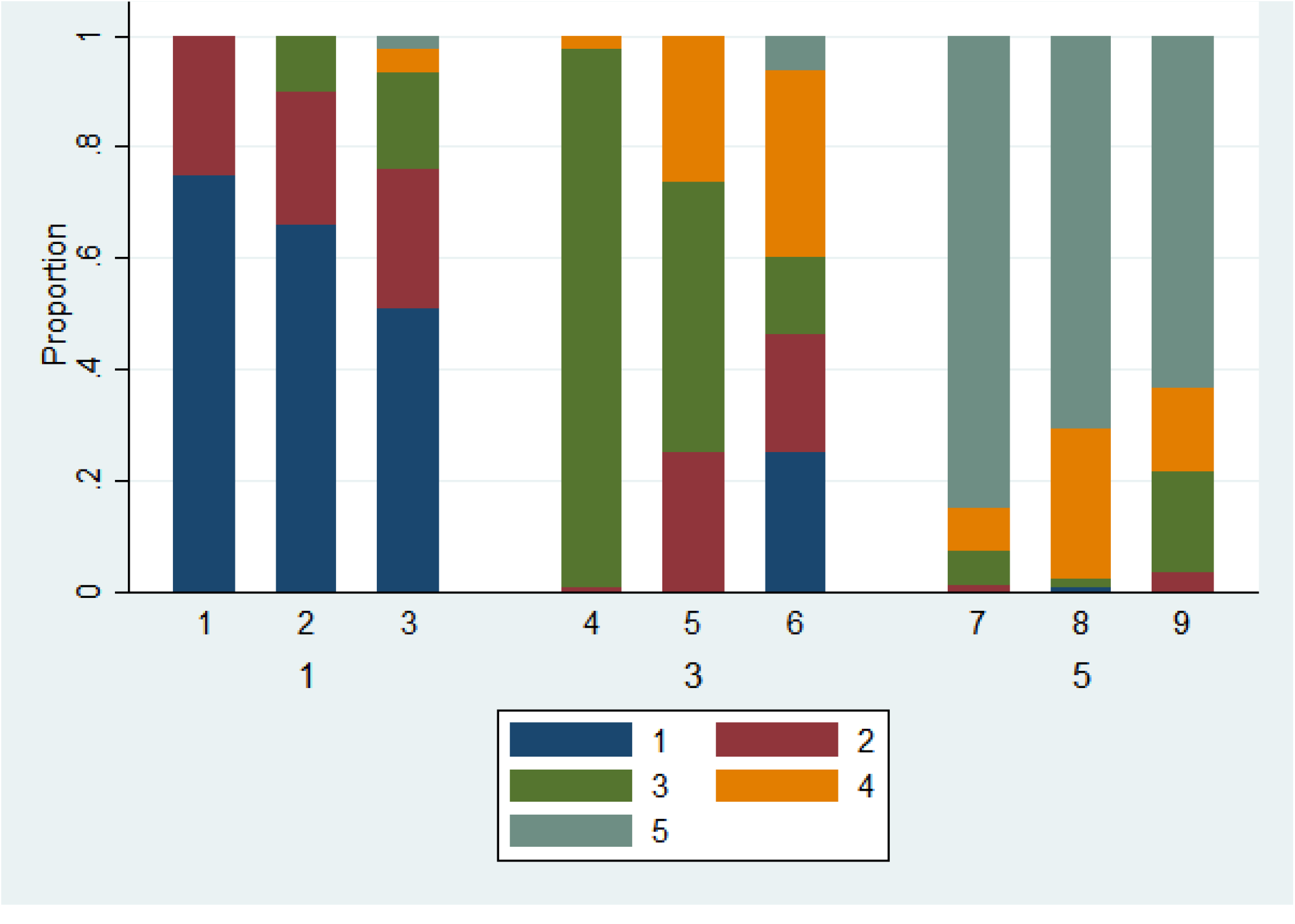
Stacked bar charts for varying median and Madm

Figure 4 shows heatmaps for three subjects from the Cloudy data with significant changes in pain over time, but no evidence of non-linearity and different amounts of deviance explained. The first subject had an expected pain level of 3 throughout the follow-up except at the end. The latent ordinal spline model explained about 1% of the reported variation in pain. The second subject had an expected pain of 3 for the first half of their follow-up, and 2 for the second half: this model explained about 5% of the variation in pain. The third had an expected pain of 2 at the beginning of their follow-up, but soon settled to a consistent expected (and observed) pain level of 1: this model explained 38% of the variation in pain.

**Figure 4:**
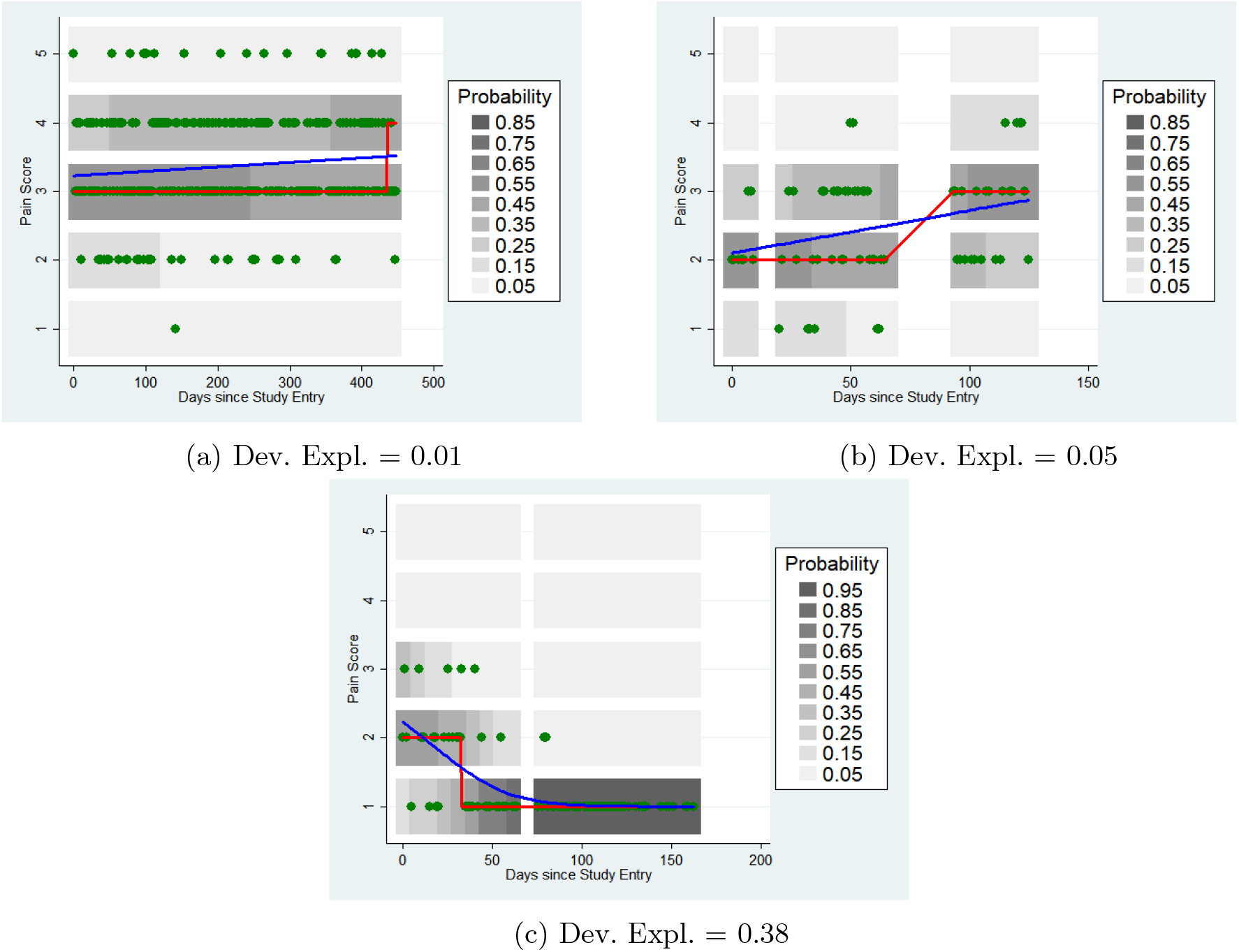
Ordinal Regression Models: Linear Trend. Blue curve is the fitted latent spline; red is the most likely pain level

If the model suggests a non-linear pain trajectory, larger differences between the heatmaps are possible. Figure 5 shows heatmaps for 9 randomly selected Cloudy subjects. The left hand column contains subjects selected from the lowest tertile of deviance explained, the middle column contains subjects from the middle tertile, and the right column contains subjects from the highest tertile. Similarly, the top row contains subjects from the lowest tertile of EDF (simplest trajectories), the middle row contains subjects from the middle tertile and the bottom row subjects from the top tertile (most complex trajectories).

**Figure 5:**
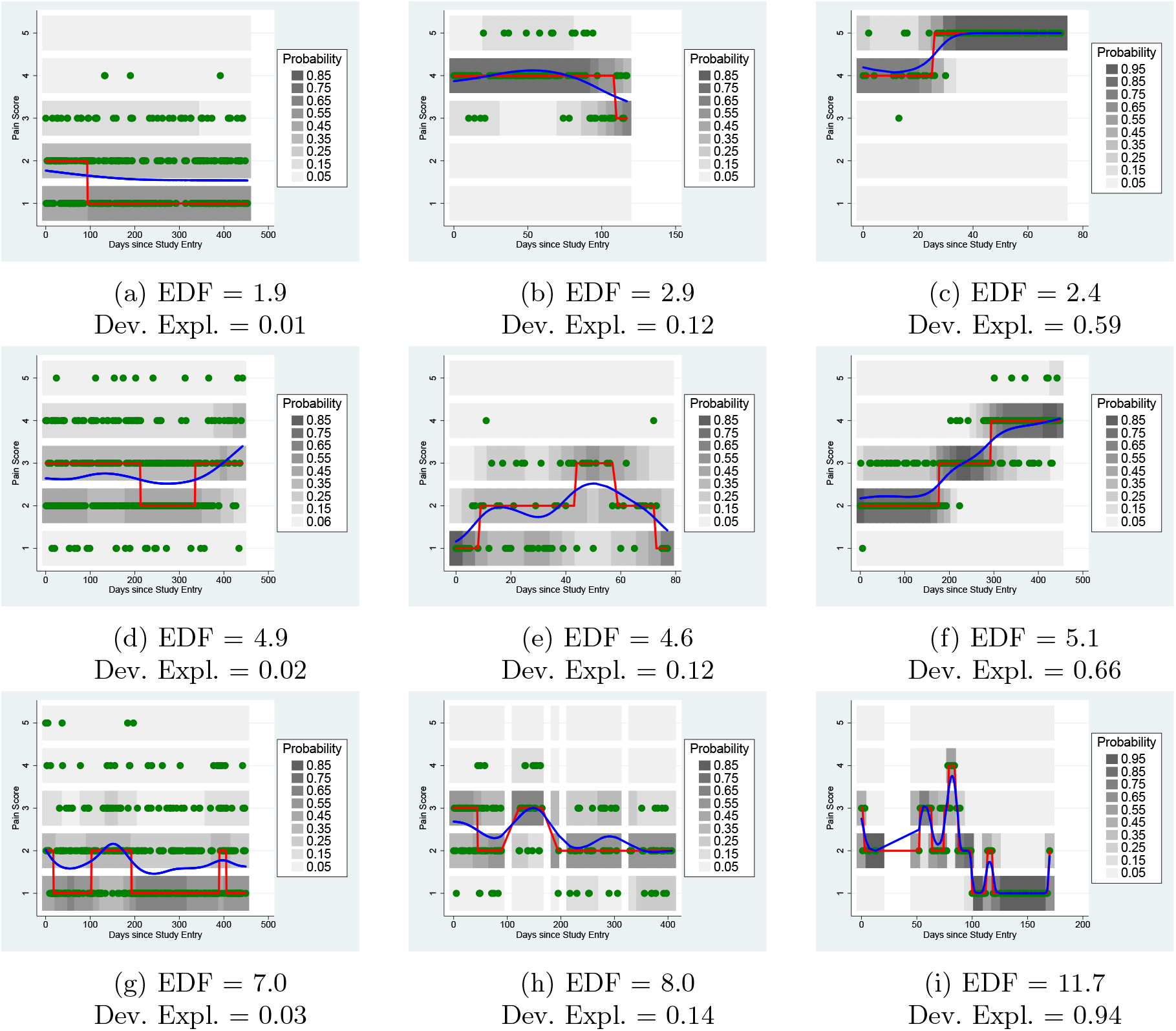
Ordinal Regression Models: Non-linear trend. Blue curve is the fitted latent spline; red is the most likely pain level

## 4 Discussion

### 4.1 Summary

In this paper, we have highlighted the challenges associated with summarising ordinal, irregular time series data of the kind typically collected in longitudinal mobile health studies. Such measurements are subjective, not necessarily comparable between patients, replete with missing values or irregular intervals and likely to exhibit temporal patterns. We have proposed alternative graphical and numerical summaries describe the subject’s experience, depending on whether the typical pain level varies over time for that subject or not. The proposed solution for those subjects whose pain does change over time can be fitted in standard statistical software and require minimal data preprocessing, imputation or hyperparameter tuning. Unlike many methods commonly used for such data, the methods we propose respect the ordinal nature of the data. We have shown that the proposed parameters vary between groups of subjects with different conditions in the way that they might have been expected to, providing some initial validity.

The latent penalised spline model allows for systematic changes in typical pain level over time. Not only does this make it possible to visualise smooth changes underlying discrete pain measurements, but it also reduces the amount of unexplained variation in the data. This increases the power with which it is possible to identify predictors of pain, since current pain levels are compared to the time-varying predictions for pain, rather than the overall mean or median for that subject.

Having an expected pain level that varies over time also improves statistical power to detect times at which pain is either higher or lower than expected. Conventionally, to detect a pain flare, the current level of pain is compared to the previous pain level: if the change is an increase greater than a prespecified threshold, a flare is said to have occurred. However, both measurements are made with error, so the error in the change will be greater than the error in either measurement separately. The error in the differences between the current measurement and the current expected measurement will be smaller, due to the smoothing applied to the expected measurements.

### 4.2 Limitations

There are, however, some additional challenges that the methods proposed do not overcome. Firstly, there is the subjective nature of pain. What one patient reports as a 3, another (or the same patient the following year) may consider a 2 or a 4, based on their individual experiences. The ordinal regression model used allows for a different scale for each subject, since it models the value that that individual would report, through random effects or simply fitting a separate model to every participant. Hence comparisons within a subject may be valid, but comparisons between subjects are less reliable. Modelling the effect of a predictor involves estimating its average effect on all of the different scales within the sample. However, this is true of all ordinal regression models.

The Madm may be criticised as assuming an interval scale, since it counts how many steps apart two measurements are. It may be that going from level 4 to level 5 is a greater increase in pain than going from level 1 to level 3, despite being fewer steps. In addition, a given change in pain level may be treated as a different number of steps by different individuals. Nonetheless, our approach is far less restrictive than treating pain as an interval measurement, which assumes not only that pain levels are the same distance apart for each subject, but also that each pain level has the same meaning for each subject. The interpretation of the Madm and Made as the ‘mean number of levels’ of deviation relies on the numerical encoding of the ordinal categories as consecutive integers (e.g. 1, 2, 3, 4, 5). While the measures are mathematically robust to a linear shift in this coding, for any other numerical labels, an implicit recoding to consecutive integers is required. The assumption about counting steps between ranks remains far less restricting than treating the ordinal outcome as a true interval-scale variable.

### 4.3 Conclusion

The proposed methods have been selected to be intuitive and interpretable such that they can be used by patients and clinicians in discussing a patient’s pain (or other symptomatic) experience. These tools should also provide an extensible framework for identifying predictors of ordinal outcomes.

A statistically valid procedure for summarising and describing variation in patient-reported symptom data, as described here, may be further extended for ætiological questions, such as the effects of interventions or time-varying lifestyle factors on day-to-day experiences of long-term conditions. Future work may extend these tools to improve the design of mobile health studies and adaptive interventions.

## Supporting information

R code for simulations + LaTeX source files

## Data Availability

Data produced in the present study are available upon reasonable request.

## Statements and declarations

### Author contributions

The first draft was written by David Selby and subsequently revised by Mark Lunt with comments from William Dixon. The application of latent spline models is attributable to David Selby, the Madm and Made summary statistics to Mark Lunt; both contributed to coding, analysis and visualizations. All authors read and approved the final manuscript.

### Data availability statement

Simulated data may be reproduced using the supplementary R code. The data from the Cloudy with a Chance of Pain study is available from the corresponding author on reasonable request.

### Ethics approval

Analyses involve either simulated data, or observational data already collected from the ‘Cloudy with a Chance of Pain’ study; for further information on that study, see [4].

## A Simulation of Example Subjects

Observations were simulated based on “states”, with each state having a fixed probablility of producing each possible pain level listed in Table 7. Each individual’s simulated data consists of 100 observations in 10 groups of 10. The states for each subject at each time point are listed in Table 8 Since the ability to handle missing data is important, having generated 100 observations, each observation was set to missing with a probability of 0.1.

**Table 7:** Probability of reporting each pain level in each state.

| State | Probability of being in pain state: |  |  |  |  |
| --- | --- | --- | --- | --- | --- |
|  | 1 | 2 | 3 | 4 | 5 |
| A | 0.00 | 0.00 | 1.00 | 0.00 | 0.00 |
| B | 0.00 | 0.33 | 0.34 | 0.33 | 0.00 |
| C | 0.20 | 0.20 | 0.20 | 0.20 | 0.20 |
| D | 0.50 | 0.50 | 0.00 | 0.00 | 0.00 |
| E | 0.33 | 0.34 | 0.33 | 0.00 | 0.00 |
| F | 0.00 | 0.33 | 0.34 | 0.33 | 0.00 |
| G | 0.00 | 0.00 | 0.33 | 0.34 | 0.33 |
| H | 0.00 | 0.00 | 0.00 | 0.50 | 0.50 |

**Table 8:** Pain states for each individual at each time period.

| Subject | Time Interval |  |  |  |  |  |  |  |  |  |
| --- | --- | --- | --- | --- | --- | --- | --- | --- | --- | --- |
|  | 1 | 2 | 3 | 4 | 5 | 6 | 7 | 8 | 9 | 10 |
| 1 | A | A | A | A | A | A | A | A | A | A |
| 2 | B | B | B | B | B | B | B | B | B | B |
| 3 | C | C | C | C | C | C | C | C | C | C |
| 4 | D | D | E | E | F | F | G | G | H | H |
| 5 | H | H | G | G | F | F | E | E | D | D |
| 6 | F | F | E | E | F | F | G | G | F | F |
| 7 | F | E | F | G | F | F | E | F | G | F |
| 8 | F | E | D | E | F | F | G | H | G | F |

## B Robustness of Median compared to Mode

Compare the two graphs in Figure 6, each showing 100 observations on a 5-level ordinal scale. The median in both cases is 3, but the mode is 1 in the first case and 5 in the second, despite the fact that only one observation differs between them.

**Figure 6:**
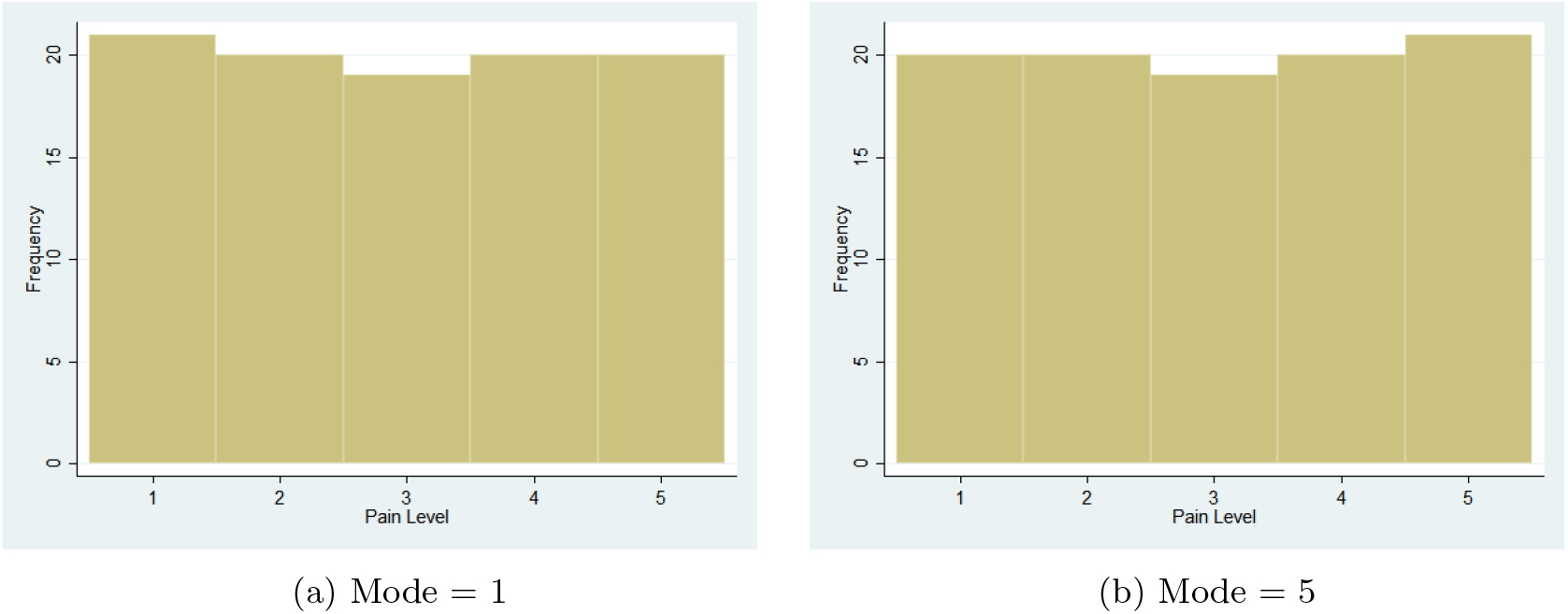
Histograms of pain levels

## C Estimating Median and Madm from Stacked Bar Chart

Alternatively, we can show graphically the proportion of time spent at each pain level via a stacked bar chart, such as Figure 7, and use this to estimate the Madm for each subject. Clearly, there is no variation for subject 1, and the Madm is 0. Subject 2 spends approximately equal times at pain levels 2, 3, and 4. The median pain level is 3, and pain levels 2 and 4 are one step away from the median. So the Madm for this subject is approximately 1*/*3 *×* 1 + 1*/*3 *×* 0 + 1*/*3 *×* 1 = 0.67. Subjects 3, 4 and 5 all appear to spend approximately equal amounts of time at each pain level, to give a Madm of 2*/*5 *×* 2 + 2*/*5 *×* 1 + 1*/*5 *×* 0 = 1.2. Subjects 6 and 7 spend a little time at pain levels 1 and 5, so their Madms are roughly 1*/*10 *×* 2 + 6*/*10 *×* 1 + 3*/*10 *×* 0 = 0.8

**Figure 7:**
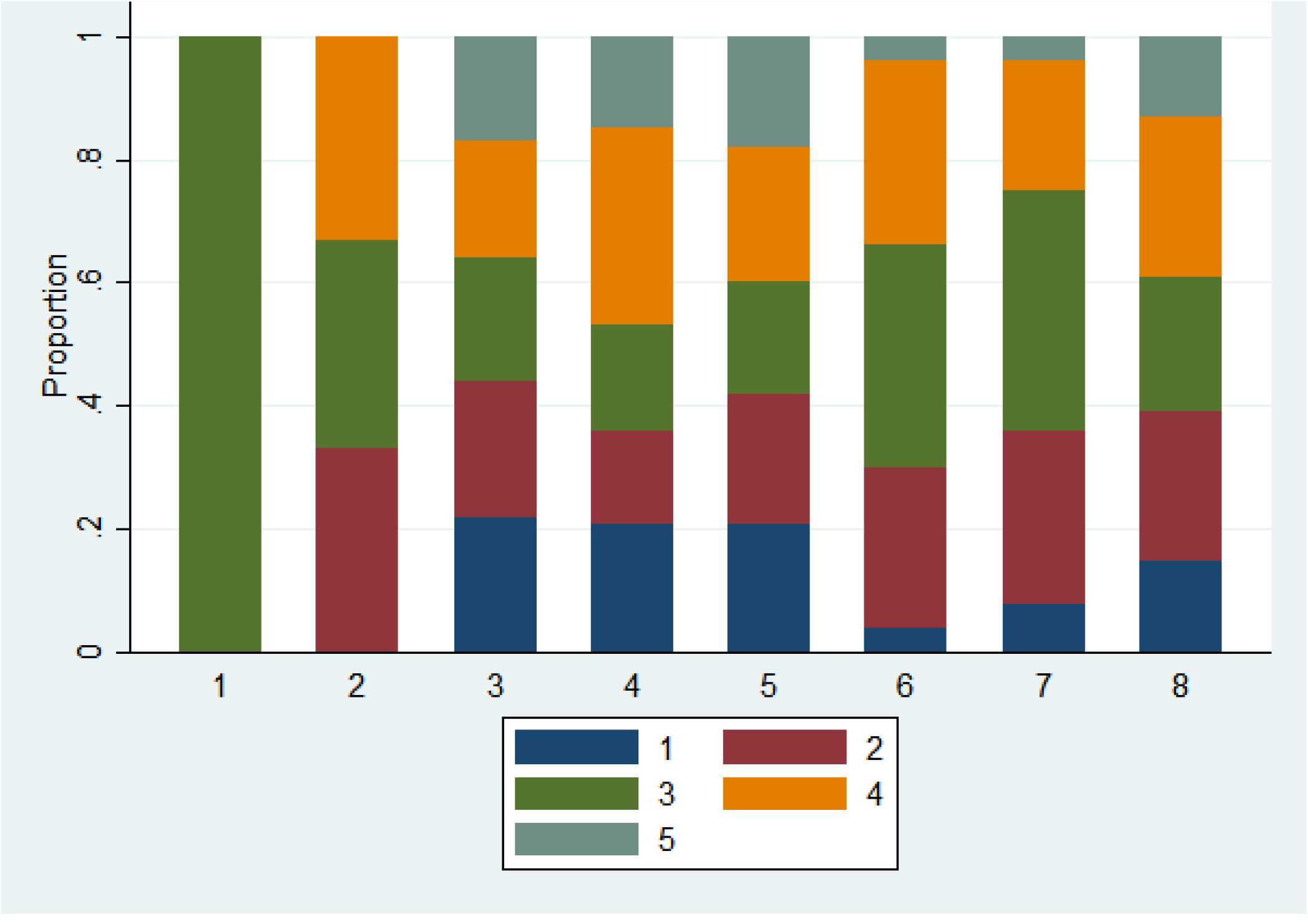
Stacked bar charts for the pain measurements

## Footnotes

1 Further simulation details are given in Appendix A

## References

[1] E. C. Hayden, ‘Mobile-phone health apps deliver data bounty,’ Nature, vol. 531, 7595 2016, issn: 14764687. doi: 10.1038/531422a.

[2] M. S. Marcolino, J. A. Q. Oliveira, M. D’Agostino, A. L. Ribeiro, M. B. M. Alkmim and D. Novillo-Ortiz, ‘The impact of mHealth interventions: Systematic review of systematic reviews,’ JMIR mHealth and uHealth, vol. 6, no. 1, e23, Jan. 2018. [Online]. Available: 10.2196/mhealth.8873.

[3] C. N. Scollon, C.-K. Prieto and E. Diener, ‘Experience Sampling: Promises and Pitfalls, Strength and Weaknesses,’ in Assessing Well-Being, Springer Netherlands, 2009, pp. 157–180. doi: 10.1007/978-90-481-2354-4_8.

[4] W. G. Dixon, A. L. Beukenhorst, B. B. Yimer et al., ‘How the weather affects the pain of citizen scientists using a smartphone app,’ npj Digital Medicine, vol. 2, 1 2019, issn: 23986352. doi: 10.1038/s41746-019-0180-3.

[5] C. Menni, A. M. Valdes, M. B. Freidin et al., ‘Real-time tracking of self-reported symptoms to predict potential covid-19,’ Nature Medicine, vol. 26, 7 2020, issn: 1546170X. doi: 10.1038/s41591-020-0916-2.

[6] P. Klasnja, E. B. Hekler, S. Shiffman et al., ‘Microrandomized trials: An experimental design for developing just-in-time adaptive interventions,’ en, Health Psychol., vol. 34S, no. Suppl, pp. 1220– 1228, Dec. 2015.

[7] P. Hall, H.-G. Müller and F. Yao, ‘Modelling sparse generalized longitudinal observations with latent Gaussian processes,’ Journal of the Royal Statistical Society: Series B (Statistical Methodology), vol. 70, no. 4, pp. 703–723, Sep. 2008. doi: 10.1111/j.1467-9868.2008.00656.x.

[8] K. L. Druce, W. G. Dixon and J. McBeth, ‘Maximizing engagement in mobile health studies,’ Rheumatic Disease Clinics of North America, vol. 45, no. 2, pp. 159–172, Ma. 2019. doi: 10.1016/j.rdc.2019.01.004.

[9] W. W. Stroup, M. Ptukhina and J. Garai, Generalized Linear Mixed Models: Modern Concepts, Methods and Applications, Second. Chapman and Hall/CRC, Mar. 2024, isbn. 9780.29092060. doi: 10.1201/9780429092060. [Online]. Available: http://dx.doi.org/10.1201/9780429092060.

[10] G. Z. Heller, M. Manuguerra and R. Chow, ‘How to analyze the visual analogue scale: Myths, truths and clinical relevance,’ Scandinavian Journal of Pain, vol. 13, no. 1, pp. 67–75, Oct. 2016. doi: 10.1016/j.sjpain.2016.06.012. [Online]. Available: 10.1016/j.sjpain.2016.06.012.

[11] T. M. Liddell and J. K. Kruschke, ‘Analyzing ordinal data with metric models: What could possibly go wrong?’ en, Journal of Experimental Social Psychology, vol. 79, pp. 328–348, Nov. 2018, issn: 0022-1031. doi: 10.1016/j.jesp.2018.08.009.

[12] C. J. Mun, H. W. Suk, M. C. Davis et al., ‘Investigating intraindividual pain variability: Methods, applications, issues, and directions,’ Pain, vol. 160, 11 2019, issn: 18726623. doi: 10.1097/j.pain.0000000000001626.

[13] X. Tan, M. P. Shiyko, R. Li, Y. Li and L. Dierker, ‘A time-varying effect model for intensive longitudinal data.,’ Psychological Methods, vol. 17, no. 1, pp. 61–77, 2012. doi: 10.1037/a0025814.

[14] S. N. Wood, N. Pya and B. Säfken, ‘Smoothing parameter and model selection for general smooth models,’ Journal of the American Statistical Association, vol. 111, no. 516, pp. 1548–1563, Oct. 2016. doi: 10.1080/01621459.2016.1180986. [Online]. Available: https://doi.org/10.1080/01621459.2016.1180986.

[15] P. Eilers, B. Marx and M. Durbán, ‘Twenty years of P-splines,’ SORT (Statistics and Operations Research Transactions), vol. 39, pp. 149–186, Jan. 2015. [Online]. Available: http://statweb.lsu.edu/faculty/marx/SORTTwentyYears.pdf.

[16] K. L. Druce, J. McBeth, S. N. van der Veer et al., ‘Recruitment and ongoing engagement in a UK smartphone study examining the association between weather and pain: Cohort study,’ JMIR mHealth and uHealth, vol. 5, no. 11, e168, Nov. 2017. doi: 10.2196/mhealth.8162.

[17] S. Wood, Generalized additive models: an introduction with R. Boca Raton: CRC Press/Taylor & Francis Group, 2017, isbn. 9781.98728331.

