## Supplementary material for "Describing variability of intensively collected longitudinal ordinal data with latent spline models": R code for simulations + LaTeX source files: examples.R

```
n <- 100     # number of timepoints
n_subj <- 8 # number of subjects
n_ord <- 5 # number of ordinal levels
set.seed(1)

cutpoints <- 2 * qnorm(seq(n_ord - 1) / n_ord)

# Simulate some patients
library(dplyr)
simulation <- tibble(
  subject = rep(LETTERS[1:n_subj], each = n),
  day = rep(1:n, times = n_subj),
  wx = c(rep(0, n),
         rep(0, n),
         rep(0, n),
         seq(-2, 2, length = 100),
         seq(2, -2, length = 100),
         1.0 * sin(seq(-pi, pi, length = 100)),
         1.0 * sin(seq(-pi, 3*pi, length = 100)),
         2 * sin(seq(-pi, pi, length = 100)))
) %>%
  mutate(epsilon = rnorm(n_subj * n,
                         mean = 0,
                         sd = c(rep(0, n), rep(.5, n), rep(1, n),
                                rep(.7, n * 2),
                                rep(.7, n * 3))),
         latent = wx + epsilon,
         y = ordered(1 + findInterval(latent, cutpoints)))

library(ggplot2)

# Plot the true latent curves. For reference only.
theme_set(theme_classic())
ggplot(simulation) +
  aes(day, latent) +
  geom_hline(yintercept = cutpoints, linetype = 'dashed') +
  geom_line() +
  geom_point() +
  facet_wrap(~subject)

# Plot the observed pain scores
ggplot(simulation) +
  aes(day, y) +
  #geom_line() +
  geom_point() +
  facet_wrap(~subject) +
  scale_x_continuous('days in study') +
  scale_y_discrete('pain severity')

# Calculate mean absolute deviation from median
madm <- function(x) {
  mean(abs(x - median(x)))
}

summaries <- simulation %>%
  group_by(subject) %>%
  mutate(y = as.numeric(y)) %>%
  summarise(n = n(),
            Median = median(y),
            Mean = mean(y),
            ISD = sd(y),
            MADM = madm(y))

# Fit latent spline models and calculate further summaries, including
# EDF
# Mean absolute deviation from expectation (where expectation is the most probable value)
# Chi^2
# Deviance explained
# 1 - MADE/MADM
library(mgcv)
get_latent <- function(model) {
  predict(model, type = 'link')
}
get_mostprob <- function(model) {
  apply(predict(model, type = 'response'), 1, which.max)
}
get_expectation <- function(model) {
  apply(predict(model, type = 'response'), 1,
        function(probs) weighted.mean(seq_along(probs), probs))
}
get_median <- function(model) {
  apply(predict(model, type = 'response'), 1,
        function(probs) which.max(cumsum(probs) > 0.5))
}
made <- function(model) {
  # mean difference between observed value and median prediction
  mean(abs(model$y - get_median(model)))
  #mean(abs(model$y - get_mostprob(model)))
  #mean(abs(model$y - get_expectation(model)))
}
made2 <- function(model) {
  # mean difference between observed value and weighted mean of pred * prob
  mean(abs(model$y - get_expectation(model)))
}

spline_models <- simulation %>%
  filter(subject != 'A') %>%
  group_by(subject) %>%
  mutate(y = as.numeric(y)) %>%
  summarise(model = list(gam(y ~ s(day, bs = 'ps'), family = ocat(R = n_ord))))

# Calculate model summary statistics
model_summaries <-
  spline_models %>%
    mutate(EDF = sapply(model, \(m) sum(m$edf) - 1),
           Dev.Expl = sapply(model, \(m) summary(m)$dev.expl),
           Chi.Sq = sapply(model, \(m) summary(m)$chi.sq),
           MADE = sapply(model, made)) %>%
  select(-model) %>%
  full_join(summaries, by = 'subject') %>%
  mutate(RelRed = 1 - MADE / MADM) %>%
  arrange(subject)
model_summaries

# Get fitted values for plotting
# fitted <- spline_models %>%
#   group_by(subject) %>%
#   group_modify(~broom::augment(.x$model[[1]]))

# Get fitted values in observation space
probs <- spline_models %>%
  group_by(subject) %>%
  group_modify(~ predict(.x$model[[1]], type = 'response') |>
                 as.data.frame()) %>%
  mutate(day = row_number()) %>%  ## Assumes no missing values
  tidyr::pivot_longer(starts_with('V'),
                      names_to = 'y', values_to = 'probability') %>%
  mutate(y = stringr::str_remove(y, 'V'))

fitted <- probs %>%
  group_by(subject, day) %>%
  summarise(plot_mean = weighted.mean(as.numeric(y), probability))

# Most probable values
ggplot(simulation) +
  aes(day, as.numeric(y)) +
  geom_tile(aes(day, y, alpha = probability), data = probs) +
  geom_point() +
  geom_step(data = probs %>% group_by(subject, day) %>%
              top_n(1, probability),
            colour = 'tomato2') +
  geom_line(data = fitted, aes(y = plot_mean), colour = 'blue') +
  facet_wrap(~subject) +
  scale_fill_continuous(limits = 0:1)
```
