## Supplementary material for "Describing variability of intensively collected longitudinal ordinal data with latent spline models": R code for simulations + LaTeX source files: references.bib

@book{Stroup_GLMMs_2024,
  title = {Generalized Linear Mixed Models: Modern Concepts, Methods and Applications},
  ISBN = {9780429092060},
  url = {http://dx.doi.org/10.1201/9780429092060},
  DOI = {10.1201/9780429092060},
  publisher = {Chapman and Hall/CRC},
  author = {Stroup,  Walter W. and Ptukhina,  Marina and Garai,  Julie},
  year = {2024},
  month = mar,
  edition = {Second}
}

@article{Wang2019,
  doi = {10.1055/s-0039-1677908},
  url = {https://doi.org/10.1055/s-0039-1677908},
  year = {2019},
  month = aug,
  publisher = {Georg Thieme Verlag {KG}},
  volume = {28},
  number = {01},
  pages = {016--026},
  author = {Fei Wang and Anita Preininger},
  title = {{AI} in Health: State of the Art,  Challenges,  and Future Directions},
  journal = {Yearbook of Medical Informatics}
}

@ARTICLE{Klasnja2015microrandomized,
  title     = "Microrandomized trials: An experimental design for developing
               just-in-time adaptive interventions",
  author    = "Klasnja, Predrag and Hekler, Eric B and Shiffman, Saul and
               Boruvka, Audrey and Almirall, Daniel and Tewari, Ambuj and
               Murphy, Susan A",
  abstract  = "OBJECTIVE: This article presents an experimental design, the
               microrandomized trial, developed to support optimization of
               just-in-time adaptive interventions (JITAIs). JITAIs are mHealth
               technologies that aim to deliver the right intervention
               components at the right times and locations to optimally support
               individuals' health behaviors. Microrandomized trials offer a
               way to optimize such interventions by enabling modeling of
               causal effects and time-varying effect moderation for individual
               intervention components within a JITAI. METHOD: The article
               describes the microrandomized trial design, enumerates research
               questions that this experimental design can help answer, and
               provides an overview of the data analyses that can be used to
               assess the causal effects of studied intervention components and
               investigate time-varying moderation of those effects. RESULTS:
               Microrandomized trials enable causal modeling of proximal
               effects of the randomized intervention components and assessment
               of time-varying moderation of those effects. CONCLUSION:
               Microrandomized trials can help researchers understand whether
               their interventions are having intended effects, when and for
               whom they are effective, and what factors moderate the
               interventions' effects, enabling creation of more effective
               JITAIs.",
  journal   = "Health Psychol.",
  publisher = "American Psychological Association (APA)",
  volume    = "34S",
  number    = "Suppl",
  pages     = "1220--1228",
  month     =  dec,
  year      =  2015,
  language  = "en"
}

@article{Okun2018,
  doi = {10.1186/s12938-018-0553-x},
  url = {https://doi.org/10.1186/s12938-018-0553-x},
  year = {2018},
  month = sep,
  publisher = {Springer Science and Business Media {LLC}},
  volume = {17},
  number = {1},
  author = {Sally Okun and Paul Wicks},
  title = {{DigitalMe}: a journey towards personalized health and thriving},
  journal = {{BioMedical} Engineering {OnLine}}
}

@article{Marcolino2018,
  url = {https://doi.org/10.2196/mhealth.8873},
  year = {2018},
  month = jan,
  publisher = {{JMIR} Publications Inc.},
  volume = {6},
  number = {1},
  pages = {e23},
  author = {Milena Soriano Marcolino and Jo{\~{a}}o Antonio Queiroz Oliveira and Marcelo D{\textquotesingle}Agostino and Antonio Luiz Ribeiro and Maria Beatriz Moreira Alkmim and David Novillo-Ortiz},
  title = {The Impact of {mHealth} Interventions: Systematic Review of Systematic Reviews},
  journal = {{JMIR} {mHealth} and {uHealth}}
}

@article{mun:suk:etal:2019,
   abstract = {Pain is a dynamic experience subject to substantial individual differences. Intensive longitudinal designs best capture the dynamical ebb and flow of the pain experience across time and settings. Thanks to the development of innovative and efficient data collection technologies, conducting an intensive longitudinal pain study has become increasingly feasible. However, the majority of longitudinal studies have tended to examine average level of pain as a predictor or as an outcome, while conceptualizing intraindividual pain variation as noise, error, or a nuisance factor. Such an approach may miss the opportunity to understand how fluctuations in pain over time are associated with pain processing, coping, other indices of adjustment, and treatment response. The present review introduces the 4 most frequently used intraindividual variability indices: The intraindividualSD/variance, autocorrelation, the mean square of successive difference, and probability of acute change. In addition, we discuss recent development in dynamic structural equation modeling in a nontechnical manner. We also consider some notable methodological issues, present a real-world example of intraindividual variability analysis, and offer suggestions for future research. Finally,we provide statistical software syntax for calculating the aforementioned intraindividual pain variability indices so that researchers can easily apply them in their research. We believe that investigating intraindividual variability of pain will provide a new perspective for understanding the complexmechanisms underlying pain coping and adjustment, as well as for enhancing efforts in precision pain medicine. Audio accompanying this abstract is available online as supplemental digital content at http://links.lww.com/PAIN/A817.},
   author = {Chung Jung Mun and Hye Won Suk and Mary C. Davis and Paul Karoly and Patrick Finan and Howard Tennen and Mark P. Jensen},
   doi = {10.1097/j.pain.0000000000001626},
   issn = {18726623},
   issue = {11},
   journal = {Pain},
   title = {Investigating intraindividual pain variability: Methods, applications, issues, and directions},
   volume = {160},
   year = {2019},
}
@article{check-hayden:2016,
   author = {Erika Check Hayden},
   doi = {10.1038/531422a},
   issn = {14764687},
   issue = {7595},
   journal = {Nature},
   title = {Mobile-phone health apps deliver data bounty},
   volume = {531},
   year = {2016},
}
@article{dixon:beukenhorst:etal:2019,
   abstract = {Patients with chronic pain commonly believe their pain is related to the weather. Scientific evidence to support their beliefs is inconclusive, in part due to difficulties in getting a large dataset of patients frequently recording their pain symptoms during a variety of weather conditions. Smartphones allow the opportunity to collect data to overcome these difficulties. Our study Cloudy with a Chance of Pain analysed daily data from 2658 patients collected over a 15-month period. The analysis demonstrated significant yet modest relationships between pain and relative humidity, pressure and wind speed, with correlations remaining even when accounting for mood and physical activity. This research highlights how citizen-science experiments can collect large datasets on real-world populations to address long-standing health questions. These results will act as a starting point for a future system for patients to better manage their health through pain forecasts.},
   author = {William G. Dixon and Anna L. Beukenhorst and Belay B. Yimer and Louise Cook and Antonio Gasparrini and Tal El-Hay and Bruce Hellman and Ben James and Ana M. Vicedo-Cabrera and Malcolm Maclure and Ricardo Silva and John Ainsworth and Huai Leng Pisaniello and Thomas House and Mark Lunt and Carolyn Gamble and Caroline Sanders and David M. Schultz and Jamie C. Sergeant and John McBeth},
   doi = {10.1038/s41746-019-0180-3},
   issn = {23986352},
   issue = {1},
   journal = {npj Digital Medicine},
   title = {How the weather affects the pain of citizen scientists using a smartphone app},
   volume = {2},
   year = {2019},
}
@article{menni:valdes:etal:2020,
   abstract = {A total of 2,618,862 participants reported their potential symptoms of COVID-19 on a smartphone-based app. Among the 18,401 who had undergone a SARS-CoV-2 test, the proportion of participants who reported loss of smell and taste was higher in those with a positive test result (4,668 of 7,178 individuals; 65.03\%) than in those with a negative test result (2,436 of 11,223 participants; 21.71\%) (odds ratio = 6.74; 95\% confidence interval = 6.31–7.21). A model combining symptoms to predict probable infection was applied to the data from all app users who reported symptoms (805,753) and predicted that 140,312 (17.42\%) participants are likely to have COVID-19.},
   author = {Cristina Menni and Ana M. Valdes and Maxim B. Freidin and Carole H. Sudre and Long H. Nguyen and David A. Drew and Sajaysurya Ganesh and Thomas Varsavsky and M. Jorge Cardoso and Julia S. El-Sayed Moustafa and Alessia Visconti and Pirro Hysi and Ruth C.E. Bowyer and Massimo Mangino and Mario Falchi and Jonathan Wolf and Sebastien Ourselin and Andrew T. Chan and Claire J. Steves and Tim D. Spector},
   doi = {10.1038/s41591-020-0916-2},
   issn = {1546170X},
   issue = {7},
   journal = {Nature Medicine},
   title = {Real-time tracking of self-reported symptoms to predict potential COVID-19},
   volume = {26},
   year = {2020},
}

@article{shiffman:stone:hufford:2008,
journal = {Annual review of clinical psychology}, 
title = {Real-time tracking of self-reported symptoms to predict potential COVID-19},
year = 2008,
volume = 4,
pages = {1-32},
author = {Saul Shiffman and Arthur A. Stone and Michael R. Hufford}
}

% Encoding: UTF-8
@article{McHugh2019,
  doi = {10.1097/pr9.0000000000000747},
  url = {https://doi.org/10.1097/pr9.0000000000000747},
  year = {2019},
  month = may,
  publisher = {Ovid Technologies (Wolters Kluwer Health)},
  volume = {4},
  number = {3},
  pages = {e747},
  author = {R. Kathryn McHugh and Robert R. Edwards and Edgar L. Ross and Robert N. Jamison},
  title = {Does bedtime matter among patients with chronic pain? A longitudinal comparison study},
  journal = {{PAIN} Reports}
}

@article{Ali2020,
  doi = {10.1002/ejp.1688},
  url = {https://doi.org/10.1002/ejp.1688},
  year = {2020},
  month = nov,
  publisher = {Wiley},
  volume = {25},
  number = {2},
  pages = {327--338},
  author = {Syed Mustafa Ali and Wei J. Lau and John McBeth and William G. Dixon and Sabine N. van der Veer},
  title = {Digital manikins to self-report pain on a smartphone: A systematic review of mobile apps},
  journal = {European Journal of Pain}
}

@article{Yang2018,
  doi = {10.1109/jbhi.2017.2776351},
  url = {https://doi.org/10.1109/jbhi.2017.2776351},
  year = {2018},
  month = nov,
  publisher = {Institute of Electrical and Electronics Engineers ({IEEE})},
  volume = {22},
  number = {6},
  pages = {1711--1719},
  author = {Geng Yang and Mingzhe Jiang and Wei Ouyang and Guangchao Ji and Haibo Xie and Amir M. Rahmani and Pasi Liljeberg and Hannu Tenhunen},
  title = {{IoT}-Based Remote Pain Monitoring System: From Device to Cloud Platform},
  journal = {{IEEE} Journal of Biomedical and Health Informatics}
}

@inproceedings{Kong2020,
  doi = {10.1109/embc44109.2020.9176077},
  url = {https://doi.org/10.1109/embc44109.2020.9176077},
  year = {2020},
  month = jul,
  publisher = {{IEEE}},
  author = {Youngsun Kong and Hugo F. Posada-Quintero and Ki H. Chon},
  title = {Pain Detection using a Smartphone in Real Time},
  booktitle = {2020 42nd Annual International Conference of the {IEEE} Engineering in Medicine {\&} Biology Society ({EMBC})}
}

@article{Jamison2017,
  doi = {10.1097/ajp.0000000000000455},
  url = {https://doi.org/10.1097/ajp.0000000000000455},
  year = {2017},
  month = aug,
  publisher = {Ovid Technologies (Wolters Kluwer Health)},
  volume = {33},
  number = {8},
  pages = {676--686},
  author = {Robert N. Jamison and Dylan C. Jurcik and Robert R. Edwards and Chuan-Chin Huang and Edgar L. Ross},
  title = {A Pilot Comparison of a Smartphone App With or Without 2-Way Messaging Among Chronic Pain Patients},
  journal = {The Clinical Journal of Pain}
}

@article{Lalloo2019,
  doi = {10.2196/11838},
  url = {https://doi.org/10.2196/11838},
  year = {2019},
  month = jan,
  publisher = {{JMIR} Publications Inc.},
  volume = {7},
  number = {1},
  pages = {e11838},
  author = {Chitra Lalloo and Amos Hundert and Lauren Harris and Quynh Pham and Fiona Campbell and Jill Chorney and Bruce Dick and Mark Simmonds and Joseph Cafazzo and Jennifer Stinson},
  title = {Capturing Daily Disease Experiences of Adolescents With Chronic Pain: {mHealth}-Mediated Symptom Tracking},
  journal = {{JMIR} {mHealth} and {uHealth}}
}

@article{Yang2019,
  doi = {10.4258/hir.2019.25.4.332},
  url = {https://doi.org/10.4258/hir.2019.25.4.332},
  year = {2019},
  publisher = {The Korean Society of Medical Informatics},
  volume = {25},
  number = {4},
  pages = {332},
  author = {Yong Sook Yang and Gi Wook Ryu and Mona Choi},
  title = {Factors Associated with Daily Completion Rates in a Smartphone-Based Ecological Momentary Assessment Study},
  journal = {Healthcare Informatics Research}
}

@article{Ono2019,
  doi = {10.2196/11398},
  url = {https://doi.org/10.2196/11398},
  year = {2019},
  month = feb,
  publisher = {{JMIR} Publications Inc.},
  volume = {21},
  number = {2},
  pages = {e11398},
  author = {Masakatsu Ono and Stefan Schneider and Doerte U Junghaenel and Arthur A Stone},
  title = {What Affects the Completion of Ecological Momentary Assessments in Chronic Pain Research? An Individual Patient Data Meta-Analysis},
  journal = {Journal of Medical Internet Research}
}

@article{Shaygan2021,
  doi = {10.1038/s41598-021-86156-8},
  url = {https://doi.org/10.1038/s41598-021-86156-8},
  year = {2021},
  month = mar,
  publisher = {Springer Science and Business Media {LLC}},
  volume = {11},
  number = {1},
  author = {Maryam Shaygan and Azita Jaberi},
  title = {The effect of a smartphone-based pain management application on pain intensity and quality of life in adolescents with chronic pain},
  journal = {Scientific Reports}
}

@Misc{ordinal,
  author = {R. H. B. Christensen},
  note   = {R package version 2019.12-10. https://CRAN.R-project.org/package=ordinal},
  title  = {ordinal---Regression Models for Ordinal Data},
  year   = {2019},
}

@article{1978,
  title = {Intraindividual Differences Dimensions of Mood Change during Pregnancy Identified in Five {{P}}-Technique Factor Analyses},
  year = {1978},
  month = jun,
  volume = {12},
  pages = {205--224},
  publisher = {{Academic Press}},
  issn = {0092-6566},
  doi = {10.1016/0092-6566(78)90098-3},
  abstract = {Daily-self-ratings of mood and state were obtained for approximately 120 days of observation from five pregnant subjects. Measurements were based on a\ldots},
  file = {C\:\\Users\\David\\Zotero\\storage\\KDSYFJEP\\0092656678900983.html},
  journal = {Journal of Research in Personality},
  language = {en},
  number = {2}
}

@Article{Schmidt2010,
  author    = {Schmidt, Carsten Oliver and Raspe, Heiner and Kohlmann, Thomas},
  journal   = {PAIN},
  title     = {Graded Back Pain Revisited \textendash{} {{Do}} Latent Variable Models Change Our Understanding of Severe Back Pain in the General Population?},
  year      = {2010},
  issn      = {0304-3959},
  month     = apr,
  number    = {1},
  pages     = {50--56},
  volume    = {149},
  abstract  = {Back pain severity has extensively been targeted in clinical and epidemiologic studies. However, despite the importance of a valid pain severity gradi\ldots},
  doi       = {10.1016/j.pain.2010.01.025},
  language  = {en},
  publisher = {Lippincott Williams & Wilkins, Inc.},
}

@article{2014,
  title = {{{GEE}} for Longitudinal Ordinal Data: {{Comparing R}}-Geepack, {{R}}-Multgee, {{R}}-Repolr, {{SAS}}-{{GENMOD}}, {{SPSS}}-{{GENLIN}}},
  shorttitle = {{{GEE}} for Longitudinal Ordinal Data},
  year = {2014},
  month = sep,
  volume = {77},
  pages = {70--83},
  publisher = {{North-Holland}},
  issn = {0167-9473},
  doi = {10.1016/j.csda.2014.03.009},
  abstract = {Studies in epidemiology and social sciences are often longitudinal and outcome measures are frequently obtained by questionnaires in ordinal scales. T\ldots},
  file = {C\:\\Users\\David\\Zotero\\storage\\NPAXTDDB\\2014 - GEE for longitudinal ordinal data Comparing R-gee.pdf},
  journal = {Computational Statistics \& Data Analysis},
  language = {en}
}

@article{Affleck1991,
  title = {Individual Differences in the Day-to-Day Experience of Chronic Pain: A Prospective Daily Study of Rheumatoid Arthritis Patients.},
  author = {Affleck, Glenn and Tennen, Howard and Urrows, Susan and Higgins, Pamela},
  year = {1991},
  volume = {10},
  pages = {419--426},
  publisher = {{American Psychological Association (APA)}},
  doi = {10.1037/0278-6133.10.6.419},
  journal = {Health Psychology},
  number = {6}
}

@article{Aguilar,
  title = {Bayesian {{Inference}} on {{Latent Structure}} in {{Time Series}}},
  author = {Aguilar, Omar and Huerta, Gabriel and Prado, Raquel and West, Mike},
  pages = {16},
  abstract = {A range of developments in Bayesian time series modelling in recent years has focussed on issues of identifying latent structure in time series. This has led to new uses and interpretations of existing theory for latent process decompositions of dynamic models, and to new models for univariate and multivariate time series. This article draws together concepts and modelling approaches that are central to applications of time series decomposition methods, and reviews recent modelling and applied developments. Several applications in time series analyses in geology, climatology, psychiatry and finance are discussed, as are related modelling directions and current research frontiers.},
  file = {C\:\\Users\\David\\Zotero\\storage\\Z44DW3II\\Aguilar et al. - Bayesian Inference on Latent Structure in Time Ser.pdf},
  language = {en}
}

@article{Asparouhov2017,
  title = {Dynamic {{Structural Equation Models}}},
  author = {Asparouhov, Tihomir and Hamaker, Ellen L. and Muth{\'e}n, Bengt},
  year = {2017},
  month = dec,
  volume = {25},
  pages = {359--388},
  publisher = {{Informa UK Limited}},
  doi = {10.1080/10705511.2017.1406803},
  journal = {Structural Equation Modeling: A Multidisciplinary Journal},
  number = {3}
}

@Article{Beyaztas2019,
  author        = {Beyaztas, Ufuk and Shang, Han Lin},
  journal       = {Journal of Statistical Computation and Simulation},
  title         = {Forecasting Functional Time Series Using Weighted Likelihood Methodology},
  year          = {2019},
  issn          = {0094-9655, 1563-5163},
  month         = nov,
  number        = {16},
  pages         = {3046--3060},
  volume        = {89},
  abstract      = {Functional time series whose sample elements are recorded sequentially over time are frequently encountered with increasing technology. Recent studies have shown that analyzing and forecasting of functional time series can be performed easily using functional principal component analysis and existing univariate/multivariate time series models. However, the forecasting performance of such functional time series models may be affected by the presence of outlying observations which are very common in many scientific fields. Outliers may distort the functional time series model structure, and thus, the underlying model may produce high forecast errors. We introduce a robust forecasting technique based on weighted likelihood methodology to obtain point and interval forecasts in functional time series in the presence of outliers. The finite sample performance of the proposed method is illustrated by Monte Carlo simulations and four real-data examples. Numerical results reveal that the proposed method exhibits superior performance compared with the existing method(s).},
  archiveprefix = {arXiv},
  doi           = {10.1080/00949655.2019.1650935},
  eprint        = {1908.00336},
  eprinttype    = {arxiv},
  file          = {C\:\\Users\\David\\Zotero\\storage\\E8FKLMNT\\Beyaztas and Shang - 2019 - Forecasting functional time series using weighted .pdf;C\:\\Users\\David\\Zotero\\storage\\YK423YB4\\1908.html},
  keywords      = {97K80, Statistics - Methodology},
}

@article{Bianconcini2018,
  title = {The {{Latent Variable}}-{{Autoregressive Latent Trajectory Model}}: {{A General Framework}} for {{Longitudinal Data Analysis}}},
  author = {Bianconcini, Silvia and Bollen, Kenneth A.},
  year = {2018},
  month = jan,
  volume = {25},
  pages = {791--808},
  publisher = {{Informa UK Limited}},
  doi = {10.1080/10705511.2018.1426467},
  journal = {Structural Equation Modeling: A Multidisciplinary Journal},
  number = {5}
}

@article{Bokil2006,
  title = {Dynamic Phenotypes},
  author = {Bokil, Hemant and Tchernichovsky, Ofer and Mitra, Partha P.},
  year = {2006},
  month = mar,
  volume = {4},
  pages = {119--128},
  publisher = {{Humana Press}},
  issn = {1559-0089},
  doi = {10.1385/NI:4:1:119},
  abstract = {We consider quantitative measures of behavioral and neuronal dynamics as a means of characterizing phenotypes. Such measures are important from a scientific perspective; because understanding brain function is contingent on understanding the link between the dynamics of the nervous system and behavioral dynamics. They are also important from a biomedical perspective because they provide a contrast to purely psychological characterizations of phenotype or characterizations via static brain images or maps, and are a potential means for differential diagnoses of neuropsychiatric illnesses. After a brief presentation of background work and some current advances, we suggest that more attention needs to be paid to dynamic characterizations of phenotypes. We will discuss some of the relevant time series analysis tools.},
  copyright = {2006 Humana Press Inc},
  file = {C\:\\Users\\David\\Zotero\\storage\\RYBBIEFQ\\Bokil et al. - 2006 - Dynamic phenotypes.pdf;C\:\\Users\\David\\Zotero\\storage\\YWNVNXNK\\10.html},
  journal = {Neuroinformatics},
  language = {en},
  number = {1}
}

@inproceedings{Brillinger1996,
  title = {An Analysis of an Ordinal-Valued Time Series},
  booktitle = {Athens {{Conference}} on {{Applied Probability}} and {{Time Series Analysis}}},
  author = {Brillinger, David R.},
  editor = {Robinson, P. M. and Rosenblatt, Murray},
  year = {1996},
  pages = {73--87},
  publisher = {{Springer}},
  address = {{New York, NY}},
  doi = {10.1007/978-1-4612-2412-9_6},
  abstract = {Time series and spatial processes are sometimes ordinal-valued. It can be convenient to handle such types of data via generalized linear model algorithms employing the complimentary loglog link function. This approach facilitates the use of standard statistical packages and leads to a convenient technique for handling serial dependence. Model fit is assessed by uniform residuals, amongst other tools. In this article an example of such an analysis is provided for a three-valued series corresponding to the possible results loss, tie, win of events involving a sports team.},
  isbn = {978-1-4612-2412-9},
  language = {en},
  series = {Lecture {{Notes}} in {{Statistics}}}
}

@article{Cazelles2007,
  title = {Time-Dependent Spectral Analysis of Epidemiological Time-Series with Wavelets},
  author = {Cazelles, Bernard and Chavez, Mario and de Magny, Guillaume Constantin and Gu{\'e}gan, Jean-Francois and Hales, Simon},
  year = {2007},
  month = aug,
  volume = {4},
  pages = {625--636},
  publisher = {{Royal Society}},
  doi = {10.1098/rsif.2007.0212},
  abstract = {In the current context of global infectious disease risks, a better understanding of the dynamics of major epidemics is urgently needed. Time-series analysis has appeared as an interesting approach to explore the dynamics of numerous diseases. Classical time-series methods can only be used for stationary time-series (in which the statistical properties do not vary with time). However, epidemiological time-series are typically noisy, complex and strongly non-stationary. Given this specific nature, wavelet analysis appears particularly attractive because it is well suited to the analysis of non-stationary signals. Here, we review the basic properties of the wavelet approach as an appropriate and elegant method for time-series analysis in epidemiological studies. The wavelet decomposition offers several advantages that are discussed in this paper based on epidemiological examples. In particular, the wavelet approach permits analysis of transient relationships between two signals and is especially suitable for gradual change in force by exogenous variables.},
  file = {C\:\\Users\\David\\Zotero\\storage\\8C2KIMJU\\Cazelles et al. - 2007 - Time-dependent spectral analysis of epidemiologica.pdf;C\:\\Users\\David\\Zotero\\storage\\3TZHGLPA\\rsif.2007.html},
  journal = {Journal of The Royal Society Interface},
  number = {15}
}

@article{Chen2014,
  title = {Is {{Rasch}} Model Analysis Applicable in Small Sample Size Pilot Studies for Assessing Item Characteristics? {{An}} Example Using {{PROMIS}} Pain Behavior Item Bank Data},
  shorttitle = {Is {{Rasch}} Model Analysis Applicable in Small Sample Size Pilot Studies for Assessing Item Characteristics?},
  author = {Chen, Wen-Hung and Lenderking, William and Jin, Ying and Wyrwich, Kathleen W. and Gelhorn, Heather and Revicki, Dennis A.},
  year = {2014},
  month = mar,
  volume = {23},
  pages = {485--493},
  publisher = {{Springer International Publishing}},
  issn = {1573-2649},
  doi = {10.1007/s11136-013-0487-5},
  abstract = {Purpose Large samples are generally considered necessary for Rasch model to obtain robust item parameter estimates. Recently, small sample Rasch analysis was suggested as preliminary assessment of items' psychometric properties. This study is to evaluate the Rasch analysis results using small sample sizes. Methods Ten PROMIS pain behavior items were used. Random samples of 30, 50, 100, and 250, and a targeted sample of 30 were drawn 10 times each from a total of 800 subjects. Rasch analysis was conducted for each of these samples and the full sample. Results In the full sample, there were 104 cases of extreme scores, no null categories, two incorrectly ordered items, and four misfit items. For samples of 250, 100, 50, 30, and targeted 30, the average numbers of extreme scores were 42.2, 17.1, 9.6, 6.1, and 1.2; the average numbers of null categories were 1.0, 3.2, 8.7, 13.4, and 8.3; the average numbers of items with incorrectly ordered item parameters were 0.1, 0.8, 2.9, 4.7, and 3.7; and the average numbers of items with fit residuals exceeding {$\pm$}2.5 were 0.8, 0.3, 0.1, 0.2, and 0.3, respectively. Conclusions Rasch analysis based on small samples ({$\leq$}50) identified a greater number of items with incorrectly ordered parameters than larger samples ({$\geq$}100). However, fewer items were identified as misfitting. Results from small samples led to opposite conclusions from those based on larger samples. Rasch analysis based on small samples should be used for exploratory purposes with extreme caution.},
  copyright = {2013 Springer Science+Business Media Dordrecht},
  file = {C\:\\Users\\David\\Zotero\\storage\\6C3FYURT\\Chen et al. - 2014 - Is Rasch model analysis applicable in small sample.pdf;C\:\\Users\\David\\Zotero\\storage\\UAZLK7K5\\10.html},
  journal = {Quality of Life Research},
  language = {en},
  number = {2}
}

@article{Chen2019,
  title = {A {{Latent Gaussian}} Process Model for Analysing Intensive Longitudinal Data},
  author = {Chen, Yunxiao and Zhang, Siliang},
  year = {2019},
  month = aug,
  volume = {73},
  pages = {237--260},
  publisher = {{Wiley}},
  doi = {10.1111/bmsp.12180},
  journal = {British Journal of Mathematical and Statistical Psychology},
  number = {2}
}

@Article{Chen2020,
  author     = {Chen, Yunxiao and Zhang, Siliang},
  journal    = {British Journal of Mathematical and Statistical Psychology},
  title      = {A {{Latent Gaussian}} Process Model for Analysing Intensive Longitudinal Data},
  year       = {2020},
  issn       = {2044-8317},
  number     = {2},
  pages      = {237--260},
  volume     = {73},
  abstract   = {Intensive longitudinal studies are becoming progressively more prevalent across many social science areas, and especially in psychology. New technologies such as smart-phones, fitness trackers, and the Internet of Things make it much easier than in the past to collect data for intensive longitudinal studies, providing an opportunity to look deep into the underlying characteristics of individuals under a high temporal resolution. In this paper we introduce a new modelling framework for latent curve analysis that is more suitable for the analysis of intensive longitudinal data than existing latent curve models. Specifically, through the modelling of an individual-specific continuous-time latent process, some unique features of intensive longitudinal data are better captured, including intensive measurements in time and unequally spaced time points of observations. Technically, the continuous-time latent process is modelled by a Gaussian process model. This model can be regarded as a semi-parametric extension of the classical latent curve models and falls under the framework of structural equation modelling. Procedures for parameter estimation and statistical inference are provided under an empirical Bayes framework and evaluated by simulation studies. We illustrate the use of the proposed model though the analysis of an ecological momentary assessment data set.},
  annotation = {\_eprint: https://bpspsychub.onlinelibrary.wiley.com/doi/pdf/10.1111/bmsp.12180},
  copyright  = {\textcopyright{} 2019 The British Psychological Society},
  doi        = {10.1111/bmsp.12180},
  file       = {C\:\\Users\\David\\Zotero\\storage\\VJJN4U7N\\Chen and Zhang - 2020 - A Latent Gaussian process model for analysing inte.pdf;C\:\\Users\\David\\Zotero\\storage\\FP7GHGUJ\\bmsp.html},
  keywords   = {ecological momentary assessment, Gaussian process, intensive longitudinal data, latent curve analysis, structural equation modelling, time-varying latent trait},
  language   = {en},
}

@article{Chiou2007,
  title = {Functional Clustering and Identifying Substructures of Longitudinal Data},
  author = {Chiou, Jeng-Min and Li, Pai-Ling},
  year = {2007},
  month = sep,
  volume = {69},
  pages = {679--699},
  publisher = {{Wiley}},
  doi = {10.1111/j.1467-9868.2007.00605.x},
  journal = {Journal of the Royal Statistical Society: Series B (Statistical Methodology)},
  number = {4}
}

@article{Costa2021,
  title = {{{ISSLS PRIZE IN CLINICAL SCIENCE}} 2021: {{What}} Are the Risk Factors for Low Back Pain Flares and Does This Depend on How Flare Is Defined?},
  shorttitle = {{{ISSLS PRIZE IN CLINICAL SCIENCE}} 2021},
  author = {Costa, Nathalia and Smits, Esther and Kasza, Jessica and Salomoni, Sauro and Ferreira, Manuela and Sullivan, Michael and Hodges, Paul W.},
  year = {2021},
  month = feb,
  issn = {1432-0932},
  doi = {10.1007/s00586-021-06730-6},
  abstract = {Although risk factors for new low back pain (LBP) episodes and acute-to-chronic transition have been identified, risk factors for flares of LBP remain largely unknown. This case-crossover study aimed to identify: (1) risk factors LBP flares and (2) whether risk factors differed when flare is defined by pain increase (pain-defined flare: PDF) or identified by participants according to a broader flare definition that considered emotions and coping (self-reported flare: SRF).},
  file = {C\:\\Users\\David\\Zotero\\storage\\AJ4FYE55\\Costa et al. - 2021 - ISSLS PRIZE IN CLINICAL SCIENCE 2021 What are the.pdf},
  journal = {European Spine Journal},
  language = {en}
}

@article{Crawford2015,
  title = {Sex, Lies and Self-Reported Counts: {{Bayesian}} Mixture Models for Heaping in Longitudinal Count Data via Birth-Death Processes},
  shorttitle = {Sex, Lies and Self-Reported Counts},
  author = {Crawford, Forrest W. and Weiss, Robert E. and Suchard, Marc A.},
  year = {2015},
  month = jun,
  volume = {9},
  issn = {1932-6157},
  doi = {10.1214/15-AOAS809},
  abstract = {Surveys often ask respondents to report nonnegative counts, but respondents may misremember or round to a nearby multiple of 5 or 10. This phenomenon is called heaping, and the error inherent in heaped self-reported numbers can bias estimation. Heaped data may be collected cross-sectionally or longitudinally and there may be covariates that complicate the inferential task. Heaping is a well-known issue in many survey settings, and inference for heaped data is an important statistical problem. We propose a novel reporting distribution whose underlying parameters are readily interpretable as rates of misremembering and rounding. The process accommodates a variety of heaping grids and allows for quasi-heaping to values nearly but not equal to heaping multiples. We present a Bayesian hierarchical model for longitudinal samples with covariates to infer both the unobserved true distribution of counts and the parameters that control the heaping process. Finally, we apply our methods to longitudinal self-reported counts of sex partners in a study of high-risk behavior in HIV-positive youth.},
  archiveprefix = {arXiv},
  eprint = {1405.4265},
  eprinttype = {arxiv},
  file = {C\:\\Users\\David\\Zotero\\storage\\T62DTST9\\Crawford et al. - 2015 - Sex, lies and self-reported counts Bayesian mixtu.pdf;C\:\\Users\\David\\Zotero\\storage\\VU3SAUEJ\\1405.html},
  journal = {The Annals of Applied Statistics},
  keywords = {Statistics - Applications},
  number = {2}
}

@article{Dartel2014,
  title = {{{THU0263 Factors Associated}} with {{Fatigue}} in {{Patients}} with {{Rheumatoid Arthritis}}: {{A Multidimensional}} ``{{Path Analysis}}'' {{Model}}},
  shorttitle = {{{THU0263 Factors Associated}} with {{Fatigue}} in {{Patients}} with {{Rheumatoid Arthritis}}},
  author = {Dartel, S. Rongen-van and {Repping-Wuts}, H. and Donders, R. and van Hoogmoed, D. and Knoop, H. and Bleijenberg, G. and van Riel, P. and Fransen, J.},
  year = {2014},
  month = jun,
  volume = {73},
  pages = {274--274},
  publisher = {{BMJ Publishing Group Ltd}},
  issn = {0003-4967, 1468-2060},
  doi = {10.1136/annrheumdis-2014-eular.1637},
  abstract = {Background Fatigue is one of the most commonly reported symptoms in rheumatoid arthritis (RA) and severe fatigue also occurs in patients with low disease activity. Many factors may play a role in causing fatigue in RA. Several cross-sectional and longitudinal studies showed that psychosocial factors such as self-efficacy and coping strategies, pain and limitations in daily functioning, rather than inflammation, are related to fatigue severity in RA (1-4). These factors could have a possible causal relationship with fatigue. Objectives To develop and test a multidimensional model of factors that determine fatigue severity (CIS-fatigue) in RA using ``path analysis'', with the aim to facilitate the development of a treatment strategy for RA fatigue. Methods A cross-sectional study (n=228) of consecutively included patients with RA was performed. Patient and disease characteristics, fatigue level, personality factors and psychosocial variables were collected (fig 1). Structural equation modeling (``path analysis'') was used to test a hypothesized model for fatigue. The hypothesized model of fatigue included latent variables of pain, physical functioning, mood, sense of control, sleep disturbances and fatigue. {$<$}img class="highwire-fragment fragment-image" alt="Figure 1." src="https://ard.bmj.com/content/annrheumdis/73/Suppl\_2/274.1/F1.medium.gif" width="440" height="357"/{$>$}Download figure Open in new tab Download powerpoint Figure 1. The final model with standardized correlation coefficients. *p{$<$}0.01. Results The final model included 4 latent variables and had a moderate fit (CFI=0.943) (figure 1). According to the model a higher pain level ({$\beta$}=0.16, p=0.274) and lower physical functioning ({$\beta$}=-0.61, p=0.002) leads to a higher fatigue level. In addition, more mood disturbance is related to a higher fatigue level ({$\beta$}=0.253, p=0.003) and a worse sense of control is related to more mood disturbance ({$\beta$}= -0.58, p{$\leq$}0.001). The final model including pain, mood and physical functioning explained 72\% of the variance of fatigue in RA. Conclusions According to the multidimensional model, RA fatigue is influenced by pain, sense of control, mood and physical functioning. Treatment of these four factors by psychological interventions and an exercise program could help to improve fatigue in patients with RA. Randomized controlled trials, are needed to test the efficacy of these interventions. References Hewlett S, Chalder T, Choy E, Cramp F, Davis B, Dures E, et al. Fatigue in rheumatoid arthritis: time for a conceptual model. Rheumatology (Oxford). 2011;50(6):1004-6. Epub 2010/09/08. van Hoogmoed D, Fransen J, Bleijenberg G, van Riel P. Physical and psychosocial correlates of severe fatigue in rheumatoid arthritis. Rheumatology (Oxford). 2010;49(7):1294-302. Epub 2010/04/01. Treharne GJ, Lyons AC, Hale ED, Goodchild CE, Booth DA, Kitas GD. Predictors of fatigue over 1 year among people with rheumatoid arthritis. Psychol Health Med. 2008;13(4):494-504. Epub 2008/10/01. Mancuso CA, Rincon M, Sayles W, Paget SA. Psychosocial variables and fatigue: a longitudinal study comparing individuals with rheumatoid arthritis and healthy controls. J Rheumatol. 2006;33(8):1496-502. Epub 2006/06/20. Disclosure of Interest : None declared DOI 10.1136/annrheumdis-2014-eular.1637},
  chapter = {EULAR 2014: Scientific Abstracts},
  copyright = {\textcopyright{} 2014, Published by the BMJ Publishing Group Limited. For permission to use (where not already granted under a licence) please go to http://group.bmj.com/group/rights-licensing/permissions},
  file = {C\:\\Users\\David\\Zotero\\storage\\ZGAV3VMH\\274.1.html},
  journal = {Annals of the Rheumatic Diseases},
  language = {en},
  number = {Suppl 2}
}

@incollection{Diener2009,
  title = {Happiness Is the {{Frequency}}, {{Not}} the {{Intensity}}, of {{Positive Versus Negative Affect}}},
  booktitle = {Assessing {{Well}}-{{Being}}},
  author = {Diener, Ed and Sandvik, Ed and Pavot, William},
  year = {2009},
  pages = {213--231},
  publisher = {{Springer Netherlands}},
  doi = {10.1007/978-90-481-2354-4_10}
}

@article{Dixon2019,
  title = {How the Weather Affects the Pain of Citizen Scientists Using a Smartphone App},
  author = {Dixon, William G. and Beukenhorst, Anna L. and Yimer, Belay B. and Cook, Louise and Gasparrini, Antonio and {El-Hay}, Tal and Hellman, Bruce and James, Ben and {Vicedo-Cabrera}, Ana M. and Maclure, Malcolm and Silva, Ricardo and Ainsworth, John and Pisaniello, Huai Leng and House, Thomas and Lunt, Mark and Gamble, Carolyn and Sanders, Caroline and Schultz, David M. and Sergeant, Jamie C. and McBeth, John},
  year = {2019},
  month = oct,
  volume = {2},
  publisher = {{Springer Science and Business Media LLC}},
  doi = {10.1038/s41746-019-0180-3},
  journal = {npj Digital Medicine},
  number = {1}
}

@article{Flugel1925,
  title = {A Quantitative Study of Feeling and Emotion in Everyday Life},
  author = {Fl{\"u}gel, J. C.},
  year = {1925},
  month = apr,
  volume = {15},
  pages = {318--355},
  issn = {03732460},
  doi = {10.1111/j.2044-8295.1925.tb00187.x},
  journal = {British Journal of Psychology. General Section},
  language = {en},
  number = {4}
}

@article{Gates2016,
  title = {Unsupervised {{Classification During Time}}-{{Series Model Building}}},
  author = {Gates, Kathleen M. and Lane, Stephanie T. and Varangis, E. and Giovanello, K. and Guiskewicz, K.},
  year = {2016},
  month = dec,
  volume = {52},
  pages = {129--148},
  publisher = {{Informa UK Limited}},
  doi = {10.1080/00273171.2016.1256187},
  journal = {Multivariate Behavioral Research},
  number = {2}
}

@article{Gelman2000,
  title = {Type {{S}} Error Rates for Classical and {{Bayesian}} Single and Multiple Comparison Procedures},
  author = {Gelman, Andrew and Tuerlinckx, Francis},
  year = {2000},
  month = sep,
  volume = {15},
  pages = {373--390},
  publisher = {{Springer Berlin Heidelberg}},
  issn = {1613-9658},
  doi = {10.1007/s001800000040},
  abstract = {In classical statistics, the significance of comparisons (e.g., \texttheta 1- \texttheta 2) is calibrated using the Type 1 error rate, relying on the assumption that the true difference is zero, which makes no sense in many applications. We set up a more relevant framework in which a true comparison can be positive or negative, and, based on the data, you can state ``\texttheta 1 {$>$} \texttheta 2 with confidence,'' ``\texttheta 2 {$>$} \texttheta 1 with confidence,'' or ``no claim with confidence.'' We focus on the Type S (for sign) error, which occurs when you claim ``\texttheta 1 {$>$} \texttheta 2 with confidence'' when \texttheta 2{$>$} \texttheta 1 (or vice-versa). We compute the Type S error rates for classical and Bayesian confidence statements and find that classical Type S error rates can be extremely high (up to 50\%). Bayesian confidence statements are conservative, in the sense that claims based on 95\% posterior intervals have Type S error rates between 0 and 2.5\%. For multiple comparison situations, the conclusions are similar.},
  copyright = {2000 Physica-Verlag},
  file = {C\:\\Users\\David\\Zotero\\storage\\NPCIMRUR\\Gelman and Tuerlinckx - 2000 - Type S error rates for classical and Bayesian sing.pdf;C\:\\Users\\David\\Zotero\\storage\\UZ5AQJ6J\\10.html},
  journal = {Computational Statistics},
  language = {en},
  number = {3}
}

@book{Golding2019,
  title = {Greta-Dev/Greta: Greta 0.3.1},
  author = {Golding, Nick and Yen, Jian and Dirmeier, Simon and Quinn, Michael and Pullin, Jeffrey and Mulvahill, Matt and Wo{\l}od{\'z}ko, Tymoteusz and Teetor, Paul and Tierney, Nicholas and Thoen, Edwin and Marwick, Ben and Fleischhacker, Adam},
  year = {2019},
  publisher = {{Zenodo}},
  doi = {10.5281/ZENODO.3364187},
  copyright = {Open Access}
}

@article{Gruen2017,
  title = {The {{Use}} of {{Functional Data Analysis}} to {{Evaluate Activity}} in a {{Spontaneous Model}} of {{Degenerative Joint Disease Associated Pain}} in {{Cats}}},
  author = {Gruen, Margaret E. and {Alfaro-C{\'o}rdoba}, Marcela and Thomson, Andrea E. and Worth, Alicia C. and Staicu, Ana-Maria and Lascelles, B. Duncan X.},
  editor = {Harezlak, Jaroslaw},
  year = {2017},
  month = jan,
  volume = {12},
  pages = {e0169576},
  publisher = {{Public Library of Science (PLoS)}},
  doi = {10.1371/journal.pone.0169576},
  journal = {PLOS ONE},
  number = {1}
}

@article{Hall2008,
  title = {Modelling Sparse Generalized Longitudinal Observations with Latent {{Gaussian}} Processes},
  author = {Hall, Peter and M{\"u}ller, Hans-Georg and Yao, Fang},
  year = {2008},
  month = sep,
  volume = {70},
  pages = {703--723},
  publisher = {{Wiley}},
  doi = {10.1111/j.1467-9868.2008.00656.x},
  journal = {Journal of the Royal Statistical Society: Series B (Statistical Methodology)},
  number = {4}
}

@Article{Hall2008a,
  author     = {Hall, Peter and M{\"u}ller, Hans-Georg and Yao, Fang},
  journal    = {Journal of the Royal Statistical Society: Series B (Statistical Methodology)},
  title      = {Modelling Sparse Generalized Longitudinal Observations with Latent {{Gaussian}} Processes},
  year       = {2008},
  issn       = {1467-9868},
  number     = {4},
  pages      = {703--723},
  volume     = {70},
  abstract   = {Summary. In longitudinal data analysis one frequently encounters non-Gaussian data that are repeatedly collected for a sample of individuals over time. The repeated observations could be binomial, Poisson or of another discrete type or could be continuous. The timings of the repeated measurements are often sparse and irregular. We introduce a latent Gaussian process model for such data, establishing a connection to functional data analysis. The functional methods proposed are non-parametric and computationally straightforward as they do not involve a likelihood. We develop functional principal components analysis for this situation and demonstrate the prediction of individual trajectories from sparse observations. This method can handle missing data and leads to predictions of the functional principal component scores which serve as random effects in this model. These scores can then be used for further statistical analysis, such as inference, regression, discriminant analysis or clustering. We illustrate these non-parametric methods with longitudinal data on primary biliary cirrhosis and show in simulations that they are competitive in comparisons with generalized estimating equations and generalized linear mixed models.},
  annotation = {\_eprint: https://rss.onlinelibrary.wiley.com/doi/pdf/10.1111/j.1467-9868.2008.00656.x},
  copyright  = {\textcopyright{} 2008 Royal Statistical Society},
  doi        = {10.1111/j.1467-9868.2008.00656.x},
  file       = {C\:\\Users\\David\\Zotero\\storage\\B3UBGAZB\\j.1467-9868.2008.00656.html},
  keywords   = {Binomial data, Eigenfunction, Functional data analysis, Functional principal component, Prediction, Random effect, Repeated measurements, Smoothing, Stochastic process},
  language   = {en},
}

@Article{Honkoop2017,
  author     = {Honkoop, Persijn J. and Simpson, Andrew and Bonini, Matteo and {Snoeck-Stroband}, Jiska B. and Meah, Sally and Chung, Kian Fan and Usmani, Omar S. and Fowler, Stephen and Sont, Jacob K.},
  journal    = {BMJ Open},
  title      = {{{MyAirCoach}}: The Use of Home-Monitoring and {{mHealth}} Systems to Predict Deterioration in Asthma Control and the Occurrence of Asthma Exacerbations; Study Protocol of an Observational Study},
  year       = {2017},
  issn       = {2044-6055, 2044-6055},
  month      = jan,
  number     = {1},
  pages      = {e013935},
  volume     = {7},
  abstract   = {Introduction Asthma is a variable lung condition whereby patients experience periods of controlled and uncontrolled asthma symptoms. Patients who experience prolonged periods of uncontrolled asthma have a higher incidence of exacerbations and increased morbidity and mortality rates. The ability to determine and to predict levels of asthma control and the occurrence of exacerbations is crucial in asthma management. Therefore, we aimed to determine to what extent physiological, behavioural and environmental data, obtained by mobile healthcare (mHealth) and home-monitoring sensors, as well as patient characteristics, can be used to predict episodes of uncontrolled asthma and the onset of asthma exacerbations. Methods and analysis In an 1-year observational study, patients will be provided with mHealth and home-monitoring systems to record daily measurements for the first-month (phase I) and weekly measurements during a follow-up period of 11 months (phase II). Our study population consists of 150 patients, aged {$\geq$}18 years, with a clinician's diagnosis of asthma, currently on controller medication, with uncontrolled asthma and/or minimally one exacerbation in the past 12 months. They will be enrolled over three participating centres, including Leiden, London and Manchester. Our main outcomes are the association between physiological, behavioural and environmental data and (1) the loss of asthma control and (2) the occurrence of asthma exacerbations. Ethics This study was approved by the Medical Ethics Committee of the Leiden University Medical Center in the Netherlands and by the NHS ethics service in the UK. Trial registration number NCT02774772.},
  chapter    = {Respiratory medicine},
  copyright  = {Published by the BMJ Publishing Group Limited. For permission to use (where not already granted under a licence) please go to http://www.bmj.com/company/products-services/rights-and-licensing/. This is an Open Access article distributed in accordance with the Creative Commons Attribution Non Commercial (CC BY-NC 4.0) license, which permits others to distribute, remix, adapt, build upon this work non-commercially, and license their derivative works on different terms, provided the original work is properly cited and the use is non-commercial. See: http://creativecommons.org/licenses/by-nc/4.0/},
  doi        = {10.1136/bmjopen-2016-013935},
  file       = {C\:\\Users\\David\\Zotero\\storage\\DWE9FTLY\\Honkoop et al. - 2017 - MyAirCoach the use of home-monitoring and mHealth.pdf},
  keywords   = {asthma control, asthma exacerbation, ehealth, home-monitoring, mHealth, self-management},
  language   = {en},
  pmid       = {28119390},
  publisher  = {{British Medical Journal Publishing Group}},
  shorttitle = {{{MyAirCoach}}},
}

@article{Jacques2013,
  title = {Functional Data Clustering: A Survey},
  author = {Jacques, Julien and Preda, Cristian},
  year = {2013},
  month = nov,
  volume = {8},
  pages = {231--255},
  publisher = {{Springer Science and Business Media LLC}},
  doi = {10.1007/s11634-013-0158-y},
  journal = {Advances in Data Analysis and Classification},
  number = {3}
}

@Article{James2019,
  author     = {James, Richard J. E. and Walsh, David A. and Ferguson, Eamonn},
  journal    = {British Journal of Health Psychology},
  title      = {Trajectories of Pain Predict Disabilities Affecting Daily Living in Arthritis},
  year       = {2019},
  issn       = {2044-8287},
  number     = {3},
  pages      = {485--496},
  volume     = {24},
  abstract   = {Purpose To examine the interplay between pain and disability in arthritis when adjusting for patient heterogeneity in pain progression. There is consistent evidence to suggest that people experience osteoarthritis heterogeneously, with subgroups of people having different trajectories of pain. However, at present it is unclear how these pain trajectories are related to functional disability. We ask the question: Do levels of disability track changes in pain across different pain trajectories? Methods Secondary analysis of a subset (n = 889) from a cohort of older English adults, representative of the general population (the English Longitudinal Study of Ageing). The relationship between pain and functional disability was compared in three domains of disability: mobility, activities of daily living (ADL) and instrumental ADL. These represent increasingly complex forms of self-care required for independent living. Data analysis compared the heterogeneous analysis of pain (different trajectories) and disability compared to treating pain as a simpler homogenous construct. Results On a population level, pain was significantly positively correlated with increased disability in all three domains, and the relationship remained stable over time. However, when heterogeneity was examined respondents whose pain improved did not show a corresponding improvement in disability in two domains (ADL and mobility). Conclusions These findings highlight how, for some people, alleviating pain, the main symptom of arthritis, might not prevent the persistence or progression of disability. Even when pain improves, further interventions that improve disability are likely to be required. Statement of contribution What is already known on this subject? Pain and functional limitation in daily living are common symptoms of arthritis. Arthritis pain is heterogeneous \textendash{} there are trajectories of people whose pain gets better or worse. However, to date no study has looked at the relationship between trajectories of arthritis pain and functional disability outside of the minority of people with rheumatoid arthritis. What does this study add? Treating pain as heterogeneous explained disability better than treating pain as a single entity. Respondents in a trajectory of worsening pain reported functional disability in two domains (mobility and activities of daily living) also got worse over time. People in a trajectory of decreasing pain over time did not experience a reduction in disability, despite pain being the most common reason for why people limit their daily functioning. This suggests further intervention is required for people with arthritis, even when the most visible symptoms have been alleviated.},
  annotation = {\_eprint: https://bpspsychub.onlinelibrary.wiley.com/doi/pdf/10.1111/bjhp.12364},
  copyright  = {\textcopyright{} 2019 The Authors. British Journal of Health Psychology published by John Wiley \& Sons Ltd on behalf of British Psychological Society},
  doi        = {10.1111/bjhp.12364},
  file       = {C\:\\Users\\David\\Zotero\\storage\\BZ966GC7\\James et al. - 2019 - Trajectories of pain predict disabilities affectin.pdf},
  keywords   = {activities of daily living, arthritis, disability, pain, personalized medicine},
  language   = {en},
}

@Article{Ji2017,
  author     = {Ji, Hao and M{\"u}ller, Hans-Georg},
  journal    = {Journal of the Royal Statistical Society: Series B (Statistical Methodology)},
  title      = {Optimal Designs for Longitudinal and Functional Data},
  year       = {2017},
  issn       = {1467-9868},
  number     = {3},
  pages      = {859--876},
  volume     = {79},
  abstract   = {We propose novel optimal designs for longitudinal data for the common situation where the resources for longitudinal data collection are limited, by determining the optimal locations in time where measurements should be taken. As for all optimal designs, some prior information is needed to implement the optimal designs proposed. We demonstrate that this prior information may come from a pilot longitudinal study that has irregularly measured and noisy measurements, where for each subject one has available a small random number of repeated measurements that are randomly located on the domain. A second possibility of interest is that a pilot study consists of densely measured functional data and one intends to take only a few measurements at strategically placed locations in the domain for the future collection of similar data. We construct optimal designs by targeting two criteria: optimal designs to recover the unknown underlying smooth random trajectory for each subject from a few optimally placed measurements such that squared prediction errors are minimized; optimal designs that minimize prediction errors for functional linear regression with functional or longitudinal predictors and scalar responses, again from a few optimally placed measurements. The optimal designs proposed address the need for sparse data collection when planning longitudinal studies, by taking advantage of the close connections between longitudinal and functional data analysis. We demonstrate in simulations that the designs perform considerably better than randomly chosen design points and include a motivating data example from the Baltimore Longitudinal Study of Aging. The designs are shown to have an asymptotic optimality property.},
  annotation = {\_eprint: https://rss.onlinelibrary.wiley.com/doi/pdf/10.1111/rssb.12192},
  copyright  = {\textcopyright{} 2016 Royal Statistical Society},
  doi        = {10.1111/rssb.12192},
  file       = {C\:\\Users\\David\\Zotero\\storage\\3GPHCGUN\\Ji and Müller - 2017 - Optimal designs for longitudinal and functional da.pdf;C\:\\Users\\David\\Zotero\\storage\\RV8SQ7JH\\rssb.html},
  keywords   = {Asymptotics, Coefficient of determination, Functional data analysis, Functional principal components, Gaussian process, Karhunen–Loève expansion, Longitudinal data, Prediction error, Sparse design},
  language   = {en},
}

@article{Johnson2019,
  title = {Use of {{Mobile Health Apps}} and {{Wearable Technology}} to {{Assess Changes}} and {{Predict Pain}} during {{Treatment}} of {{Acute Pain}} in {{Sickle Cell Disease}}: {{Feasibility Study}}},
  author = {Johnson, Amanda and Yang, Fan and Gollarahalli, Siddharth and Banerjee, Tanvi and Abrams, Daniel and Jonassaint, Jude and Jonassaint, Charles and Shah, Nirmish},
  year = {2019},
  month = dec,
  volume = {7},
  pages = {e13671},
  publisher = {{JMIR Publications Inc.}},
  doi = {10.2196/13671},
  journal = {JMIR mHealth and uHealth},
  number = {12}
}

@article{Kannampallil2016,
  title = {Characterizing the Pain Score Trajectories of Hospitalized Adult Medical and Surgical Patients: A Retrospective Cohort Study},
  shorttitle = {Characterizing the Pain Score Trajectories of Hospitalized Adult Medical and Surgical Patients},
  author = {Kannampallil, Thomas and Galanter, William L. and Falck, Suzanne and Gaunt, Michael J. and Gibbons, Robert D. and McNutt, Robert and Odwazny, Richard and Schiff, Gordon and Vaida, Allen J. and Wilkie, Diana J. and Lambert, Bruce L.},
  year = {2016},
  month = dec,
  volume = {157},
  pages = {2739--2746},
  issn = {0304-3959},
  doi = {10.1097/j.pain.0000000000000693},
  abstract = {Pain care for hospitalized patients is often suboptimal. Representing pain scores as a graphical trajectory may provide insights into the understanding and treatment of pain. We describe a 1-year, retrospective, observational study to characterize pain trajectories of hospitalized adults during the first 48 hours after admission at an urban academic medical center. Using a subgroup of patients who presented with significant pain (pain score {$>$}4; n = 7762 encounters), we characterized pain trajectories and measured area under the curve, slope of the trajectory for the first 2 hours after admission, and pain intensity at plateau. We used mixed-effects regression to assess the association between pain score and sociodemographics (age, race, and gender), pain medication orders (opioids, nonopioids, and no medications), and medical service (obstetrics, psychiatry, surgery, sickle cell, intensive care unit, and medicine). K-means clustering was used to identify patient subgroups with similar trajectories. Trajectories showed differences based on race, gender, service, and initial pain score. Patients presumed to have dissimilar pain experiences (eg, sickle vs obstetrical) had markedly different pain trajectories. Patients with higher initial pain had a more rapid reduction during their first 2 hours of treatment. Pain reduction achieved in the 48 hours after admission was approximately 50\% of the initial pain, regardless of the initial pain. Most patients' pain failed to fully resolve, plateauing at a pain score of 4 or greater. Visualizing pain scores as graphical trajectories illustrates the dynamic variability in pain, highlighting pain responses over a period of observation, and may yield new insights for quality improvement and research.},
  file = {C\:\\Users\\David\\Zotero\\storage\\XF5XI9AL\\Kannampallil et al. - 2016 - Characterizing the pain score trajectories of hosp.pdf},
  journal = {PAIN},
  language = {en-US},
  number = {12}
}

@Article{Kersten2014,
  author     = {Kersten, Paula and White, Peter J. and Tennant, Alan},
  journal    = {PLOS ONE},
  title      = {Is the {{Pain Visual Analogue Scale Linear}} and {{Responsive}} to {{Change}}? {{An Exploration Using Rasch Analysis}}},
  year       = {2014},
  issn       = {1932-6203},
  month      = jun,
  number     = {6},
  pages      = {e99485},
  volume     = {9},
  abstract   = {Objectives Pain visual analogue scales (VAS) are commonly used in clinical trials and are often treated as an interval level scale without evidence that this is appropriate. This paper examines the internal construct validity and responsiveness of the pain VAS using Rasch analysis. Methods Patients (n = 221, mean age 67, 58\% female) with chronic stable joint pain (hip 40\% or knee 60\%) of mechanical origin waiting for joint replacement were included. Pain was scored on seven daily VASs. Rasch analysis was used to examine fit to the Rasch model. Responsiveness (Standardized Response Means, SRM) was examined on the raw ordinal data and the interval data generated from the Rasch analysis. Results Baseline pain VAS scores fitted the Rasch model, although 15 aberrant cases impacted on unidimensionality. There was some local dependency between items but this did not significantly affect the person estimates of pain. Daily pain (item difficulty) was stable, suggesting that single measures can be used. Overall, the SRMs derived from ordinal data overestimated the true responsiveness by 59\%. Changes over time at the lower and higher end of the scale were represented by large jumps in interval equivalent data points; in the middle of the scale the reverse was seen. Conclusions The pain VAS is a valid tool for measuring pain at one point in time. However, the pain VAS does not behave linearly and SRMs vary along the trait of pain. Consequently, Minimum Clinically Important Differences using raw data, or change scores in general, are invalid as these will either under- or overestimate true change; raw pain VAS data should not be used as a primary outcome measure or to inform parametric-based Randomised Controlled Trial power calculations in research studies; and Rasch analysis should be used to convert ordinal data to interval data prior to data interpretation.},
  doi        = {10.1371/journal.pone.0099485},
  file       = {C\:\\Users\\David\\Zotero\\storage\\VWNAZP89\\Kersten et al. - 2014 - Is the Pain Visual Analogue Scale Linear and Respo.pdf;C\:\\Users\\David\\Zotero\\storage\\RLEREFDL\\article.html},
  keywords   = {Acupuncture, Clinical trials, Hip, Knee joints, Measurement, Osteoarthritis, Pain, Randomized controlled trials},
  language   = {en},
  publisher  = {{Public Library of Science}},
  shorttitle = {Is the {{Pain Visual Analogue Scale Linear}} and {{Responsive}} to {{Change}}?},
}

@article{Laffont2014,
  title = {Multivariate Analysis of Longitudinal Ordinal Data with Mixed Effects Models, with Application to Clinical Outcomes in Osteoarthritis},
  author = {Laffont, Celine Marielle and Vandemeulebroecke, Marc and Concordet, Didier},
  year = {2014},
  month = may,
  volume = {109},
  pages = {955--966},
  publisher = {{Informa UK Limited}},
  doi = {10.1080/01621459.2014.917977},
  journal = {Journal of the American Statistical Association},
  number = {507}
}

@article{Lane2019,
  title = {Uncovering General, Shared, and Unique Temporal Patterns in Ambulatory Assessment Data.},
  author = {Lane, Stephanie T. and Gates, Kathleen M. and Pike, Hallie K. and Beltz, Adriene M. and Wright, Aidan G. C.},
  year = {2019},
  month = feb,
  volume = {24},
  pages = {54--69},
  publisher = {{American Psychological Association (APA)}},
  doi = {10.1037/met0000192},
  journal = {Psychological Methods},
  number = {1}
}

@article{Larsen1987,
  title = {The Stability of Mood Variability: {{A}} Spectral Analytic Approach to Daily Mood Assessments.},
  author = {Larsen, Randy J.},
  year = {1987},
  volume = {52},
  pages = {1195--1204},
  publisher = {{American Psychological Association (APA)}},
  doi = {10.1037/0022-3514.52.6.1195},
  journal = {Journal of Personality and Social Psychology},
  number = {6}
}

@article{Larsen1990,
  title = {Individual Differences in Entrainment of Mood to the Weekly Calendar.},
  author = {Larsen, Randy J. and Kasimatis, Margaret},
  year = {1990},
  volume = {58},
  pages = {164--171},
  publisher = {{American Psychological Association (APA)}},
  doi = {10.1037/0022-3514.58.1.164},
  journal = {Journal of Personality and Social Psychology},
  number = {1}
}

@Article{Lee2007,
  author     = {Lee, Keunbaik and Daniels, Michael J.},
  journal    = {Biometrics},
  title      = {A {{Class}} of {{Markov Models}} for {{Longitudinal Ordinal Data}}},
  year       = {2007},
  issn       = {1541-0420},
  number     = {4},
  pages      = {1060--1067},
  volume     = {63},
  abstract   = {Generalized linear models with serial dependence are often used for short longitudinal series. Heagerty (2002, Biometrics58, 342\textendash 351) has proposed marginalized transition models for the analysis of longitudinal binary data. In this article, we extend this work to accommodate longitudinal ordinal data. Fisher-scoring algorithms are developed for estimation. Methods are illustrated on quality-of-life data from a recent colorectal cancer clinical trial.},
  annotation = {\_eprint: https://onlinelibrary.wiley.com/doi/pdf/10.1111/j.1541-0420.2007.00800.x},
  doi        = {10.1111/j.1541-0420.2007.00800.x},
  file       = {C\:\\Users\\David\\Zotero\\storage\\IMHGJBST\\Lee and Daniels - 2007 - A Class of Markov Models for Longitudinal Ordinal .pdf},
  keywords   = {Fisher scoring, Generalized linear models, QOL},
  language   = {en},
}

@Article{Lee2008,
  author     = {Lee, Keunbaik and Daniels, Michael J.},
  journal    = {Statistics in Medicine},
  title      = {Marginalized Models for Longitudinal Ordinal Data with Application to Quality of Life Studies},
  year       = {2008},
  issn       = {1097-0258},
  number     = {21},
  pages      = {4359--4380},
  volume     = {27},
  abstract   = {Random effects are often used in generalized linear models to explain the serial dependence for longitudinal categorical data. Marginalized random effects models (MREMs) for the analysis of longitudinal binary data have been proposed to permit likelihood-based estimation of marginal regression parameters. In this paper, we propose a model to extend the MREM to accommodate longitudinal ordinal data. Maximum marginal likelihood estimation is proposed utilizing quasi-Newton algorithms with Monte Carlo integration of the random effects. Our approach is applied to analyze the quality of life data from a recent colorectal cancer clinical trial. Dropout occurs at a high rate and is often due to tumor progression or death. To deal with events due to progression/death, we used a mixture model for the joint distribution of longitudinal measures and progression/death times and use principal stratification to draw causal inferences about survivors. Copyright \textcopyright{} 2008 John Wiley \& Sons, Ltd.},
  annotation = {\_eprint: https://onlinelibrary.wiley.com/doi/pdf/10.1002/sim.3352},
  copyright  = {Copyright \textcopyright{} 2008 John Wiley \& Sons, Ltd.},
  doi        = {10.1002/sim.3352},
  file       = {C\:\\Users\\David\\Zotero\\storage\\Z92TLARC\\Lee and Daniels - 2008 - Marginalized models for longitudinal ordinal data .pdf;C\:\\Users\\David\\Zotero\\storage\\MQIPZ2SX\\sim.html},
  keywords   = {dropout, marginalized likelihood-based models, ordinal data models},
  language   = {en},
}

@Article{Lee2008a,
  author     = {Lee, Keunbaik and Daniels, Michael J.},
  journal    = {Statistics in Medicine},
  title      = {Marginalized Models for Longitudinal Ordinal Data with Application to Quality of Life Studies},
  year       = {2008},
  issn       = {1097-0258},
  number     = {21},
  pages      = {4359--4380},
  volume     = {27},
  abstract   = {Random effects are often used in generalized linear models to explain the serial dependence for longitudinal categorical data. Marginalized random effects models (MREMs) for the analysis of longitudinal binary data have been proposed to permit likelihood-based estimation of marginal regression parameters. In this paper, we propose a model to extend the MREM to accommodate longitudinal ordinal data. Maximum marginal likelihood estimation is proposed utilizing quasi-Newton algorithms with Monte Carlo integration of the random effects. Our approach is applied to analyze the quality of life data from a recent colorectal cancer clinical trial. Dropout occurs at a high rate and is often due to tumor progression or death. To deal with events due to progression/death, we used a mixture model for the joint distribution of longitudinal measures and progression/death times and use principal stratification to draw causal inferences about survivors. Copyright \textcopyright{} 2008 John Wiley \& Sons, Ltd.},
  annotation = {\_eprint: https://onlinelibrary.wiley.com/doi/pdf/10.1002/sim.3352},
  copyright  = {Copyright \textcopyright{} 2008 John Wiley \& Sons, Ltd.},
  doi        = {10.1002/sim.3352},
  file       = {C\:\\Users\\David\\Zotero\\storage\\9IAZUM75\\Lee and Daniels - 2008 - Marginalized models for longitudinal ordinal data .pdf;C\:\\Users\\David\\Zotero\\storage\\X6AQ3W7L\\sim.html},
  keywords   = {dropout, marginalized likelihood-based models, ordinal data models},
  language   = {en},
}

@Article{Liddell2018,
  author     = {Liddell, Torrin M. and Kruschke, John K.},
  journal    = {Journal of Experimental Social Psychology},
  title      = {Analyzing Ordinal Data with Metric Models: {{What}} Could Possibly Go Wrong?},
  year       = {2018},
  issn       = {0022-1031},
  month      = nov,
  pages      = {328--348},
  volume     = {79},
  abstract   = {We surveyed all articles in the Journal of Personality and Social Psychology (JPSP), Psychological Science (PS), and the Journal of Experimental Psychology: General (JEP:G) that mentioned the term ``Likert,'' and found that 100\% of the articles that analyzed ordinal data did so using a metric model. We present novel evidence that analyzing ordinal data as if they were metric can systematically lead to errors. We demonstrate false alarms (i.e., detecting an effect where none exists, Type I errors) and failures to detect effects (i.e., loss of power, Type II errors). We demonstrate systematic inversions of effects, for which treating ordinal data as metric indicates the opposite ordering of means than the true ordering of means. We show the same problems \textemdash{} false alarms, misses, and inversions \textemdash{} for interactions in factorial designs and for trend analyses in regression. We demonstrate that averaging across multiple ordinal measurements does not solve or even ameliorate these problems. A central contribution is a graphical explanation of how and when the misrepresentations occur. Moreover, we point out that there is no sure-fire way to detect these problems by treating the ordinal values as metric, and instead we advocate use of ordered-probit models (or similar) because they will better describe the data. Finally, although frequentist approaches to some ordered-probit models are available, we use Bayesian methods because of their flexibility in specifying models and their richness and accuracy in providing parameter estimates. An R script is provided for running an analysis that compares ordered-probit and metric models.},
  doi        = {10.1016/j.jesp.2018.08.009},
  file       = {C\:\\Users\\David\\Zotero\\storage\\T5DUK7IM\\Liddell and Kruschke - 2018 - Analyzing ordinal data with metric models What co.pdf;C\:\\Users\\David\\Zotero\\storage\\IYU6B3B6\\S0022103117307746.html},
  keywords   = {Bayesian analysis, Likert, Ordered-probit, Ordinal data},
  language   = {en},
  shorttitle = {Analyzing Ordinal Data with Metric Models},
}

@article{Lo2016,
  title = {Determining the Number of Factors in {{P}}-Technique Factor Analysis},
  author = {Lo, Lawrence L. and Molenaar, Peter C. M. and Rovine, Michael},
  year = {2016},
  month = may,
  volume = {21},
  pages = {94--105},
  publisher = {{Informa UK Limited}},
  doi = {10.1080/10888691.2016.1173549},
  journal = {Applied Developmental Science},
  number = {2}
}

@article{Lo2017,
  title = {Determining the Number of Factors in {{P}}-Technique Factor Analysis},
  author = {Lo, Lawrence L. and Molenaar, Peter C. M. and Rovine, Michael},
  year = {2017},
  month = apr,
  volume = {21},
  pages = {94--105},
  publisher = {{Routledge}},
  issn = {1088-8691},
  doi = {10.1080/10888691.2016.1173549},
  abstract = {Determining the number of factors is a critical first step in exploratory factor analysis. Although various criteria and methods for determining the number of factors have been evaluated in the usual between-subjects R-technique factor analysis, there is still question of how these methods perform in within-subjects P-technique factor analysis. A novel feature of P-technique data is that observations are usually sequentially dependent in some way. The current study evaluates 10 criteria for determining the number of factors for data simulated to resemble intensive repeated-measures data for which P-technique is usually applied. The number of factors, loading size, observed-to-latent ratio, serial-dependency, and sample size were simulation design factors with conditions simulated to resemble P-technique data. The methods are compiled from various statistical techniques and have been demonstrated in previous R-technique simulations. Finally, a real data example demonstrates applying the acceleration factor method to several time series data sets.},
  annotation = {\_eprint: https://doi.org/10.1080/10888691.2016.1173549},
  journal = {Applied Developmental Science},
  number = {2}
}

@book{McElreath2020,
  title = {Statistical Rethinking: A {{Bayesian}} Course with Examples in {{R}} and {{Stan}}},
  shorttitle = {Statistical Rethinking},
  author = {McElreath, Richard},
  year = {2020},
  edition = {Second},
  publisher = {{Taylor and Francis, CRC Press}},
  address = {{Boca Raton}},
  abstract = {"Statistical Rethinking: A Bayesian Course with Examples in R and Stan, Second Edition builds knowledge/confidence in statistical modeling. Pushes readers to perform step-by-step calculations (usually automated.) Unique, computational approach ensures readers understand details to make reasonable choices and interpretations in their modeling work"--},
  isbn = {978-0-367-13991-9},
  series = {{{CRC}} Texts in Statistical Science}
}

@article{Molenaar1985,
  title = {A Dynamic Factor Model for the Analysis of Multivariate Time Series},
  author = {Molenaar, Peter C. M.},
  year = {1985},
  month = jun,
  volume = {50},
  pages = {181--202},
  publisher = {{Springer Science and Business Media LLC}},
  doi = {10.1007/bf02294246},
  journal = {Psychometrika},
  number = {2}
}

@article{MOLENBERGHS1997,
  title = {The Analysis of Longitudinal Ordinal Data with Nonrandom Drop-Out},
  author = {MOLENBERGHS, G. and KENWARD, M. G. and LESAFFRE, E.},
  year = {1997},
  month = mar,
  volume = {84},
  pages = {33--44},
  issn = {0006-3444},
  doi = {10.1093/biomet/84.1.33},
  abstract = {A model is proposed for longitudinal ordinal data with nonrandom drop-out, which combines the multivariate Dale model for longitudinal ordinal data with a logistic regression model for dropout. Since response and drop-out are modelled as conditionally independent given complete data, the resulting likelihood can be maximised relatively simply, using the EM algorithm, which with acceleration is acceptably fast and, with appropriate additions, can produce estimates of precision. The approach is illustrated with an example. Such modelling of nonrandom drop-out requires caution because the interpretation of the fitted models depends on assumptions that are unexaminable in a fundamental sense, and the conclusions cannot be regarded as necessarily robust. The main role of such modelling may be as a component of a sensitivity analysis.},
  file = {C\:\\Users\\David\\Zotero\\storage\\6NEZEBMB\\MOLENBERGHS et al. - 1997 - The analysis of longitudinal ordinal data with non.pdf;C\:\\Users\\David\\Zotero\\storage\\CCVK3M2W\\243041.html},
  journal = {Biometrika},
  number = {1}
}

@article{Morris2015,
  title = {Functional Regression},
  author = {Morris, Jeffrey S.},
  year = {2015},
  month = apr,
  volume = {2},
  pages = {321--359},
  publisher = {{Annual Reviews}},
  doi = {10.1146/annurev-statistics-010814-020413},
  journal = {Annual Review of Statistics and Its Application},
  number = {1}
}

@article{Mun2019,
  title = {Investigating Intraindividual Pain Variability},
  author = {Mun, Chung Jung and Suk, Hye Won and Davis, Mary C. and Karoly, Paul and Finan, Patrick and Tennen, Howard and Jensen, Mark P.},
  year = {2019},
  month = nov,
  volume = {160},
  pages = {2415--2429},
  publisher = {{Ovid Technologies (Wolters Kluwer Health)}},
  doi = {10.1097/j.pain.0000000000001626},
  journal = {PAIN},
  number = {11}
}

@InCollection{NapaScollon2009,
  author     = {Napa Scollon, Christie and Prieto, Chu-Kim and Diener, Ed},
  booktitle  = {Assessing {{Well}}-{{Being}}: {{The Collected Works}} of {{Ed Diener}}},
  publisher  = {{Springer Netherlands}},
  title      = {Experience {{Sampling}}: {{Promises}} and {{Pitfalls}}, {{Strength}} and {{Weaknesses}}},
  year       = {2009},
  address    = {{Dordrecht}},
  editor     = {Diener, Ed},
  isbn       = {978-90-481-2354-4},
  pages      = {157--180},
  series     = {Social {{Indicators Research Series}}},
  abstract   = {The experience-sampling (ESM) technique is a method in which recording of feelings and activities is done on-line at the moment, either at randomly selected moments or at predetermined times. This method has the advantage of being able to not only assess people's general feelings, but to link feelings with situations, times of day, and other circumstances. Thus, ESM provides a powerful way of moving beyond simple questions about who is ``happy'' and who is not, to more intricate questions about when and why people experience positive and negative feelings. Compared to retrospective reports of feelings, ESM is less influenced by memory biases. ESM also allows researchers to analyze the patterning and relationships of feelings as they unfold over time. Despite the strengths and promise of this method, there are also limitations. For example, the heavy demand placed on research participants means that the sample might be biased toward highly conscientious individuals, and repeatedly reporting one's moods might itself influence feelings. How to analyze ESM data is discussed, including the issue of how to aggregate momentary feelings into global measures of the average subjective well-being of individuals.},
  doi        = {10.1007/978-90-481-2354-4_8},
  file       = {C\:\\Users\\David\\Zotero\\storage\\L4ITTIQ8\\Napa Scollon et al. - 2009 - Experience Sampling Promises and Pitfalls, Streng.pdf},
  keywords   = {Autobiographical Memory, Ecological Momentary Assessment, Experience Sampling, Experience Sampling Method, Memory Bias},
  language   = {en},
  shorttitle = {Experience {{Sampling}}},
}

@article{Nelson2011,
  title = {Dynamic {{P}}-{{Technique}} for {{Modeling Patterns}} of {{Data}}: {{Applications}} to {{Pediatric Psychology Research}}},
  author = {Nelson, T. D. and Aylward, B. S. and Rausch, J. R.},
  year = {2011},
  month = apr,
  volume = {36},
  pages = {959--968},
  publisher = {{Oxford University Press (OUP)}},
  doi = {10.1093/jpepsy/jsr023},
  journal = {Journal of Pediatric Psychology},
  number = {9}
}

@article{Nelson2011a,
  title = {Dynamic {{P}}-{{Technique}} for {{Modeling Patterns}} of {{Data}}: {{Applications}} to {{Pediatric Psychology Research}}},
  shorttitle = {Dynamic {{P}}-{{Technique}} for {{Modeling Patterns}} of {{Data}}},
  author = {Nelson, Timothy D. and Aylward, Brandon S. and Rausch, Joseph R.},
  year = {2011},
  month = oct,
  volume = {36},
  pages = {959--968},
  issn = {0146-8693},
  doi = {10.1093/jpepsy/jsr023},
  abstract = {Objective\hspace{1em}Dynamic p-technique (DPT) is a potentially useful statistical method for examining relationships among dynamic constructs in a single individual or small group of individuals over time. The purpose of this article is to offer a nontechnical introduction to DPT.\hspace{1em}Method\hspace{1em}An overview of DPT analysis, with an emphasis on potential applications to pediatric psychology research, is provided. To illustrate how DPT might be applied, an example using simulated data is presented for daily pain and negative mood ratings.\hspace{1em}Results\hspace{1em}The simulated example demonstrates the application of DPT to a relevant pediatric psychology research area. In addition, the potential application of DPT to the longitudinal study of adherence is presented.\hspace{1em}Conclusion\hspace{1em}Although it has not been utilized frequently within pediatric psychology, DPT could be particularly well-suited for research in this field because of its ability to powerfully model repeated observations from very small samples.},
  file = {C\:\\Users\\David\\Zotero\\storage\\QDQJ6VCX\\Nelson et al. - 2011 - Dynamic P-Technique for Modeling Patterns of Data.pdf},
  journal = {Journal of Pediatric Psychology},
  number = {9},
  pmcid = {PMC3173743},
  pmid = {21486938}
}

@article{Nesselroade2011,
  title = {Studying {{Intraindividual Variability}}: {{What We Have Learned That Will Help Us Understand Lives}} in {{Context}}},
  shorttitle = {Studying {{Intraindividual Variability}}},
  author = {Nesselroade, John R. and Ram, Nilam},
  year = {2011},
  month = jun,
  publisher = {{Lawrence Erlbaum Associates, Inc.}},
  issn = {1542-7609},
  abstract = {We examine some features of intraindividual variability and the research outcomes from its study. Selected current modeling techniques focused on individual-level analyses are briefly discussed, in...},
  copyright = {Copyright Taylor and Francis Group, LLC},
  journal = {Research in Human Development},
  language = {en}
}

@article{Nguyen2009,
  title = {{{WAVELET ESTIMATIONS FOR LONGITUDINAL DATA}}},
  author = {Nguyen, Trung Tu},
  year = {2009},
  volume = {19},
  pages = {667--684},
  publisher = {{Institute of Statistical Science, Academia Sinica}},
  issn = {1017-0405},
  abstract = {In the context of longitudinal data analysis, we study a random function X that represents patients or subjects observed at randomly distributed points. Principal components analysis (PCA) is useful in understanding the random effects of X. In this paper, we estimate the mean and covariance functions of X by wavelet methods. Proposed wavelet estimators give interesting performances over a wide class of functions, even if the regularity parameters of the original functions are not assumed to be greater than 2. In another problem of longitudinal analysis, we study the regression of observations at recorded times, when the number of observations per unit is assumed to be a finite integer. In this context, under Gaussian assumptions, our wavelet estimator could be proved to be optimal.},
  journal = {Statistica Sinica},
  number = {2}
}

@article{Pagani2012,
  title = {Longitudinal {{Rasch}} Model for Self Assessment of Side Effects in Chemotherapy Cycles},
  author = {Pagani, Laura and Zanarotti, M. C.},
  year = {2012},
  doi = {10.1285/i20705948v5n3p387},
  file = {C\:\\Users\\David\\Zotero\\storage\\WUIQ8EJ8\\53345071.html},
  language = {en-gb}
}

@article{Parry2019,
  title = {`{{Acute}} Flare-Ups' in Patients with, or at High Risk of, Knee Osteoarthritis: A Daily Diary Study with Case-Crossover Analysis},
  author = {Parry, E. and Ogollah, R. and Peat, G.},
  year = {2019},
  month = aug,
  volume = {27},
  pages = {1124--1128},
  publisher = {{Elsevier BV}},
  doi = {10.1016/j.joca.2019.04.003},
  journal = {Osteoarthritis and Cartilage},
  number = {8}
}

@inproceedings{Peladeau2018,
  title = {Comparison of {{Latent Dirichlet Modeling}} and {{Factor Analysis}} for {{Topic Extraction}}: {{A Lesson}} of {{History}}},
  booktitle = {Proceedings of the 51st {{Hawaii International Conference}} on {{System Sciences}}},
  author = {Peladeau, Normand and Davoodi, Elnaz},
  year = {2018},
  publisher = {{Hawaii International Conference on System Sciences}},
  doi = {10.24251/hicss.2018.078}
}

@article{Pesudovs2005,
  title = {Improving {{Subjective Scaling}} of {{Pain Using Rasch Analysis}}},
  author = {Pesudovs, Konrad and Noble, Bruce A.},
  year = {2005},
  month = sep,
  volume = {6},
  pages = {630--636},
  publisher = {{Elsevier}},
  issn = {1526-5900, 1528-8447},
  doi = {10.1016/j.jpain.2005.04.001},
  abstract = {{$<$}h2{$>$}Abstract{$<$}/h2{$><$}p{$>$}Pain management outcomes assessment depends on valid measurement of pain. However, the validity of single-item scales, such as numeric or faces scales, with the assignment of ordinal numerical values to response scale categories, is questionable. The universal assumption that equal distances between response choices represent equal distances on the dimension being measured is essentially erroneous. Herein we demonstrate that Rasch analysis can be used to expose and repair scale inequity and reengineer scale structure. Thirty-one subjects with severe ocular surface disease repeatedly completed a 7-category faces pain scale. Rasch analysis demonstrated that response category 5 was underutilized, leading to disordering of the response scale. Collapsing category 5 into either category 4 or 6 produced an ordered 6-category faces scale that could be recalibrated with average Rasch person measures to create linear measurement on a continuous latent variable. The value of further alterations to the scale was explored, and the implication for scale redesign discussed. Rasch analysis could be applied to any subjective pain measure post-hoc to create linear measurement or applied during instrument development to optimize design.{$<$}/p{$><$}h3{$>$}Perspective{$<$}/h3{$><$}p{$>$}Single-item scales like the faces scale or a 1\textendash 10 numerical rating scale are commonly used for the subjective assessment of pain. However, scores applied to response categories are arbitrary, so do not represent equidistant steps in the underlying latent variable (pain). Scale inequities are easily demonstrable and repairable with Rasch analysis.{$<$}/p{$>$}},
  file = {C\:\\Users\\David\\Zotero\\storage\\N44E5U32\\fulltext.html},
  journal = {The Journal of Pain},
  language = {English},
  number = {9},
  pmid = {16139782}
}

@article{Phillips2020,
  title = {Daily {{Physical Activity}} and {{Symptom Reporting}} in {{Breast Cancer Patients Undergoing Chemotherapy}}: {{An Intensive Longitudinal Examination}}},
  shorttitle = {Daily {{Physical Activity}} and {{Symptom Reporting}} in {{Breast Cancer Patients Undergoing Chemotherapy}}},
  author = {Phillips, Siobhan M. and Welch, Whitney A. and Fanning, Jason and {Santa-Maria}, Cesar A. and Gavin, Kara L. and {Auster-Gussman}, Lisa A. and Solk, Payton and Lu, Marilyn and Cullather, Erin and Khan, Seema A. and Kulkarni, Swati A. and Gradishar, William and Siddique, Juned},
  year = {2020},
  month = dec,
  volume = {29},
  pages = {2608--2616},
  publisher = {{American Association for Cancer Research}},
  issn = {1055-9965, 1538-7755},
  doi = {10.1158/1055-9965.EPI-20-0659},
  abstract = {Background: Increased activity is beneficial during chemotherapy, but treatment-related symptoms may be a barrier. This study examines the relationship between daily fluctuations in symptoms and activity during chemotherapy. Methods: Women undergoing chemotherapy for breast cancer [n = 67; Mage = 48.6 (SD = 10.3)] wore an accelerometer 24 hours/day and received four text prompts/day to rate symptoms for 10 consecutive days at the beginning, middle, and end of chemotherapy. Mixed-effects models were used to examine the between and within-person relationships between symptom ratings on a given day and moderate to vigorous physical activity (MVPA) and light physical activity (LPA) on that day and the following day controlling for relevant covariates and using the Bonferroni correction for multiple comparisons. Results: For MVPA and LPA, within-person associations were statistically significant for same day affect, fatigue, pain, walking, activities of daily living (ADL) physical function, and cognitive function. Previous day anxiety was associated with next day LPA. Every one point worse symptom rating than an individual's overall average was associated with: (i) between 1.49 (pain) and 4.94 (fatigue) minutes less MVPA and between 4.48 (pain) and 24.72 (ADL physical function) minutes less LPA that day, and (ii) 11.28 minutes less LPA the next day. No between-person effects were significant for MVPA or LPA. Conclusions: Daily within-person variations in symptoms were associated with MVPA and LPA during chemotherapy for breast cancer. Impact: Future work should explore relationships between symptoms and activity further and identify whether tailoring to symptoms enhances efficacy of physical activity promotion interventions during chemotherapy. Background: Increased activity is beneficial during chemotherapy, but treatment-related symptoms may be a barrier. This study examines the relationship between daily fluctuations in symptoms and activity during chemotherapy. Methods: Women undergoing chemotherapy for breast cancer [n = 67; Mage = 48.6 (SD = 10.3)] wore an accelerometer 24 hours/day and received four text prompts/day to rate symptoms for 10 consecutive days at the beginning, middle, and end of chemotherapy. Mixed-effects models were used to examine the between and within-person relationships between symptom ratings on a given day and moderate to vigorous physical activity (MVPA) and light physical activity (LPA) on that day and the following day controlling for relevant covariates and using the Bonferroni correction for multiple comparisons. Results: For MVPA and LPA, within-person associations were statistically significant for same day affect, fatigue, pain, walking, activities of daily living (ADL) physical function, and cognitive function. Previous day anxiety was associated with next day LPA. Every one point worse symptom rating than an individual's overall average was associated with: (i) between 1.49 (pain) and 4.94 (fatigue) minutes less MVPA and between 4.48 (pain) and 24.72 (ADL physical function) minutes less LPA that day, and (ii) 11.28 minutes less LPA the next day. No between-person effects were significant for MVPA or LPA. Conclusions: Daily within-person variations in symptoms were associated with MVPA and LPA during chemotherapy for breast cancer. Impact: Future work should explore relationships between symptoms and activity further and identify whether tailoring to symptoms enhances efficacy of physical activity promotion interventions during chemotherapy.},
  chapter = {Research Articles},
  copyright = {\textcopyright 2020 American Association for Cancer Research.},
  file = {C\:\\Users\\David\\Zotero\\storage\\XWF7AMBM\\2608.full-text.html},
  journal = {Cancer Epidemiology and Prevention Biomarkers},
  language = {en},
  number = {12},
  pmid = {32994340}
}

@article{Plan2012,
  title = {Likert Pain Score Modeling: A {{Markov}} Integer Model and an Autoregressive Continuous Model},
  author = {Plan, E. L. and Elshoff, J.-P. and Stockis, A. and {Sargentini-Maier}, M. L. and Karlsson, M. O.},
  year = {2012},
  month = mar,
  volume = {91},
  pages = {820--828},
  publisher = {{Springer Science and Business Media LLC}},
  doi = {10.1038/clpt.2011.301},
  journal = {Clinical Pharmacology \& Therapeutics},
  number = {5}
}

@phdthesis{Pope2019,
  title = {The Confluence of {{Gaussian}} Process Emulation and Wavelets},
  author = {Pope, Christopher Alexander},
  year = {2019},
  month = aug,
  abstract = {We discuss two thriving research areas, emulation (in the statistical sense) and wavelet analysis, and explore ways in which the two areas can complement each other to tackle problems that both areas face. The Gaussian process, which is the popular choice in emulation, is used due to its ability to be a surrogate for a function when we are only able to make a limited number of observations from the function. The Gaussian process, however, does not perform well when the underlying function contains a discontinuity. Wavelet analysis, on the other hand, is known for its ability to model and analyse functions that contain discontinuities. Wavelet analysis tends to require a large number of datapoints to be able to model functions accurately, tending to struggle when the amount of data is limited. As it appears that one area's strength is the other area's weakness, this thesis is aimed at exploring the possible overlaps between the two methods, and the ways in which they could benefit each other. Particular attention in the thesis is paid to the challenges that are faced when the function that we are attempting to model contains discontinuities, or, areas of space in which there is a sharp increase/decrease in the value of our observations. We develop methods to select the location of additional design points after we have observed the function at our original design points with the objective of better defining the location of the discontinuity. We also develop novel methods to model the unknown function that we believe contains discontinuities, and look to accurately find our uncertainty in this function.},
  copyright = {cc\_by\_nc\_sa},
  file = {C\:\\Users\\David\\Zotero\\storage\\RVKC9ZEB\\Pope - 2019 - The confluence of Gaussian process emulation and w.pdf;C\:\\Users\\David\\Zotero\\storage\\X4K5KGZ7\\25066.html},
  language = {en},
  school = {University of Leeds}
}

@article{Rahman2018,
  title = {Defining and {{Predicting Pain Volatility}} in {{Users}} of the {{Manage My Pain App}}: {{Analysis Using Data Mining}} and {{Machine Learning Methods}}},
  author = {Rahman, Quazi Abidur and Janmohamed, Tahir and Pirbaglou, Meysam and Clarke, Hance and Ritvo, Paul and Heffernan, Jane M. and Katz, Joel},
  year = {2018},
  month = nov,
  volume = {20},
  pages = {e12001},
  publisher = {{JMIR Publications Inc.}},
  doi = {10.2196/12001},
  journal = {Journal of Medical Internet Research},
  number = {11}
}

@article{Ram2005,
  title = {Examining {{Interindividual Differences}} in {{Cyclicity}} of {{Pleasant}} and {{Unpleasant Affects Using Spectral Analysis}} and {{Item Response Modeling}}},
  author = {Ram, Nilam and Chow, Sy-Miin and Bowles, Ryan P. and Wang, Lijuan and Grimm, Kevin and Fujita, Frank and Nesselroade, John R.},
  year = {2005},
  month = dec,
  volume = {70},
  pages = {773--790},
  publisher = {{Springer Science and Business Media LLC}},
  doi = {10.1007/s11336-001-1270-5},
  journal = {Psychometrika},
  number = {4}
}

@article{Ram2005a,
  title = {Examining {{Interindividual Differences}} in {{Cyclicity}} of {{Pleasant}} and {{Unpleasant Affects Using Spectral Analysis}} and {{Item Response Modeling}}},
  author = {Ram, Nilam and Chow, Sy-Miin and Bowles, Ryan P. and Wang, Lijuan and Grimm, Kevin and Fujita, Frank and Nesselroade, John R.},
  year = {2005},
  month = dec,
  volume = {70},
  pages = {773--790},
  publisher = {{Kluwer Academic Publishers-Plenum Publishers}},
  issn = {1860-0980},
  doi = {10.1007/s11336-001-1270-5},
  abstract = {Weekly cycles in emotion were examined by combining item response modeling and spectral analysis approaches in an analysis of 179 college students' reports of daily emotions experienced over 7 weeks. We addressed the measurement of emotion using an item response model. Spectral analysis and multilevel sinusoidal models were used to identify interindividual differences in intraindividual cyclic change. Simulations and incomplete data designs were used to examine how well this combination of analysis techniques might work when applied to other practical data problems. Empirically, we found systematic individual differences in the extent to which individuals' emotions follow a weekly cycle, and in how such cycles are exhibited. Weekly cycles accounted for very little variance in day to day emotions at the individual level. Analytically, we illustrate how measurement, change, and interindividual difference models from different traditions may be combined in a practical manner to describe some of the complexities of human behavior.},
  copyright = {2005 The Psychometric Society},
  file = {C\:\\Users\\David\\Zotero\\storage\\FUVQ5DZG\\Ram et al. - 2005 - Examining Interindividual Differences in Cyclicity.pdf;C\:\\Users\\David\\Zotero\\storage\\KT8ZICVX\\10.html},
  journal = {Psychometrika},
  language = {en},
  number = {4}
}

@book{Ram2013,
  title = {Dynamic {{Factor Analysis}}: {{Modeling Person}}-{{Specific Process}}},
  author = {Ram, Nilam and Brose, Annette and Molenaar, Peter C. M.},
  year = {2013},
  month = mar,
  publisher = {{Oxford University Press}},
  doi = {10.1093/oxfordhb/9780199934898.013.0021}
}

@misc{Ram2013a,
  title = {Dynamic {{Factor Analysis}}: {{Modeling Person}}-{{Specific Process}}},
  shorttitle = {Dynamic {{Factor Analysis}}},
  author = {Ram, Nilam and Brose, Annette and Molenaar, Peter C. M.},
  year = {2013},
  month = mar,
  doi = {10.1093/oxfordhb/9780199934898.013.0021},
  abstract = {"Dynamic Factor Analysis: Modeling Person-Specific Process" published on  by Oxford University Press.},
  howpublished = {https://www.oxfordhandbooks.com/view/10.1093/oxfordhb/9780199934898.001.0001/oxfordhb-9780199934898-e-021},
  isbn = {9780199934898},
  journal = {The Oxford Handbook of Quantitative Methods in Psychology: Vol. 2},
  language = {en}
}

@Book{Rasmussen2006,
  author     = {Rasmussen, Carl Edward and Williams, Christopher K. I.},
  publisher  = {{MIT Press}},
  title      = {Gaussian Processes for Machine Learning},
  year       = {2006},
  address    = {{Cambridge, Mass}},
  isbn       = {978-0-262-18253-9},
  series     = {Adaptive Computation and Machine Learning},
  annotation = {OCLC: ocm61285753},
  keywords   = {Data processing, Gaussian processes, Machine learning, Mathematical models},
  lccn       = {QA274.4 .R37 2006},
}

@article{Roberts2013,
  title = {Gaussian Processes for Time-Series Modelling},
  author = {Roberts, S. and Osborne, M. and Ebden, M. and Reece, S. and Gibson, N. and Aigrain, S.},
  year = {2013},
  month = feb,
  volume = {371},
  pages = {20110550},
  publisher = {{The Royal Society}},
  doi = {10.1098/rsta.2011.0550},
  journal = {Philosophical Transactions of the Royal Society A: Mathematical, Physical and Engineering Sciences},
  number = {1984}
}

@article{Roberts2013a,
  title = {Gaussian Processes for Time-Series Modelling},
  author = {Roberts, S. and Osborne, M. and Ebden, M. and Reece, S. and Gibson, N. and Aigrain, S.},
  year = {2013},
  month = feb,
  volume = {371},
  pages = {20110550},
  publisher = {{Royal Society}},
  doi = {10.1098/rsta.2011.0550},
  abstract = {In this paper, we offer a gentle introduction to Gaussian processes for time-series data analysis. The conceptual framework of Bayesian modelling for time-series data is discussed and the foundations of Bayesian non-parametric modelling presented for Gaussian processes. We discuss how domain knowledge influences design of the Gaussian process models and provide case examples to highlight the approaches.},
  file = {C\:\\Users\\David\\Zotero\\storage\\N5A8CSH9\\Roberts et al. - 2013 - Gaussian processes for time-series modelling.pdf},
  journal = {Philosophical Transactions of the Royal Society A: Mathematical, Physical and Engineering Sciences},
  number = {1984}
}

@inproceedings{Saatci2010,
  title = {Gaussian {{Process Change Point Models}}},
  booktitle = {{{ICML}}},
  author = {Saatci, Yunus and Turner, Ryan D. and Rasmussen, Carl Edward},
  year = {2010},
  month = jan,
  abstract = {We combine Bayesian online change point detection with Gaussian processes to create a nonparametric time series model which can handle change points. The model can be used to locate change points...},
  file = {C\:\\Users\\David\\Zotero\\storage\\2JHXJPMD\\Saatci et al. - 2010 - Gaussian Process Change Point Models.pdf;C\:\\Users\\David\\Zotero\\storage\\XGK4MQCS\\forum.html},
  language = {en}
}

@incollection{Scollon2009,
  title = {Experience {{Sampling}}: {{Promises}} and {{Pitfalls}}, {{Strength}} and {{Weaknesses}}},
  booktitle = {Assessing {{Well}}-{{Being}}},
  author = {Scollon, Christie Napa and Prieto, Chu-Kim and Diener, Ed},
  year = {2009},
  pages = {157--180},
  publisher = {{Springer Netherlands}},
  doi = {10.1007/978-90-481-2354-4_8}
}

@Misc{Stan2020,
  author = {{Stan Development Team}},
  note   = {R package version 2.21.2},
  title  = {{RStan}: the {R} interface to {Stan}},
  year   = {2020},
  url    = {http://mc-stan.org/},
}

@article{STOFFER1993,
  title = {Spectral Analysis for Categorical Time Series: {{Scaling}} and the Spectral Envelope},
  shorttitle = {Spectral Analysis for Categorical Time Series},
  author = {STOFFER, DAVID S. and TYLER, DAVID E. and MCDOUGALL, ANDREW J.},
  year = {1993},
  month = sep,
  volume = {80},
  pages = {611--622},
  issn = {0006-3444},
  doi = {10.1093/biomet/80.3.611},
  abstract = {Many studies produce categorical time series in which harmonic analysis is of interest. Although there exist time domain approaches for the analysis of categorical time series such as Markov chains or link function based regression models, there is apparently little statistical theory or methodology for analyzing qualitative-valued time series in the frequency domain. The purpose of this paper is to initiate the development of a general framework for the frequency domain analysis of categorical time series. In doing so, we discuss the scaling of categorical time series and introduce a new concept that we call the spectral envelope of a categorical time series. We demonstrate our methodology on a data set from a problem in molecular biology.},
  file = {C\:\\Users\\David\\Zotero\\storage\\EYXBCUR5\\STOFFER et al. - 1993 - Spectral analysis for categorical time series Sca.pdf},
  journal = {Biometrika},
  number = {3}
}

@article{Sun2020,
  title = {Using {{Smartphones}} and {{Wearable Devices}} to {{Monitor Behavioral Changes During COVID}}-19},
  author = {Sun, Shaoxiong and Folarin, Amos A. and Ranjan, Yatharth and Rashid, Zulqarnain and Conde, Pauline and Stewart, Callum and Cummins, Nicholas and Matcham, Faith and Costa, Gloria Dalla and Simblett, Sara and Leocani, Letizia and Lamers, Femke and S{\o}rensen, Per Soelberg and Buron, Mathias and Zabalza, Ana and P{\'e}rez, Ana Isabel Guerrero and Penninx, Brenda WJH and Siddi, Sara and Haro, Josep Maria and {Myin-Germeys}, Inez and Rintala, Aki and Wykes, Til and Narayan, Vaibhav A. and Comi, Giancarlo and Hotopf, Matthew and Dobson, Richard JB and Consortium, Radar-Cns},
  year = {2020},
  month = sep,
  volume = {22},
  pages = {e19992},
  publisher = {{JMIR Publications Inc., Toronto, Canada}},
  doi = {10.2196/19992},
  abstract = {Background: In the absence of a vaccine or effective treatment for COVID-19, countries have adopted nonpharmaceutical interventions (NPIs) such as social distancing and full lockdown. An objective and quantitative means of passively monitoring the impact and response of these interventions at a local level is needed. Objective: We aim to explore the utility of the recently developed open-source mobile health platform Remote Assessment of Disease and Relapse (RADAR)\textendash base as a toolbox to rapidly test the effect and response to NPIs intended to limit the spread of COVID-19. Methods: We analyzed data extracted from smartphone and wearable devices, and managed by the RADAR-base from 1062 participants recruited in Italy, Spain, Denmark, the United Kingdom, and the Netherlands. We derived nine features on a daily basis including time spent at home, maximum distance travelled from home, the maximum number of Bluetooth-enabled nearby devices (as a proxy for physical distancing), step count, average heart rate, sleep duration, bedtime, phone unlock duration, and social app use duration. We performed Kruskal-Wallis tests followed by post hoc Dunn tests to assess differences in these features among baseline, prelockdown, and during lockdown periods. We also studied behavioral differences by age, gender, BMI, and educational background. Results: We were able to quantify expected changes in time spent at home, distance travelled, and the number of nearby Bluetooth-enabled devices between prelockdown and during lockdown periods (P\&lt;.001 for all five countries). We saw reduced sociality as measured through mobility features and increased virtual sociality through phone use. People were more active on their phones (P\&lt;.001 for Italy, Spain, and the United Kingdom), spending more time using social media apps (P\&lt;.001 for Italy, Spain, the United Kingdom, and the Netherlands), particularly around major news events. Furthermore, participants had a lower heart rate (P\&lt;.001 for Italy and Spain; P=.02 for Denmark), went to bed later (P\&lt;.001 for Italy, Spain, the United Kingdom, and the Netherlands), and slept more (P\&lt;.001 for Italy, Spain, and the United Kingdom). We also found that young people had longer homestay than older people during the lockdown and fewer daily steps. Although there was no significant difference between the high and low BMI groups in time spent at home, the low BMI group walked more. Conclusions: RADAR-base, a freely deployable data collection platform leveraging data from wearables and mobile technologies, can be used to rapidly quantify and provide a holistic view of behavioral changes in response to public health interventions as a result of infectious outbreaks such as COVID-19. RADAR-base may be a viable approach to implementing an early warning system for passively assessing the local compliance to interventions in epidemics and pandemics, and could help countries ease out of lockdown.},
  file = {C\:\\Users\\David\\Zotero\\storage\\NGDTG4PZ\\Sun et al. - 2020 - Using Smartphones and Wearable Devices to Monitor .pdf},
  journal = {Journal of Medical Internet Research},
  language = {EN},
  number = {9}
}

@Article{Tan2020,
  author        = {Tan, Chang Wei and Bergmeir, Christoph and Petitjean, Francois and Webb, Geoffrey I.},
  title         = {Time Series Regression},
  year          = {2020},
  month         = jun,
  abstract      = {This paper introduces Time Series Regression (TSR): a little-studied task of which the aim is to learn the relationship between a time series and a continuous target variable. In contrast to time series classification (TSC), which predicts a categorical class label, TSR predicts a numerical value. This task generalizes forecasting, relaxing the requirement that the value predicted be a future value of the input series or primarily depend on more recent values. In this paper, we motivate and introduce this task, and benchmark possible solutions to tackling it on a novel archive of 19 TSR datasets which we have assembled. Our results show that the state-of-the-art TSC model Rocket, when adapted for regression, performs the best overall compared to other TSC models and state-of-the-art machine learning (ML) models such as XGBoost, Random Forest and Support Vector Regression.More importantly, we show that much research is needed in this field to improve the accuracy of ML models.},
  archiveprefix = {arXiv},
  eprint        = {2006.12672v1},
  eprinttype    = {arxiv},
  keywords      = {cs.LG, stat.ML},
}

@article{Tatar2018,
  title = {Development of a {{Self Report Stress Scale Using Item Response Theory}}-{{I}}: {{Item Selection}}, {{Formation}} of {{Factor Structure}} and {{Examination}} of {{Its Psychometric Properties}}},
  shorttitle = {Development of a {{Self Report Stress Scale Using Item Response Theory}}-{{I}}},
  author = {Tatar, Arkun and Saltuko{\u g}lu, Gaye and {\"O}zmen, Ercan},
  year = {2018},
  month = may,
  volume = {55},
  pages = {161--170},
  issn = {1300-0667},
  doi = {10.5152/npa.2017.18065},
  abstract = {Introduction: Due to the absence of Turkish psychometric devices assessing stress, in the present study it was aimed to develop a stress scale, and examine its basic psychometric properties. Methods: Current study included two processes, formation of item pool and examination of psychometric properties of the selected items through three studies. In the first study, 611 individuals aged between 18 and 77 responded to 130 selected items. In the second study, 2223 individuals aged between 18 and 68 responded to 80 items. In the third study, 1969 individuals aged between 18 and 79 responded to the final form of 36 items. Further, in study 3 for criterion related validity 163 individuals completed the Coopersmith Self-Esteem Inventory, 113 individuals completed the Beck Anxiety Inventory, 104 individuals completed the Hospital Anxiety and Depression Scale, 107 individuals completed the Beck Depression Inventory, and 265 individuals completed the Perceived Stress Scale. Moreover, in the investigation of test-retest reliability, 119 individuals took the final form of the test after 2 weeks, and 111 individuals took the final form of the test after 3 weeks. Results: In the first study, out of 130 items, 54 that showed item-total score correlations below 0.30 were excluded from the scale. Fifty-seven items were preserved exactly, and 19 items' sentence structures were changed. Furthermore, by adding 4 new items, 80 were prepared for the second study. In the second study, two factors structure namely ``Physiological Reactions/Strain'' and ``Psychological/Cognitive Appraisals'' sub-dimensions were identified, and 36 items were selected via Item Response Theory representing these sub-dimensions. In study 3, exploratory factor analysis provided strong support for our hypothesized two factors structure. Confirmatory factor analysis indicated hypothesized model had a better fit to the data. Internal consistency coefficients were 0.94 for the entire scale, 0.90 for Physiological Reactions/Strain sub-dimension, and 0.91 for Psychological/Cognitive Appraisals sub-dimension. Correlation coefficients between the entire scale and other criterion scales ranged from 0.22 to 0.63. Test-retest correlation coefficients between the first administration of the scale, and the administrations at two and three week intervals were 0.88. Conclusion: Results showed that the scale has basic psychometric requirements provided that the scale will be supported by validity studies.},
  file = {C\:\\Users\\David\\Zotero\\storage\\NBZ3ZYGJ\\Tatar et al. - 2018 - Development of a Self Report Stress Scale Using It.pdf},
  journal = {Archives of Neuropsychiatry},
  number = {2},
  pmcid = {PMC6060653},
  pmid = {30057459}
}

@Article{Timonen2020,
  author        = {Timonen, Juho and Mannerstr{\"o}m, Henrik and Vehtari, Aki and L{\"a}hdesm{\"a}ki, Harri},
  journal       = {arXiv:1912.03549 [cs, q-bio, stat]},
  title         = {Lgpr: {{An}} Interpretable Nonparametric Method for Inferring Covariate Effects from Longitudinal Data},
  year          = {2020},
  month         = oct,
  abstract      = {Longitudinal study designs are indispensable for studying disease progression. Inferring covariate effects from longitudinal data, however, requires interpretable methods that can model complicated covariance structures and detect nonlinear effects of both categorical and continuous covariates, as well as their interactions. Detecting disease effects is hindered by the fact that they often occur rapidly near the disease initiation time, and this time point cannot be exactly observed. An additional challenge is that the effect magnitude can be heterogeneous over the subjects. We present lgpr, a widely applicable and interpretable method for nonparametric analysis of longitudinal data using additive Gaussian processes. We demonstrate that it outperforms previous approaches in identifying the relevant categorical and continuous covariates in various settings. Furthermore, it implements important novel features, including the ability to account for the heterogeneity of covariate effects, their temporal uncertainty, and appropriate observation models for different types of biomedical data. The lgpr tool is implemented as a comprehensive and user-friendly R-package. lgpr is available at jtimonen.github.io/lgpr-usage with documentation, tutorials, test data, and code for reproducing the experiments of this paper.},
  archiveprefix = {arXiv},
  eprint        = {1912.03549},
  eprinttype    = {arxiv},
  file          = {C\:\\Users\\David\\Zotero\\storage\\M9B55P8Z\\Timonen et al. - 2020 - lgpr An interpretable nonparametric method for in.pdf;C\:\\Users\\David\\Zotero\\storage\\ZNZY3IXZ\\1912.html},
  keywords      = {Computer Science - Machine Learning, Quantitative Biology - Quantitative Methods, Statistics - Machine Learning, Statistics - Methodology},
  primaryclass  = {cs, q-bio, stat},
  shorttitle    = {Lgpr},
}

@article{Timonen2021,
  title = {Lgpr: An Interpretable Non-Parametric Method for Inferring Covariate Effects from Longitudinal Data},
  shorttitle = {Lgpr},
  author = {Timonen, Juho and Mannerstr{\"o}m, Henrik and Vehtari, Aki and L{\"a}hdesm{\"a}ki, Harri},
  year = {2021},
  month = jan,
  issn = {1367-4803},
  doi = {10.1093/bioinformatics/btab021},
  abstract = {Longitudinal study designs are indispensable for studying disease progression. Inferring covariate effects from longitudinal data, however, requires interpretable methods that can model complicated covariance structures and detect non-linear effects of both categorical and continuous covariates, as well as their interactions. Detecting disease effects is hindered by the fact that they often occur rapidly near the disease initiation time, and this time point cannot be exactly observed. An additional challenge is that the effect magnitude can be heterogeneous over the subjects.We present lgpr, a widely applicable and interpretable method for non-parametric analysis of longitudinal data using additive Gaussian processes. We demonstrate that it outperforms previous approaches in identifying the relevant categorical and continuous covariates in various settings. Furthermore, it implements important novel features, including the ability to account for the heterogeneity of covariate effects, their temporal uncertainty, and appropriate observation models for different types of biomedical data. The lgpr tool is implemented as a comprehensive and user-friendly R-package.lgpr is available at jtimonen.github.io/lgpr-usage with documentation, tutorials, test data and code for reproducing the experiments of this article.Supplementary data are available at Bioinformatics online.},
  file = {C\:\\Users\\David\\Zotero\\storage\\G32II4J3\\Timonen et al. - 2021 - lgpr an interpretable non-parametric method for i.pdf;C\:\\Users\\David\\Zotero\\storage\\DAQKZKNP\\6104850.html},
  journal = {Bioinformatics},
  number = {btab021}
}

@article{Torrence1998,
  title = {A {{Practical Guide}} to {{Wavelet Analysis}}},
  author = {Torrence, Christopher and Compo, Gilbert P},
  year = {1998},
  volume = {79},
  pages = {18},
  abstract = {A practical step-by-step guide to wavelet analysis is given, with examples taken from time series of the El Ni\~no\textendash Southern Oscillation (ENSO). The guide includes a comparison to the windowed Fourier transform, the choice of an appropriate wavelet basis function, edge effects due to finite-length time series, and the relationship between wavelet scale and Fourier frequency. New statistical significance tests for wavelet power spectra are developed by deriving theoretical wavelet spectra for white and red noise processes and using these to establish significance levels and confidence intervals. It is shown that smoothing in time or scale can be used to increase the confidence of the wavelet spectrum. Empirical formulas are given for the effect of smoothing on significance levels and confidence intervals. Extensions to wavelet analysis such as filtering, the power Hovm\"oller, cross-wavelet spectra, and coherence are described.},
  file = {C\:\\Users\\David\\Zotero\\storage\\K8UNTR9H\\Torrence and Compo - 1998 - A Practical Guide to Wavelet Analysis.pdf},
  journal = {Bulletin of the American Meteorological Society},
  language = {en},
  number = {1}
}

@article{Ullah2013,
  title = {Applications of Functional Data Analysis: A Systematic Review},
  author = {Ullah, Shahid and Finch, Caroline F.},
  year = {2013},
  month = mar,
  volume = {13},
  publisher = {{Springer Science and Business Media LLC}},
  doi = {10.1186/1471-2288-13-43},
  journal = {BMC Medical Research Methodology},
  number = {1}
}

@phdthesis{VanBerkel2019,
  title = {Data Quality and Quantity in Mobile Experience Sampling},
  author = {{van Berkel}, Niels},
  year = {2019},
  abstract = {The widespread availability of technologically-advanced mobile devices has brought researchers the opportunity to observe human life in day-to-day circumstances. Rather than studying human behaviour through extensive surveys or in artificial laboratory situations, this research instrument allows us to systematically capture human life in naturalistic settings. Mobile devices can capture two distinct data streams. First, the data from sensors embedded within these devices can be appropriated to construct the context of study participants. Second, participants can be asked to actively and repeatedly provide data on phenomena which cannot be reliably collected using the aforementioned sensor streams. This method is known as Experience Sampling. Researchers employing this method ask participants to provide observations multiple times per day, across a range of contexts, and to reflect on current rather than past experiences. This approach brings a number of advantages over existing methods, such as the ability to observe shifts in participant experiences over time and context, and reducing reliance on the participant's ability to accurately recall past events. As the onus of data collection lies with participants rather researchers, there is a firm reliance on the reliability of participant contributions. While previous work has focused on increasing the number of participant contributions, the quality of these contributions has remained relatively unexplored.    This thesis focuses on improving the quality and quantity of participant data collected through mobile Experience Sampling. Assessing and subsequently improving the quality of participant responses is a crucial step towards increasing the reliability of this increasingly popular data collection method. Previous recommendations for researchers are based primarily on anecdotal evidence or personal experience in running Experience Sampling studies. While such insights are valuable, it is challenging to replicate these recommendations and quantify their effect. Furthermore, we evaluate the application of this method in light of recent developments in mobile devices. The opportunities and challenges introduced by smartphone-based Experience Sampling studies remain underexplored in the current literature. Such devices can be utilised to infer participants' context and optimise questionnaire scheduling and presentation to increase data quality and quantity. By deploying our studies on these devices, we explore the opportunities of mobile sensing and interaction in the context of mobile Experience Sampling studies.    Our findings illustrate the feasibility of assessing and quantifying participant accuracy through the use of peer assessment, ground truth questions, and the assessment of cognitive skills. We empirically evaluate these approaches across a variety of study goals. Furthermore, our results provide recommendations on study design, motivation and data collection practices, and appropriate analysis techniques of participant data concerning response accuracy. Researchers can use our findings to increase the reliability of their data, to collect participant responses more evenly across different contexts in order to reduce the potential for bias, and to increase the total number of collected responses. The goal of this thesis is to improve the collection of human-labelled data in ESM studies, thereby strengthening the role of smartphones as valuable scientific instruments. Our work reveals a clear opportunity in the combination of human and sensor data sensing techniques for researchers interested in studying human behaviour in situ.},
  annotation = {Accepted: 2019-09-19T02:15:15Z},
  copyright = {Terms and Conditions: Copyright in works deposited in Minerva Access is retained by the copyright owner. The work may not be altered without permission from the copyright owner. Readers may only download, print and save electronic copies of whole works for their own personal non-commercial use. Any use that exceeds these limits requires permission from the copyright owner. Attribution is essential when quoting or paraphrasing from these works.},
  file = {C\:\\Users\\David\\Zotero\\storage\\H2UNLCGN\\van Berkel - 2019 - Data quality and quantity in mobile experience sam.pdf;C\:\\Users\\David\\Zotero\\storage\\S7E6FJAS\\227682.html},
  language = {en},
  school = {University of Melbourne}
}

@incollection{VanLear2015,
  title = {Time {{Series Analysis}} ({{ARIMA}}, {{Spectral Analysis}})},
  booktitle = {The {{International Encyclopedia}} of {{Interpersonal Communication}}},
  author = {VanLear, C. Arthur},
  editor = {Berger, Charles R. and Roloff, Michael E.},
  year = {2015},
  month = dec,
  pages = {1--5},
  publisher = {{Wiley}},
  doi = {10.1002/9781118540190.wbeic092}
}

@Article{VanSteen2001,
  author    = {{van Steen}, Kristel and Molenberghs, Geert and Verbeke, Geert and Thijs, Herbert},
  journal   = {Statistical Modelling},
  title     = {A Local Influence Approach to Sensitivity Analysis of Incomplete Longitudinal Ordinal Data},
  year      = {2001},
  issn      = {1471-082X},
  month     = jul,
  number    = {2},
  pages     = {125--142},
  volume    = {1},
  abstract  = {One of the major concerns when analysing incomplete longitudinal data is the fact that models necessarily rest on strong assumptions, unverifiable from the data. In response to these concerns, there is growing awareness of the usefulness of sensitivity analysis. In this paper we will focus on repeated ordinal data. Specifically, we implement a formal approach to such a sensitivity assessment, based on local influence, in the presence of multivariate categorical data. We explore the influence of perturbing a MAR dropout model in the direction of non-random dropout, and apply the proposed method to data from a longitudinal multicentre psychiatric study.},
  doi       = {10.1177/1471082X0100100203},
  keywords  = {incomplete data, influence analysis, missing data, multivariate Dale model, perturbation scheme, sensitivity analysis},
  language  = {en},
  publisher = {{SAGE Publications India}},
}

@article{Varin2009,
  title = {A Mixed Autoregressive Probit Model for Ordinal Longitudinal Data},
  author = {Varin, C. and Czado, C.},
  year = {2009},
  month = nov,
  volume = {11},
  pages = {127--138},
  publisher = {{Oxford University Press (OUP)}},
  doi = {10.1093/biostatistics/kxp042},
  journal = {Biostatistics},
  number = {1}
}

@article{Varin2010,
  title = {A Mixed Autoregressive Probit Model for Ordinal Longitudinal Data},
  author = {Varin, Cristiano and Czado, Claudia},
  year = {2010},
  month = jan,
  volume = {11},
  pages = {127--138},
  issn = {1465-4644},
  doi = {10.1093/biostatistics/kxp042},
  abstract = {Longitudinal data with binary and ordinal outcomes routinely appear in medical applications. Existing methods are typically designed to deal with short measurement series. In contrast, modern longitudinal data can result in large numbers of subject-specific serial observations. In this framework, we consider multivariate probit models with random effects to capture heterogeneity and autoregressive terms for describing the serial dependence. Since likelihood inference for the proposed class of models is computationally burdensome because of high-dimensional intractable integrals, a pseudolikelihood approach is followed. The methodology is motivated by the analysis of a large longitudinal study on the determinants of migraine severity.},
  file = {C\:\\Users\\David\\Zotero\\storage\\8LZZ599P\\Varin and Czado - 2010 - A mixed autoregressive probit model for ordinal lo.pdf},
  journal = {Biostatistics},
  number = {1}
}

@article{Verbeke1996,
  title = {A {{Linear Mixed}}-{{Effects Model}} with {{Heterogeneity}} in the {{Random}}-{{Effects Population}}},
  author = {Verbeke, Geert and Lesaffre, Emmanuel},
  year = {1996},
  month = mar,
  volume = {91},
  pages = {217--221},
  issn = {0162-1459, 1537-274X},
  doi = {10.1080/01621459.1996.10476679},
  journal = {Journal of the American Statistical Association},
  language = {en},
  number = {433}
}

@article{Vowels2018,
  title = {{{THE APPLICATION OF SPECTRAL AND CROSS}}-{{SPECTRAL ANALYSIS TO SOCIAL SCIENCES DATA}}},
  author = {Vowels, Matthew},
  year = {2018},
  month = jan,
  doi = {10.13023/ETD.2018.042},
  journal = {Theses and Dissertations--Family Sciences}
}

@Article{Vowels2020,
  author    = {Vowels, Matthew J. and Vowels, Laura M. and Wood, Nathan D.},
  title     = {Spectral and {{Cross}}-{{Spectral Analysis}} - a {{Tutorial}} for {{Psychologists}} and {{Social Scientists}}},
  year      = {2020},
  month     = sep,
  abstract  = {Social scientists have become increasingly interested in using intensive longitudinal methods to study social phenomena that change over time. Many of these phenomena are expected to exhibit cycling fluctuations (e.g., sleep, mood, sexual desire). However, researchers typically employ analytical methods which are unable to model such patterns. We present spectral and cross-spectral analysis as means to address this limitation. Spectral analysis provides a means to interrogate time series from a different, frequency domain perspective, and to understand how the time series may be decomposed into their constituent periodic components. Cross-spectral extends this to dyadic data and allows for synchrony and time offsets to be identified. The techniques are commonly used in the physical and engineering sciences, and we discuss how to apply these popular analytical techniques to the social sciences while also demonstrating how to undertake estimations of significance and effect size. In this tutorial we begin by introducing spectral and cross-spectral analysis, before demonstrating its application to simulated univariate and bivariate individual- and group-level data. We employ cross-power spectral density techniques to understand synchrony between the individual time series in a dyadic time series, and circular statistics and polar plots to understand phase offsets between constituent periodic components. Finally, we present a means to undertake non-parameteric boostrapping in order to estimate the significance, and derive a proxy for effect size. A Jupyter Notebook (Python 3.6) is provided as supplementary material to aid researchers who intend to apply these techniques.},
  doi       = {10.31234/osf.io/mj75a},
  file      = {C\:\\Users\\David\\Zotero\\storage\\89AJGMU8\\Vowels et al. - 2020 - Spectral and Cross-Spectral Analysis - a Tutorial .pdf},
  keywords  = {cross-spectral analysis, dyadic, Quantitative Methods, Social and Behavioral Sciences, spectral analysis, Statistical Methods, time series analysis},
  publisher = {{PsyArXiv}},
}

@article{Wang2016,
  title = {Functional Data Analysis},
  author = {Wang, Jane-Ling and Chiou, Jeng-Min and M{\"u}ller, Hans-Georg},
  year = {2016},
  month = jun,
  volume = {3},
  pages = {257--295},
  publisher = {{Annual Reviews}},
  doi = {10.1146/annurev-statistics-041715-033624},
  journal = {Annual Review of Statistics and Its Application},
  number = {1}
}

@article{Watson2020,
  title = {Latent {{Class Growth Analysis}} of {{Gout Flare Trajectories}}: {{A Three}}-{{Year Prospective Cohort Study}} in {{Primary Care}}},
  shorttitle = {Latent {{Class Growth Analysis}} of {{Gout Flare Trajectories}}},
  author = {Watson, Lorraine and Belcher, John and Nicholls, Elaine and Muller, Sara and Mallen, Christian and Roddy, Edward},
  year = {2020},
  volume = {72},
  pages = {1928--1935},
  issn = {2326-5205},
  doi = {10.1002/art.41476},
  abstract = {Objective To investigate the existence of distinct classes of gout flare trajectories and compare their gout-specific, comorbid, and sociodemographic characteristics. Methods In a prospective cohort study, adults with gout who were registered with 20 general practices self-reported the number of gout flares experienced at baseline and after 6, 12, 24, and 36 months via postal questionnaires. Latent class growth analysis (LCGA) was used to identify distinct gout flare trajectory classes. Statistical criteria and clinical interpretability were used to decide the optimal number of classes. Baseline comorbidities, medications, and sociodemographic and gout-specific characteristics of members of each class were described. Results A total of 1,164 participants (mean {$\pm$} SD age 65.6 {$\pm$} 12.5 years; 972 [84\%] male) were included. Six latent gout flare trajectory classes were identified: ``frequent and persistent'' (n = 95), ``gradually worsening'' (n = 276), ``frequent then improving'' (n = 14), ``moderately frequent'' (n = 287), ``moderately frequent then improving'' (n = 143), and ``infrequent'' (n = 349). The ``frequent and persistent'' trajectory had the most class members classified as obese and, along with the ``gradually worsening'' class, the highest proportion who were socioeconomically deprived. The ``frequent and persistent,'' ``gradually worsening,'' and ''frequent then improving'' classes had the highest proportions of class members with an estimated glomerular filtration rate {$<$}60 ml/minute/1.73 m2. The ``infrequent'' gout flare class was associated with more frequent allopurinol use and lower urate levels. Conclusion Six distinct gout flare trajectories were identified. Infrequent flares were associated with allopurinol use and lower serum urate levels, supporting the use of urate-lowering therapy to reduce flare frequency. The characteristics of flare trajectory classes could help to target interventions and improve patient care.},
  annotation = {\_eprint: https://onlinelibrary.wiley.com/doi/pdf/10.1002/art.41476},
  copyright = {\textcopyright{} 2020, American College of Rheumatology},
  file = {C\:\\Users\\David\\Zotero\\storage\\B9WWBQAV\\art.html},
  journal = {Arthritis \& Rheumatology},
  language = {en},
  number = {11}
}

@article{Wellhagen2019,
  title = {A {{Bounded Integer Model}} for {{Rating}} and {{Composite Scale Data}}},
  author = {Wellhagen, Gustaf J. and Kjellsson, Maria C. and Karlsson, Mats O.},
  year = {2019},
  month = jun,
  volume = {21},
  pages = {74},
  issn = {1550-7416},
  doi = {10.1208/s12248-019-0343-9},
  abstract = {Rating and composite scales are commonly used to assess treatment efficacy. The two main strategies for modelling such endpoints are to treat them as a continuous or an ordered categorical variable (CV or OC). Both strategies have disadvantages, including making assumptions that violate the integer nature of the data (CV) and requiring many parameters for scales with many response categories (OC). We present a method, called the bounded integer (BI) model, which utilises the probit function with fixed cut-offs to estimate the probability of a certain score through a latent variable. This method was successfully implemented to describe six data sets from four different therapeutic areas: Parkinson's disease, Alzheimer's disease, schizophrenia, and neuropathic pain. Five scales were investigated, ranging from 11 to 181 categories. The fit (likelihood) was better for the BI model than for corresponding OC or CV models ({$\Delta$}AIC range 11\textendash 1555) in all cases but one ({$\increment$}AIC -\,63), while the number of parameters was the same or lower. Markovian elements were successfully implemented within the method. The performance in external validation, assessed through cross-validation, was also in favour of the new model ({$\Delta$}OFV range 22\textendash 1694) except in one case ({$\increment$}OFV -\,70). A residual for diagnostic purposes is discussed. This study shows that the BI model respects the integer nature of data and is parsimonious in terms of number of estimated parameters.},
  file = {C\:\\Users\\David\\Zotero\\storage\\9378BPXP\\Wellhagen et al. - 2019 - A Bounded Integer Model for Rating and Composite S.pdf},
  journal = {The AAPS Journal},
  language = {en},
  number = {4}
}

@article{Wood1994,
  title = {The Study of Intraindividual Differences by Means of Dynamic Factor Models: {{Rationale}}, Implementation, and Interpretation.},
  author = {Wood, Phillip and Brown, David},
  year = {1994},
  volume = {116},
  pages = {166--186},
  publisher = {{American Psychological Association (APA)}},
  doi = {10.1037/0033-2909.116.1.166},
  journal = {Psychological Bulletin},
  number = {1}
}

@Article{Wood1994a,
  author     = {Wood, Phillip and Brown, David},
  journal    = {Psychological Bulletin},
  title      = {The Study of Intraindividual Differences by Means of Dynamic Factor Models: {{Rationale}}, Implementation, and Interpretation},
  year       = {1994},
  issn       = {1939-1455(Electronic),0033-2909(Print)},
  number     = {1},
  pages      = {166--186},
  volume     = {116},
  abstract   = {Study of reactive elements of behavior such as those responsible for mood disorder, arousal, and impulsivity often requires use of statistical models that involve time-bound relationships. P. C. Molenaar (see record 1985-27072-001) proposed the dynamic factor model for the analysis of multivariate repeated data. Conducting dynamic factor analyses using traditional structural equation programs is difficult because specification of such models involves extensive programming data manipulation and hand calculation to conduct such a time series analysis. This article outlines the dynamic factor model in terms of path diagrams and provides 3 SAS\textregistered{} macros that make specifying and conducting analyses easy. Problems of nonconvergence, improper solutions, and appropriate cutoff values for incremental fit indices are explored by means of a small Monte Carlo study. An example of physiological monitoring illustrates application of the model and use of the macros. (PsycINFO Database Record (c) 2016 APA, all rights reserved)},
  address    = {{US}},
  doi        = {10.1037/0033-2909.116.1.166},
  keywords   = {Factor Analysis, Individual Differences, Multivariate Analysis, Repeated Measures},
  publisher  = {{American Psychological Association}},
  shorttitle = {The Study of Intraindividual Differences by Means of Dynamic Factor Models},
}

@Article{Wu2020,
  author        = {Wu, Yue and Lyons, Terry J. and Saunders, Kate E. A.},
  journal       = {arXiv:2006.15030 [stat]},
  title         = {Deriving Information from Missing Data: Implications for Mood Prediction},
  year          = {2020},
  month         = jul,
  abstract      = {The availability of mobile technologies has enabled the efficient collection prospective longitudinal, ecologically valid self-reported mood data from psychiatric patients. These data streams have potential for improving the efficiency and accuracy of psychiatric diagnosis as well predicting future mood states enabling earlier intervention. However, missing responses are common in such datasets and there is little consensus as to how this should be dealt with in practice. A signature-based method was used to capture different elements of self-reported mood alongside missing data to both classify diagnostic group and predict future mood in patients with bipolar disorder, borderline personality disorder and healthy controls. The missing-response-incorporated signature-based method achieves roughly 66\textbackslash\% correct diagnosis, with f1 scores for three different clinic groups 59\textbackslash\% (bipolar disorder), 75\textbackslash\% (healthy control) and 61\textbackslash\% (borderline personality disorder) respectively. This was significantly more efficient than the naive model which excluded missing data. Accuracies of predicting subsequent mood states and scores were also improved by inclusion of missing responses. The signature method provided an effective approach to the analysis of prospectively collected mood data where missing data was common and should be considered as an approach in other similar datasets.},
  archiveprefix = {arXiv},
  eprint        = {2006.15030},
  eprinttype    = {arxiv},
  file          = {C\:\\Users\\David\\Zotero\\storage\\F5XAEJYI\\Wu et al. - 2020 - Deriving information from missing data implicatio.pdf;C\:\\Users\\David\\Zotero\\storage\\EHQF38T6\\2006.html},
  keywords      = {60L10; 60L90; 62D10; 92-08, Statistics - Applications},
  primaryclass  = {stat},
  shorttitle    = {Deriving Information from Missing Data},
}

@article{Zhu2019,
  title = {Clustering {{Functional Data}} with {{Application}} to {{Electronic Medication Adherence Monitoring}} in {{HIV Prevention Trials}}},
  author = {Zhu, Yifan and Di, Chongzhi and Chen, Ying Qing},
  year = {2019},
  month = jul,
  volume = {11},
  pages = {238--261},
  publisher = {{Springer US}},
  issn = {1867-1772},
  doi = {10.1007/s12561-019-09232-8},
  abstract = {Maintaining high medication adherence is essential for achieving desired efficacy in clinical trials, especially prevention trials. However, adherence is traditionally measured by self-reports that are subject to reporting biases and measurement error. Recently, electronic medication dispenser devices have been adopted in several HIV pre-exposure prophylaxis prevention studies. These devices are capable of collecting objective, frequent, and timely drug adherence data. The device opening signals generated by such devices are often represented as regularly or irregularly spaced discrete functional data, which are challenging for statistical analysis. In this paper, we focus on clustering the adherence monitoring data from such devices. We first pre-process the raw discrete functional data into smoothed functional data. Parametric mixture models with change-points, as well as several non-parametric and semi-parametric functional clustering approaches, are adapted and applied to the smoothed adherence data. Simulation studies were conducted to evaluate finite sample performances, on the choices of tuning parameters in the pre-processing step as well as the relative performance of different clustering algorithms. We applied these methods to the HIV Prevention Trials Network 069 study for identifying subgroups with distinct adherence behavior over the study period.},
  copyright = {2019 International Chinese Statistical Association},
  file = {C\:\\Users\\David\\Zotero\\storage\\P9Q7T5SC\\Zhu et al. - 2019 - Clustering Functional Data with Application to Ele.pdf},
  journal = {Statistics in Biosciences},
  language = {en},
  number = {2}
}

@book{zotero-109,
  type = {Book}
}

@misc{zotero-158,
  title = {Individual Differences in Entrainment of Mood to the Weekly Calendar. - {{PsycNET}}},
  file = {C\:\\Users\\David\\Zotero\\storage\\P6ZPJLVF\\doiLanding.html},
  howpublished = {https://doi.apa.org/doiLanding?doi=10.1037\%2F0022-3514.58.1.164}
}

@article{zotero-54,
  doi = {10.1080/01621459.1996.10476679}
}

@Article{Jahng2008,
  author    = {Seungmin Jahng and Phillip K. Wood and Timothy J. Trull},
  journal   = {Psychological Methods},
  title     = {Analysis of affective instability in ecological momentary assessment: Indices using successive difference and group comparison via multilevel modeling.},
  year      = {2008},
  month     = {dec},
  number    = {4},
  pages     = {354--375},
  volume    = {13},
  doi       = {10.1037/a0014173},
  publisher = {American Psychological Association ({APA})},
}

@InCollection{Vermunt2004,
  author    = {Jeroen K. Vermunt and Jacques A. Hagenaars},
  booktitle = {Methods in Human Growth Research},
  publisher = {Cambridge University Press},
  title     = {Ordinal longitudinal data analysis},
  year      = {2004},
  month     = {jun},
  pages     = {374--393},
  doi       = {10.1017/cbo9780511542411.016},
}

@Book{Agresti2002,
  author    = {Agresti, Alan},
  publisher = {Wiley},
  title     = {Categorical Data Analysis},
  year      = {2002},
  address   = {New York},
  isbn      = {0471360937},
}

@Article{druce:mcbeth:etal:2017,
  author    = {Katie L. Druce and John McBeth and Sabine N. van der Veer and David A. Selby and Bertie Vidgen and Konstantinos Georgatzis and Bruce Hellman and Rashmi Lakshminarayana and Afiqul Chowdhury and David M. Schultz and Caroline Sanders and Jamie C. Sergeant and William G. Dixon},
  journal   = {{JMIR} {mHealth} and {uHealth}},
  title     = {Recruitment and Ongoing Engagement in a {UK} Smartphone Study Examining the Association Between Weather and Pain: Cohort Study},
  year      = {2017},
  month     = {11},
  number    = {11},
  pages     = {e168},
  volume    = {5},
  doi       = {10.2196/mhealth.8162},
  publisher = {{JMIR} Publications Inc.},
}

@Article{Druce2019,
  author    = {Katie L. Druce and William G. Dixon and John McBeth},
  journal   = {Rheumatic Disease Clinics of North America},
  title     = {Maximizing Engagement in Mobile Health Studies},
  year      = {2019},
  month     = may,
  number    = {2},
  pages     = {159--172},
  volume    = {45},
  doi       = {10.1016/j.rdc.2019.01.004},
  publisher = {Elsevier {BV}},
}

@Article{Kahl2005,
  author    = {Cristiana Kahl and Joshua A. Cleland},
  journal   = {Physical Therapy Reviews},
  title     = {Visual analogue scale, numeric pain rating scale and the {McGill} pain Questionnaire: an overview of psychometric properties},
  year      = {2005},
  month     = {jun},
  number    = {2},
  pages     = {123--128},
  volume    = {10},
  doi       = {10.1179/108331905x55776},
  publisher = {Informa {UK} Limited},
}

@Article{Kimeldorf1970,
  author = {George S. Kimeldorf and Grace Wahba},
  title  = {A Correspondence Between {B}ayesian Estimation on Stochastic Processes and Smoothing by Splines},
  year   = {1970},
  issn   = {0003-4851},
  pages  = {495-502},
  volume = {41},
  doi    = {10.1214/aoms/1177697089},
}

@Article{Barbieri2017,
  author    = {Antoine Barbieri and Jean Peyhardi and Thierry Conroy and Sophie Gourgou and Christian Lavergne and Caroline Mollevi},
  journal   = {{BMC} Medical Research Methodology},
  title     = {Item response models for the longitudinal analysis of health-related quality of life in cancer clinical trials},
  year      = {2017},
  month     = {sep},
  number    = {1},
  volume    = {17},
  doi       = {10.1186/s12874-017-0410-9},
  publisher = {Springer Science and Business Media {LLC}},
}

@Article{Andrich1978,
  author    = {David Andrich},
  journal   = {Psychometrika},
  title     = {A rating formulation for ordered response categories},
  year      = {1978},
  month     = {dec},
  number    = {4},
  pages     = {561--573},
  volume    = {43},
  doi       = {10.1007/bf02293814},
  publisher = {Springer Science and Business Media {LLC}},
}

@Article{Buerkner2017,
  author    = {Paul-Christian Bürkner},
  journal   = {Journal of Statistical Software},
  title     = {brms: An R Package for Bayesian Multilevel Models Using Stan},
  year      = {2017},
  number    = {1},
  volume    = {80},
  doi       = {10.18637/jss.v080.i01},
  publisher = {Foundation for Open Access Statistic},
}

@Misc{Karch2016,
  author    = {Karch, Julian},
  title     = {A machine learning perspective on repeated measures},
  year      = {2016},
  copyright = {Namensnennung - Keine kommerzielle Nutzung - Keine Bearbeitung},
  doi       = {10.18452/17641},
  keywords  = {Längsschnittliche Datenanalyse, EEG-Analyse, Machine learning, Inter- and intraindividuelle Variation, Longitudinal data analysis, EEG analysis, Inter- and intraindividual variation, 150 Psychologie, 11 Psychologie, CM 3200},
  publisher = {Humboldt-Universität zu Berlin, Lebenswissenschaftliche Fakultät},
}

@Article{Karch2017,
  author    = {Julian Karch and Andreas Markus Brandmaier and Manuel Voelkle},
  title     = {Gaussian Process Panel Modeling {\textendash} Kernel-Based Longitudinal Modeling},
  year      = {2017},
  month     = {dec},
  doi       = {10.31234/osf.io/kvw5y},
  publisher = {Center for Open Science},
}

@Manual{R2021,
  title        = {R: A Language and Environment for Statistical Computing},
  address      = {Vienna, Austria},
  author       = {{R Core Team}},
  organization = {R Foundation for Statistical Computing},
  year         = {2021},
  url          = {https://www.R-project.org/},
}

@Article{Kosorok1996,
  author = {Michael R. Kosorok and Wei-Hsiung Chao},
  title  = {The Analysis of Longitudinal Ordinal Response Data in Continuous Time},
  year   = {1996},
  issn   = {0162-1459},
  pages  = {807-817},
  volume = {91},
  doi    = {10.1080/01621459.1996.10476948},
}

@Article{Ursino2016,
  author    = {Moreno Ursino and Mauro Gasparini},
  journal   = {Statistical Methods in Medical Research},
  title     = {A new parsimonious model for ordinal longitudinal data with application to subjective evaluations of a gastrointestinal disease},
  year      = {2016},
  month     = {aug},
  number    = {5},
  pages     = {1376--1393},
  volume    = {27},
  doi       = {10.1177/0962280216661370},
  publisher = {{SAGE} Publications},
}

@article{Heller2016,
  doi = {10.1016/j.sjpain.2016.06.012},
  url = {https://doi.org/10.1016/j.sjpain.2016.06.012},
  year = {2016},
  month = oct,
  publisher = {Walter de Gruyter {GmbH}},
  volume = {13},
  number = {1},
  pages = {67--75},
  author = {Gillian Z. Heller and Maurizio Manuguerra and Roberta Chow},
  title = {How to analyze the Visual Analogue Scale: Myths, truths and clinical relevance},
  journal = {Scandinavian Journal of Pain}
}

@Article{daSilva2016,
  author  = {Nívea B. {da Silva} and Leila D. A. F. Amorim and Enrico A. Colosimo and Leo Heller},
  journal = {Chilean Journal of Statistics},
  title   = {Modeling ordinal longitudinal outcomes: an applied perspective of marginal and conditional approaches},
  year    = {2016},
  issn    = {0718-7920},
  month   = sep,
  number  = {2},
  pages   = {51--68},
  volume  = {7},
  url     = {http://soche.cl/chjs/volumes/07/02/da_Silva_etal(2016).pdf},
}

@Article{RiutortMayol2020,
  author        = {Gabriel Riutort-Mayol and Paul-Christian Bürkner and Michael R. Andersen and Arno Solin and Aki Vehtari},
  title         = {Practical Hilbert space approximate Bayesian Gaussian processes for probabilistic programming},
  year          = {2020},
  month         = apr,
  abstract      = {Gaussian processes are powerful non-parametric probabilistic models for stochastic functions. However they entail a complexity that is computationally intractable when the number of observations is large, especially when estimated with fully Bayesian methods such as Markov chain Monte Carlo. In this paper, we focus on a novel approach for low-rank approximate Bayesian Gaussian processes, based on a basis function approximation via Laplace eigenfunctions for stationary covariance functions. The main contribution of this paper is a detailed analysis of the performance and practical implementation of the method in relation to key factors such as the number of basis functions, domain of the prediction space, and smoothness of the latent function. We provide intuitive visualizations and recommendations for choosing the values of these factors, which make it easier for users to improve approximation accuracy and computational performance. We also propose diagnostics for checking that the number of basis functions and the domain of the prediction space are adequate given the data. The proposed approach is simple and exhibits an attractive computational complexity due to its linear structure, and it is easy to implement in probabilistic programming frameworks. Several illustrative examples of the performance and applicability of the method in the probabilistic programming language Stan are presented together with the underlying Stan model code.},
  archiveprefix = {arXiv},
  eprint        = {2004.11408},
  file          = {:http\://arxiv.org/pdf/2004.11408v1:PDF},
  keywords      = {stat.CO, stat.ME},
  primaryclass  = {stat.CO},
}

@Article{Armstrong1989,
  author    = {Ben G. Armstrong and Margaret Sloan},
  journal   = {American Journal of Epidemiology},
  title     = {Ordinal regression models for epidemiologic data},
  year      = {1989},
  month     = jan,
  number    = {1},
  pages     = {191--204},
  volume    = {129},
  doi       = {10.1093/oxfordjournals.aje.a115109},
  publisher = {Oxford University Press},
}

@Article{Anderson1984,
  author    = {J. A. Anderson},
  journal   = {Journal of the Royal Statistical Society: Series B (Methodological)},
  title     = {Regression and Ordered Categorical Variables},
  year      = {1984},
  month     = {sep},
  number    = {1},
  pages     = {1--22},
  volume    = {46},
  doi       = {10.1111/j.2517-6161.1984.tb01270.x},
  publisher = {Wiley},
}

@Article{Ananth1997,
  author    = {C. V. Ananth and D. G. Kleinbaum},
  journal   = {International Journal of Epidemiology},
  title     = {Regression models for ordinal responses: a review of methods and applications},
  year      = {1997},
  month     = {dec},
  number    = {6},
  pages     = {1323--1333},
  volume    = {26},
  doi       = {10.1093/ije/26.6.1323},
  publisher = {Oxford University Press ({OUP})},
}

@Article{Liang1986,
  author    = {Kung-Yee Liang and Scott L. Zeger},
  journal   = {Biometrika},
  title     = {Longitudinal data analysis using generalized linear models},
  year      = {1986},
  number    = {1},
  pages     = {13--22},
  volume    = {73},
  doi       = {10.1093/biomet/73.1.13},
  publisher = {Oxford University Press ({OUP})},
}

@Article{Buerkner2019,
  author    = {Paul-Christian Bürkner and Matti Vuorre},
  journal   = {Advances in Methods and Practices in Psychological Science},
  title     = {Ordinal Regression Models in Psychology: A Tutorial},
  year      = {2019},
  month     = {feb},
  number    = {1},
  pages     = {77--101},
  volume    = {2},
  doi       = {10.1177/2515245918823199},
  publisher = {{SAGE} Publications},
}

@Article{Li2021,
  author    = {Fuxiao Li and Mengli Hao and Lijuan Yang},
  journal   = {{PLOS} {ONE}},
  title     = {Structural change detection in ordinal time series},
  year      = {2021},
  month     = {aug},
  number    = {8},
  pages     = {e0256128},
  volume    = {16},
  doi       = {10.1371/journal.pone.0256128},
  editor    = {Christophe Letellier},
  publisher = {Public Library of Science ({PLoS})},
}

@Article{Weiss2019,
  author    = {Christian H. Wei{\ss}},
  journal   = {Journal of the American Statistical Association},
  title     = {Distance-Based Analysis of Ordinal Data and Ordinal Time Series},
  year      = {2019},
  month     = {jun},
  number    = {531},
  pages     = {1189--1200},
  volume    = {115},
  doi       = {10.1080/01621459.2019.1604370},
  publisher = {Informa {UK} Limited},
}

@Article{Tan2012,
  author    = {Xianming Tan and Mariya P. Shiyko and Runze Li and Yuelin Li and Lisa Dierker},
  journal   = {Psychological Methods},
  title     = {A time-varying effect model for intensive longitudinal data.},
  year      = {2012},
  number    = {1},
  pages     = {61--77},
  volume    = {17},
  doi       = {10.1037/a0025814},
  publisher = {American Psychological Association ({APA})},
}

@Article{Ruppert2002,
  author    = {David Ruppert},
  journal   = {Journal of Computational and Graphical Statistics},
  title     = {Selecting the Number of Knots for Penalized Splines},
  year      = {2002},
  month     = {dec},
  number    = {4},
  pages     = {735--757},
  volume    = {11},
  doi       = {10.1198/106186002853},
  publisher = {Informa {UK} Limited},
}

@Article{Eilers2015,
  author  = {Eilers, Paul and Marx, Brian and Durbán, María},
  journal = {SORT (Statistics and Operations Research Transactions)},
  title   = {Twenty years of {P}-splines},
  year    = {2015},
  month   = {01},
  pages   = {149-186},
  volume  = {39},
  url     = {http://statweb.lsu.edu/faculty/marx/SORTTwentyYears.pdf},
}

@Article{Driver2018,
  author    = {Charles C. Driver and Manuel C. Voelkle},
  journal   = {Psychological Methods},
  title     = {Hierarchical Bayesian continuous time dynamic modeling.},
  year      = {2018},
  month     = {dec},
  number    = {4},
  pages     = {774--799},
  volume    = {23},
  doi       = {10.1037/met0000168},
  publisher = {American Psychological Association ({APA})},
}

@Book{Wood2017,
  author    = {Wood, Simon},
  publisher = {CRC Press/Taylor \& Francis Group},
  title     = {Generalized additive models: an introduction with {R}},
  year      = {2017},
  address   = {Boca Raton},
  isbn      = {9781498728331},
}

@Book{Hastie2009,
  author    = {Hastie, Trevor and Tibshirani, Robert and Friedman, Jerome},
  publisher = {Springer-Verlag GmbH},
  title     = {The Elements of Statistical Learning},
  year      = {2009},
  isbn      = {9780387848587},
  month     = aug,
  ean       = {9780387848587},
  pagetotal = {745},
  url       = {https://www.ebook.de/de/product/12472347/trevor_hastie_robert_tibshirani_jerome_friedman_the_elements_of_statistical_learning.html},
}

@Article{Perperoglou2019,
  author    = {Aris Perperoglou and Willi Sauerbrei and Michal Abrahamowicz and Matthias Schmid},
  journal   = {{BMC} Medical Research Methodology},
  title     = {A review of spline function procedures in {R}},
  year      = {2019},
  month     = {mar},
  number    = {1},
  volume    = {19},
  doi       = {10.1186/s12874-019-0666-3},
  publisher = {Springer Science and Business Media {LLC}},
}

@Article{Schultz2020,
  author    = {David M. Schultz and Anna L. Beukenhorst and Belay Birlie Yimer and Louise Cook and Huai Leng Pisaniello and Thomas House and Carolyn Gamble and Jamie C. Sergeant and John McBeth and William G. Dixon},
  journal   = {Bulletin of the American Meteorological Society},
  title     = {Weather Patterns Associated with Pain in Chronic-Pain Sufferers},
  year      = {2020},
  month     = {may},
  number    = {5},
  pages     = {E555--E566},
  volume    = {101},
  doi       = {10.1175/bams-d-19-0265.1},
  publisher = {American Meteorological Society},
}

@Book{Schloemilch1891,
  author    = {Schl{\"o}milch, Oskar Xaver and Witzschel, B and Cantor, Moritz and Kahl, Emil and Mehmke, Rudolf and Runge, Carl},
  publisher = {BG Teubner.},
  title     = {Zeitschrift f{\"u}r Mathematik und Physik},
  year      = {1891},
  volume    = {36},
}

@TechReport{CfE2022,
  author      = {{Centre for Epidemiology Versus Arthritis}},
  institution = {University of Manchester},
  title       = {The future of patient-generated data},
  year        = {2022},
  url         = {https://www.cfe.manchester.ac.uk/about/reports/},
}

@Article{Krieke2017,
  author    = {Lian van der Krieke and Frank J. Blaauw and Ando C. Emerencia and Hendrika M. Schenk and Joris P.J. Slaets and Elisabeth H. Bos and Peter de Jonge and Bertus F. Jeronimus},
  journal   = {Psychosomatic Medicine},
  title     = {Temporal Dynamics of Health and Well-Being: A Crowdsourcing Approach to Momentary Assessments and Automated Generation of Personalized Feedback},
  year      = {2017},
  month     = {feb},
  number    = {2},
  pages     = {213--223},
  volume    = {79},
  doi       = {10.1097/psy.0000000000000378},
  publisher = {Ovid Technologies (Wolters Kluwer Health)},
}

@Article{Bashir2018,
  author    = {Faraj Bashir and Hua-Liang Wei},
  journal   = {Neurocomputing},
  title     = {Handling missing data in multivariate time series using a vector autoregressive model-imputation ({VAR}-{IM}) algorithm},
  year      = {2018},
  month     = {feb},
  pages     = {23--30},
  volume    = {276},
  doi       = {10.1016/j.neucom.2017.03.097},
  publisher = {Elsevier {BV}},
}

@article{Wood2016,
  doi = {10.1080/01621459.2016.1180986},
  url = {https://doi.org/10.1080/01621459.2016.1180986},
  year = {2016},
  month = oct,
  publisher = {Informa {UK} Limited},
  volume = {111},
  number = {516},
  pages = {1548--1563},
  author = {Simon N. Wood and Natalya Pya and Benjamin S\"{a}fken},
  title = {Smoothing Parameter and Model Selection for General Smooth Models},
  journal = {Journal of the American Statistical Association}
}

@Comment{jabref-meta: databaseType:bibtex;}
