## Supplementary material for "Describing variability of intensively collected longitudinal ordinal data with latent spline models": R code for simulations + LaTeX source files: sparsity-results-table.tex

\begin{table}
\label{tab:sparsity-sim}
\caption{Simulation results comparing true vs. estimated MADE for $n=100$ simulations per sample size.}
\centering
\begin{tabular}[t]{rrrrr}
\toprule
Sample Size & Avg. True \textsc{Made} & Avg. Estimated \textsc{Made} & Avg. Bias & Avg. Error (\%)\\
\midrule
20 & 0.410 & 0.273 & --0.137 & --33.406\\
40 & 0.412 & 0.345 & --0.068 & --16.424\\
60 & 0.408 & 0.352 & --0.055 & --13.601\\
80 & 0.435 & 0.394 & --0.041 & --9.342\\
100 & 0.407 & 0.384 & --0.023 & --5.697\\
\bottomrule
\end{tabular}
\end{table}
