## Supplementary material for "Describing variability of intensively collected longitudinal ordinal data with latent spline models": R code for simulations + LaTeX source files: sparsity_simulation.R

```
# ---
# Supplementary Simulation Study: The Effect of Data Sparsity on MADE
# ---
# This script assesses how the number of available observations per subject
# influences the accuracy of the Mean Absolute Deviation from Expectation (MADE)
# statistic derived from a latent ordinal spline model.

# 1. SETUP: LOAD LIBRARIES AND DEFINE PARAMETERS
# -------------------------------------------------

# Required packages for modelling and data manipulation
library(mgcv)
library(dplyr)
library(knitr) # For nice table formatting

# --- Simulation Parameters ---
set.seed(42) # For reproducibility
n_sims <- 50 # Number of simulations per scenario
sample_sizes <- c(20, 40, 60, 80, 100) # Scenarios for data sparsity
n_ord <- 5 # Number of ordinal levels for the pain scale

# --- Model Parameters ---
# Cutpoints for the 5-level ordinal scale on the latent (normal) scale
cutpoints <- qnorm(seq(n_ord - 1) / n_ord)

# True latent curve for "Subject H" from the original manuscript simulation
# A sinusoidal pattern over 100 days
days <- 1:100
wx_h <- 2 * sin(seq(-pi, pi, length.out = length(days)))
# Standard deviation of the noise for Subject H
sd_h <- 0.7

# 2. HELPER FUNCTIONS
# -------------------------------------------------

#' Calculate the median predicted level from a gam model's response probabilities
#' @param model A fitted gam object with family = ocat()
#' @return A vector of predicted median ordinal levels.
get_median_prediction <- function(model) {
  # Get the matrix of response probabilities (rows=observations, cols=levels)
  probs <- predict(model, type = 'response')
  # For each row, find the first level where the cumulative probability exceeds 0.5
  apply(probs, 1, function(p) which.max(cumsum(p) > 0.5))
}


# 3. THE SIMULATION FUNCTION
# -------------------------------------------------

#' Runs a single simulation for a given sample size.
#' @param sample_size The number of observations to sample from the full 100 days.
#' @return A tibble with the true_made and the estimated_made for one simulation run.
run_one_simulation <- function(sample_size) {
  #message(sample_size)
  # --- a) Generate a single "full" dataset (n=100) from the true process ---
  full_data <- tibble(
    day = days,
    wx = wx_h,
    epsilon = rnorm(length(days), mean = 0, sd = sd_h),
    latent = wx + epsilon,
    y = 1L + findInterval(latent, cutpoints) #ordered(1 + findInterval(latent, cutpoints))
  )

  # --- b) Calculate the "True MADE" for this realization ---
  # The "true" prediction is the median level derived from the noiseless latent curve 'wx'.
  true_pred_median <- 1 + findInterval(full_data$wx, cutpoints)
  true_made <- mean(abs(as.numeric(full_data$y) - true_pred_median))

  # --- c) Create the sparse dataset by subsampling ---
  sparse_data <- full_data %>% sample_n(sample_size)

  # --- d) Fit the GAM model to the sparse data ---
  # Using penalized splines (ps) and the ordered categorical (ocat) family.
  # k=10 is a reasonable basis size for up to 100 days, allowing penalization
  # to control the smoothness.
  # browser()
  fit <- gam(
    y ~ s(day, bs = 'ps', k = 10),
    data = sparse_data,
    family = ocat(R = n_ord)
  )

  # --- e) Calculate the "Estimated MADE" from the fitted model ---
  # This is the MADE calculated using the model fitted to the sparse data.
  est_pred_median <- get_median_prediction(fit)
  est_made <- mean(abs(as.numeric(sparse_data$y) - est_pred_median))

  # --- f) Return results for this single run ---
  return(tibble(true_made = true_made, est_made = est_made))
}

# Sometimes mgcv will fail. Handle these errors gracefully.
try_one_simulation <- purrr::safely(
  run_one_simulation,
  otherwise = tibble(true_made = NA, est_made = NA)
)


# 4. RUN THE FULL SIMULATION
# -------------------------------------------------

# Create a data frame that pairs each sample size with the number of simulations
simulation_plan <- tibble(
  sample_size = rep(sample_sizes, each = n_sims)
)

# Run the simulation for each row in the plan
# This will take a minute or two to complete.
message("Running simulations... This may take a moment.")
simulation_results <- simulation_plan %>%
  mutate(
    results = purrr::map(sample_size, try_one_simulation, .progress = TRUE) |>
      purrr::transpose() |>
      purrr::pluck('result')
  ) %>%
  tidyr::unnest(cols = c(results))
message("Simulations complete.\n\n")


# 5. SUMMARIZE AND PRESENT RESULTS
# -------------------------------------------------

summary_table <- simulation_results %>%
  group_by(sample_size) %>%
  summarise(
    # Average the "true" MADE across all simulations for a stable baseline
    mean_true_made = mean(true_made, na.rm = TRUE),
    # Calculate the average estimated MADE from the models
    mean_est_made = mean(est_made, na.rm = TRUE),
    # Calculate the average bias (difference between estimated and true)
    bias = mean(est_made - true_made, na.rm = TRUE),
    .groups = 'drop'
  ) %>%
  mutate(
    # Express the bias as a percentage of the true value
    percent_error = (bias / mean_true_made) * 100
  )

# Print the final summary table in a clean format
cat("--- Simulation Summary ---\n")
cat("Performance of MADE estimation under varying data sparsity\n\n")

kable(
  summary_table,
  'latex',
  booktabs = TRUE,
  digits = 3,
  col.names = c(
    "Sample Size",
    "Avg. True MADE",
    "Avg. Estimated MADE",
    "Avg. Bias",
    "Avg. Error (%)"
  ),
  caption = "Simulation results comparing true vs. estimated MADE for $n=100$ simulations per sample size."
)

# --- Optional: Visualization ---
# A boxplot can show the distribution of the estimated MADE for each scenario
library(ggplot2)

ggplot(simulation_results, aes(x = factor(sample_size), y = est_made)) +
  geom_boxplot(fill = "steelblue", alpha = 0.7) +
  geom_hline(
    data = summary_table,
    aes(yintercept = mean_true_made),
    linetype = "dashed",
    color = "red"
  ) +
  facet_wrap(~sample_size, scales = "free_x", nrow = 1) +
  labs(
    title = "Distribution of Estimated MADE by Sample Size",
    subtitle = "Red dashed line indicates the average 'True MADE'",
    x = "Sample Size per Subject",
    y = "Estimated MADE"
  ) +
  theme_minimal() +
  theme(strip.text = element_blank()) # Hide facet labels as x-axis is sufficient
```
