## Supplementary figures and images for "Describing variability of intensively collected longitudinal ordinal data with latent spline models"

### fig_mode1.png

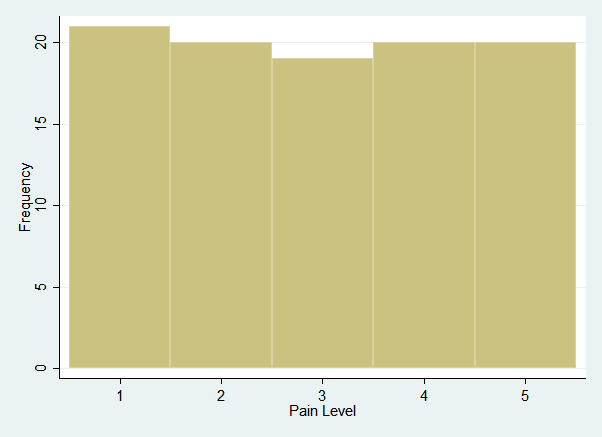

### fig_mode5.png

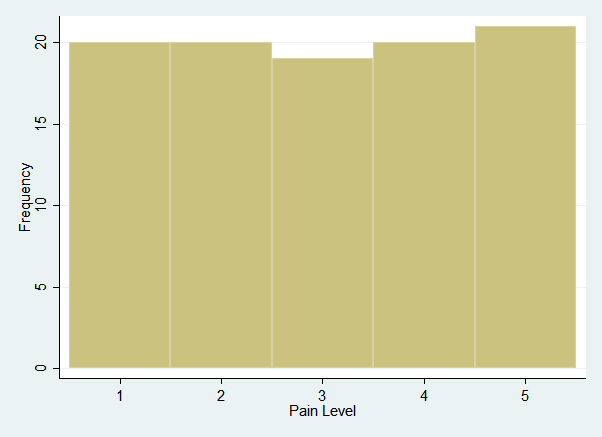

### heatplot_2.png

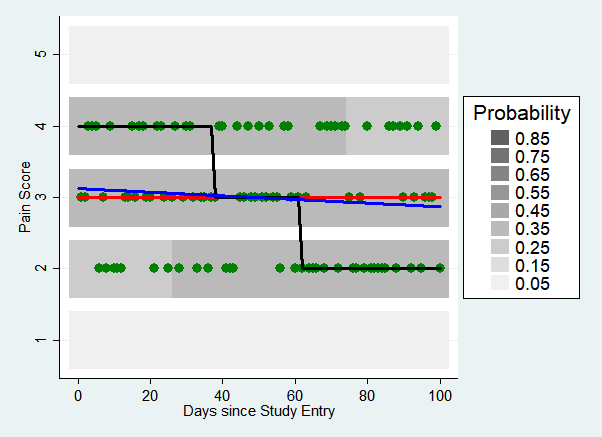

### heatplot_2_bw.png

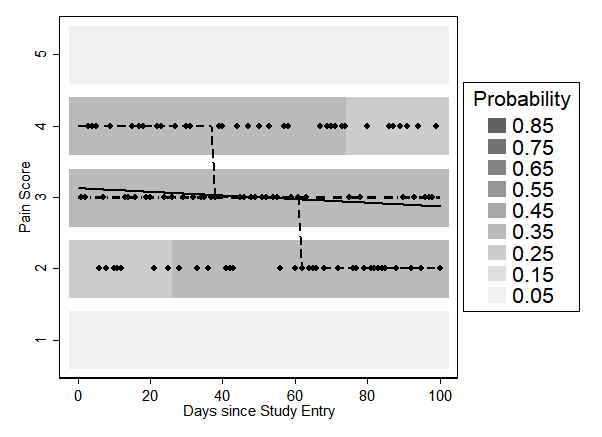

### heatplot_3.png

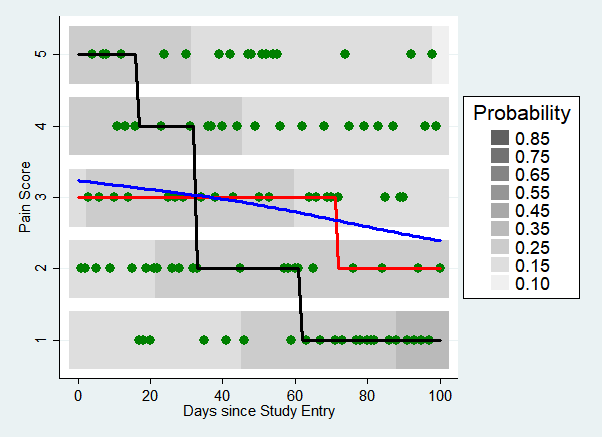

### heatplot_3_bw.png

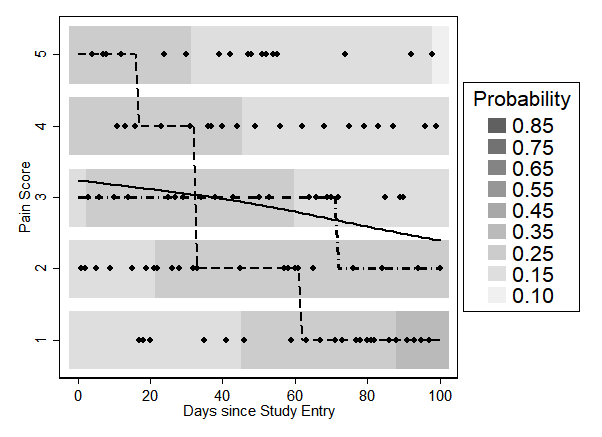

### heatplot_4.png

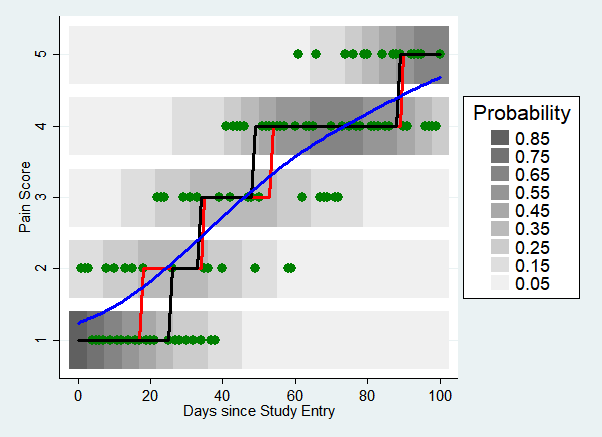

### heatplot_4_bw.png

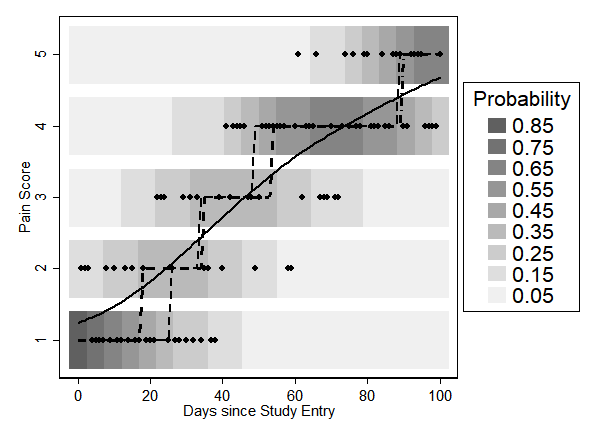

### heatplot_5.png

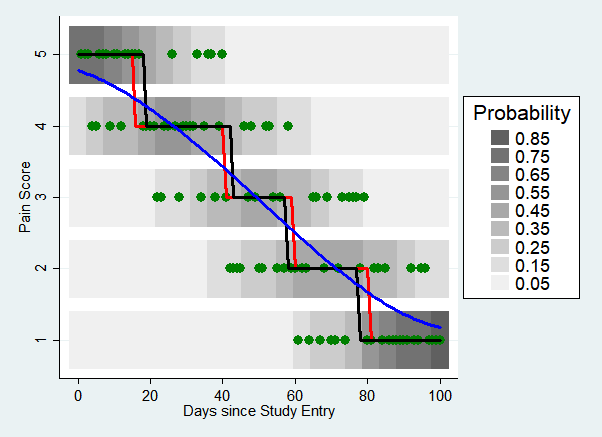

### heatplot_5_bw.png

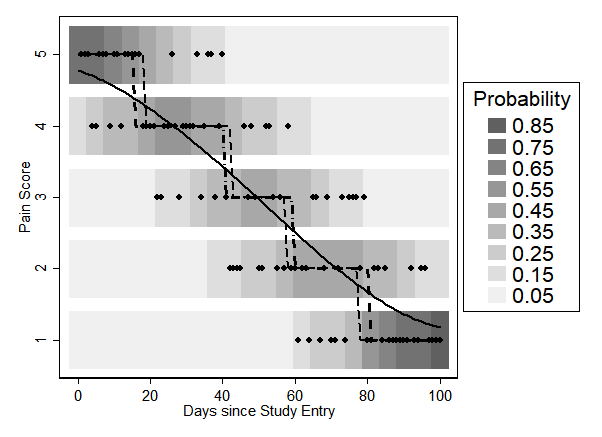

### heatplot_6.png

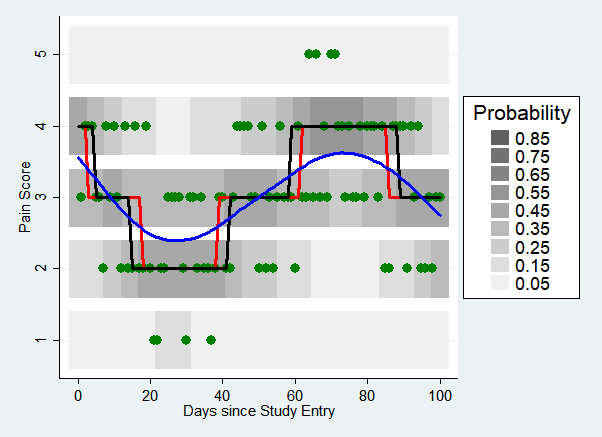

### heatplot_6_bw.png

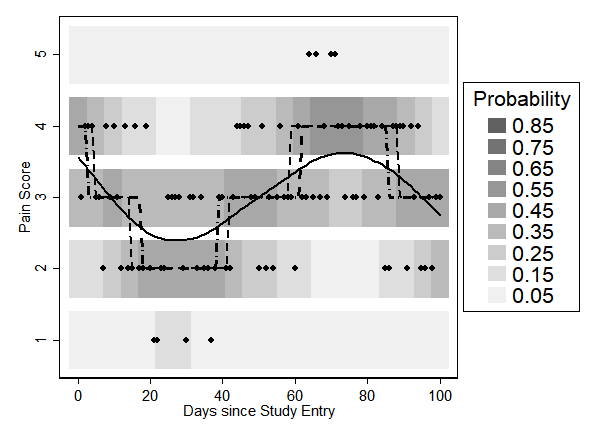

### heatplot_7.png

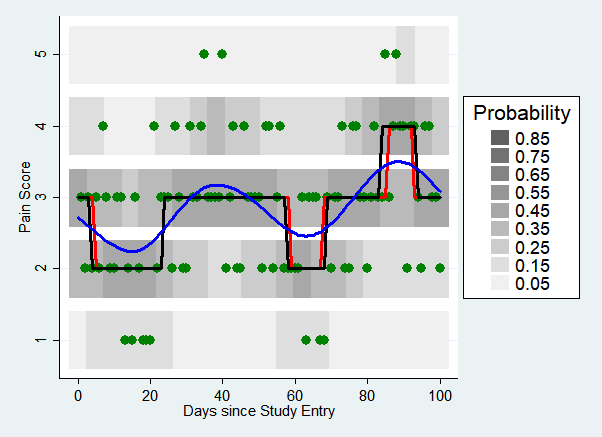

### heatplot_7_bw.png

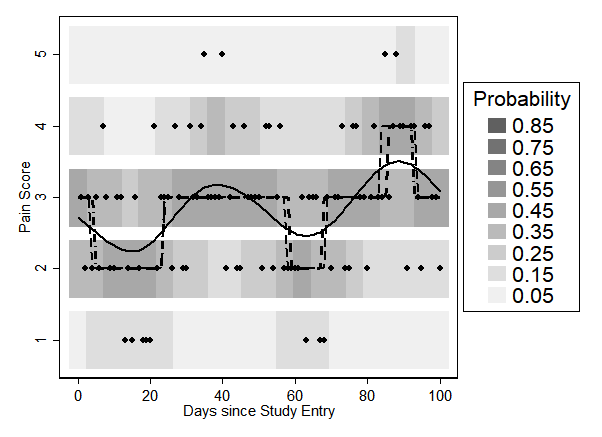

### heatplot_8.png

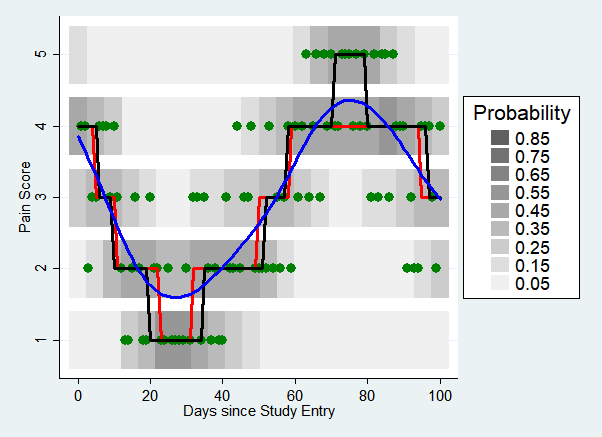

### heatplot_8_bw.png

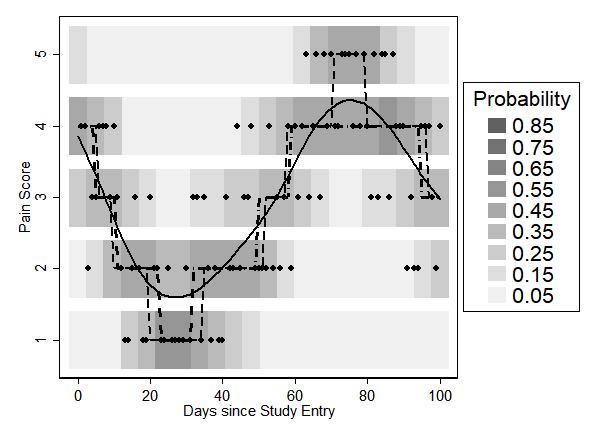

### legend-eps-converted-to.pdf

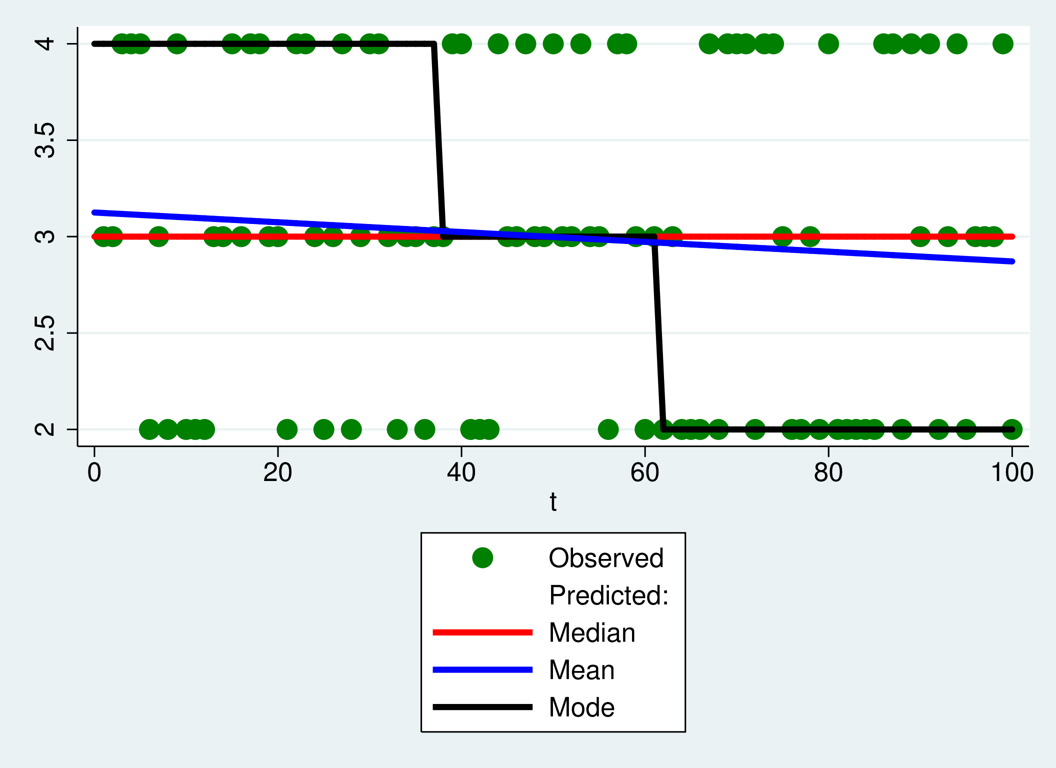

### lin_plot_1.png

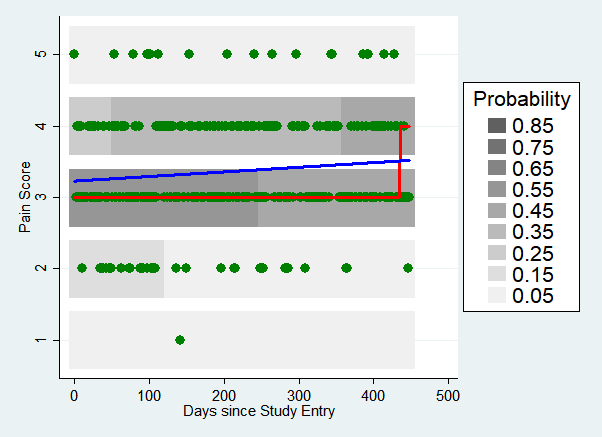

### lin_plot_1_bw.png

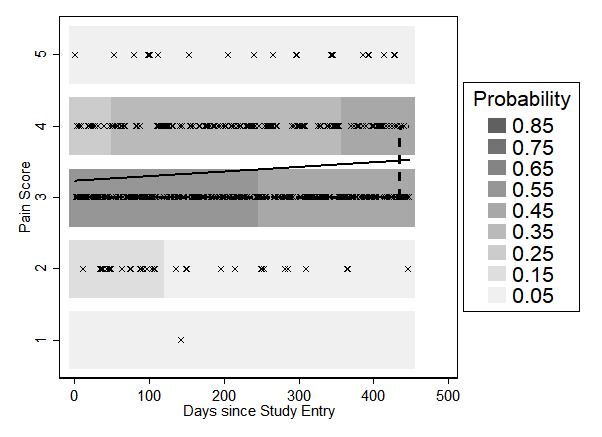

### lin_plot_2.png

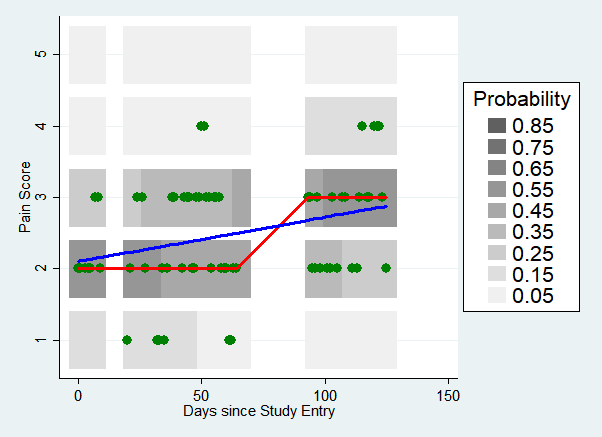

### lin_plot_2_bw.png

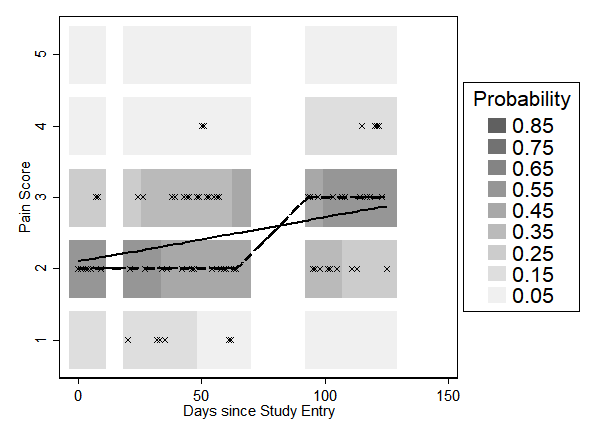

### lin_plot_3.png

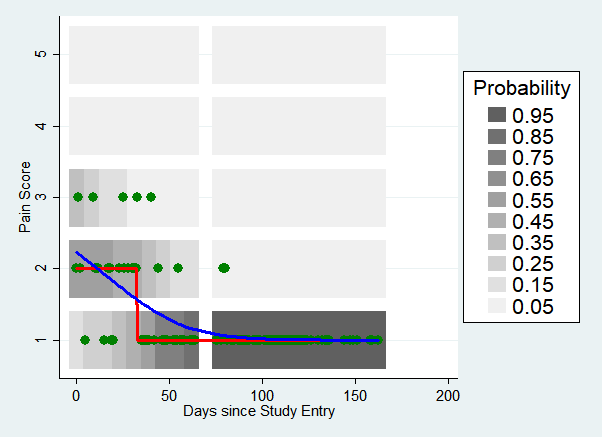

### lin_plot_3_bw.png

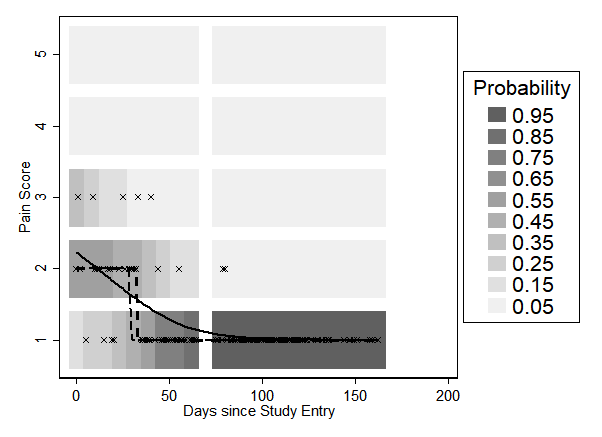

### nlin_plot_1.pdf

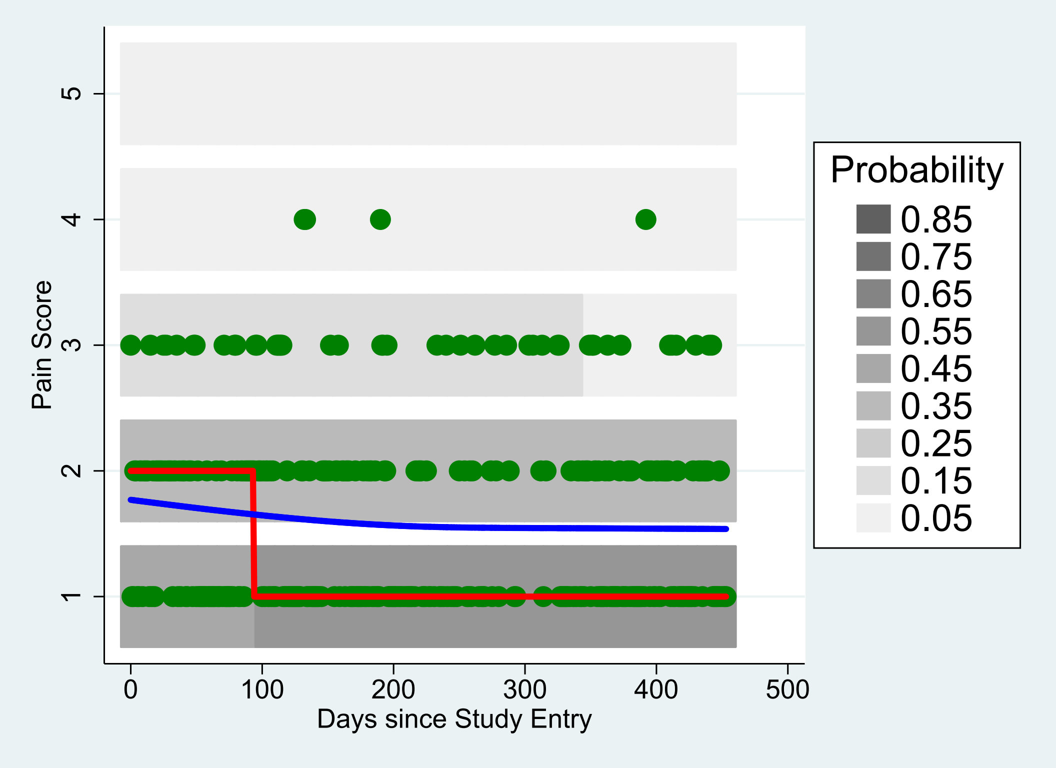

### nlin_plot_1.png

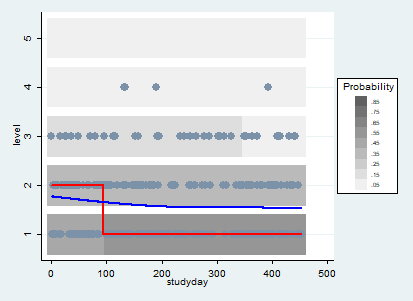

### nlin_plot_1_bw.png

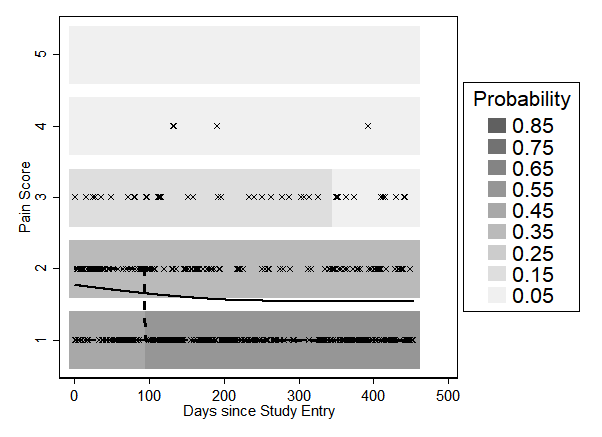

### nlin_plot_2.pdf

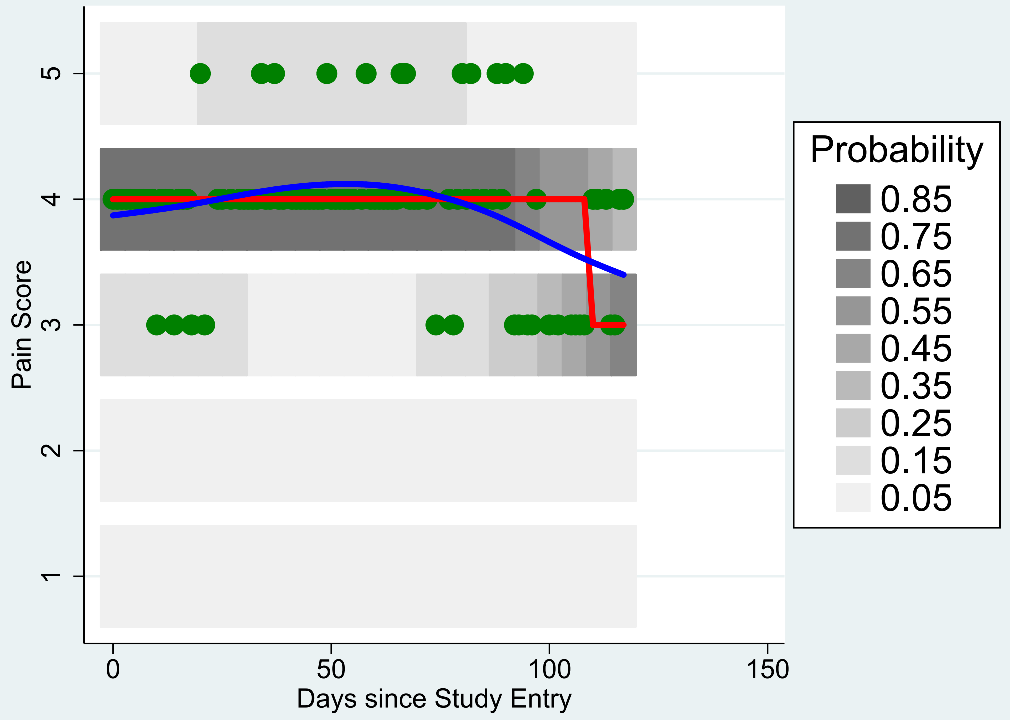

### nlin_plot_2.png

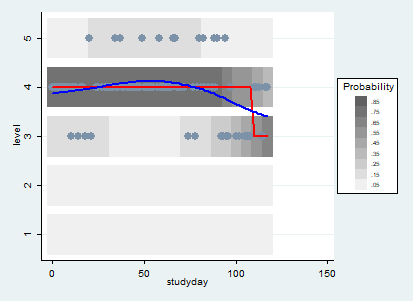

### nlin_plot_2_bw.png

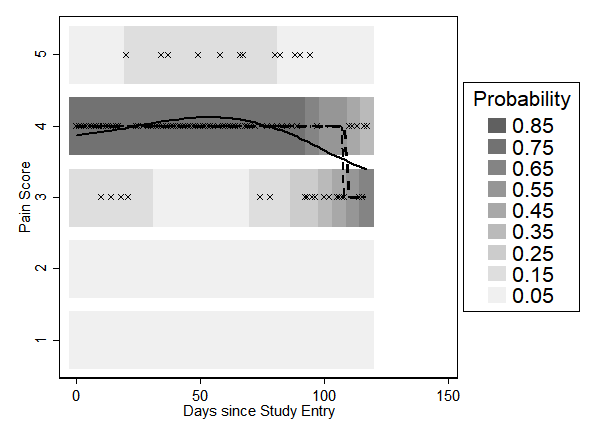

### nlin_plot_3.pdf

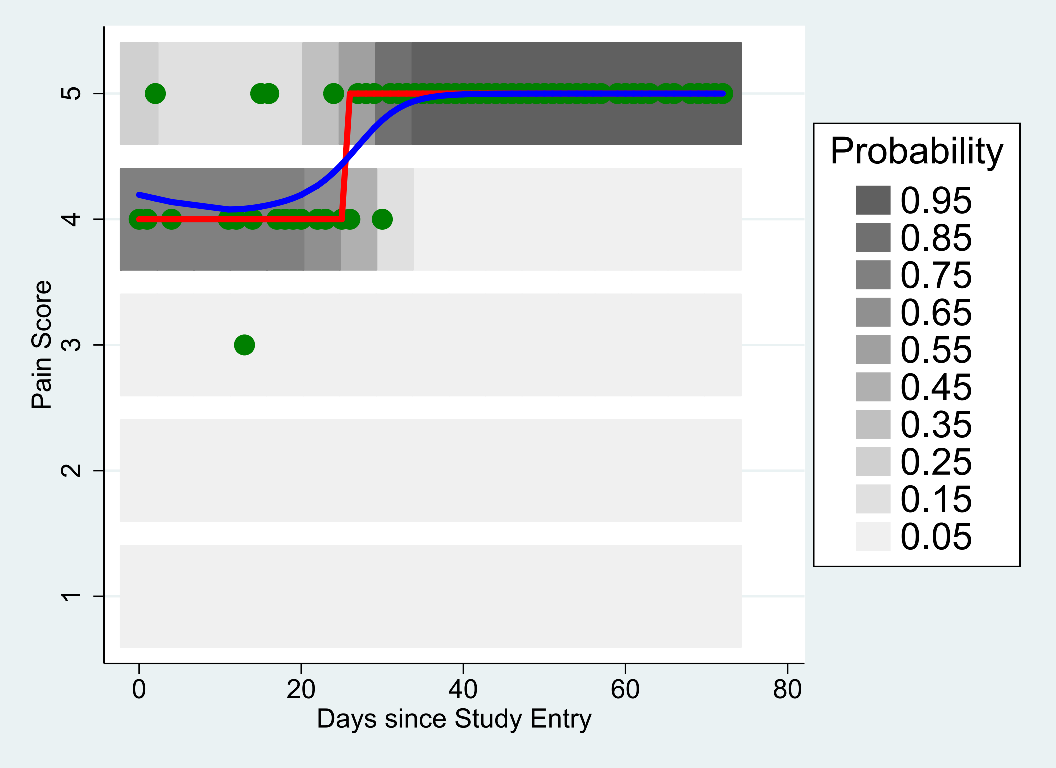

### nlin_plot_3.png

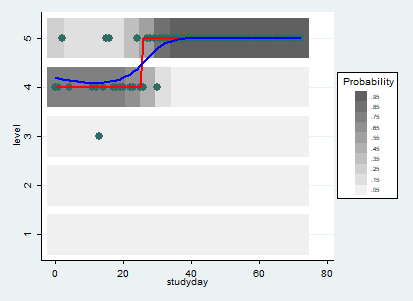

### nlin_plot_3_bw.png

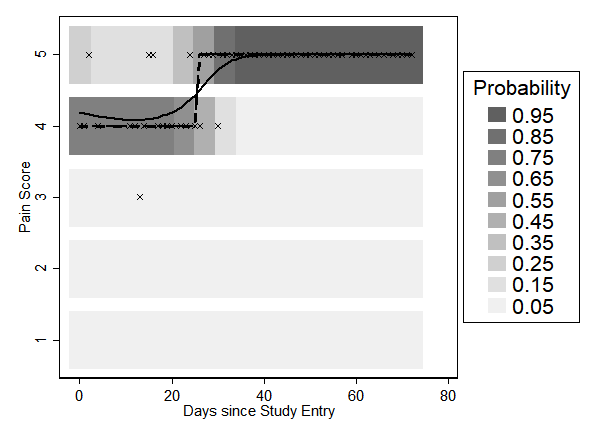

### nlin_plot_4.pdf

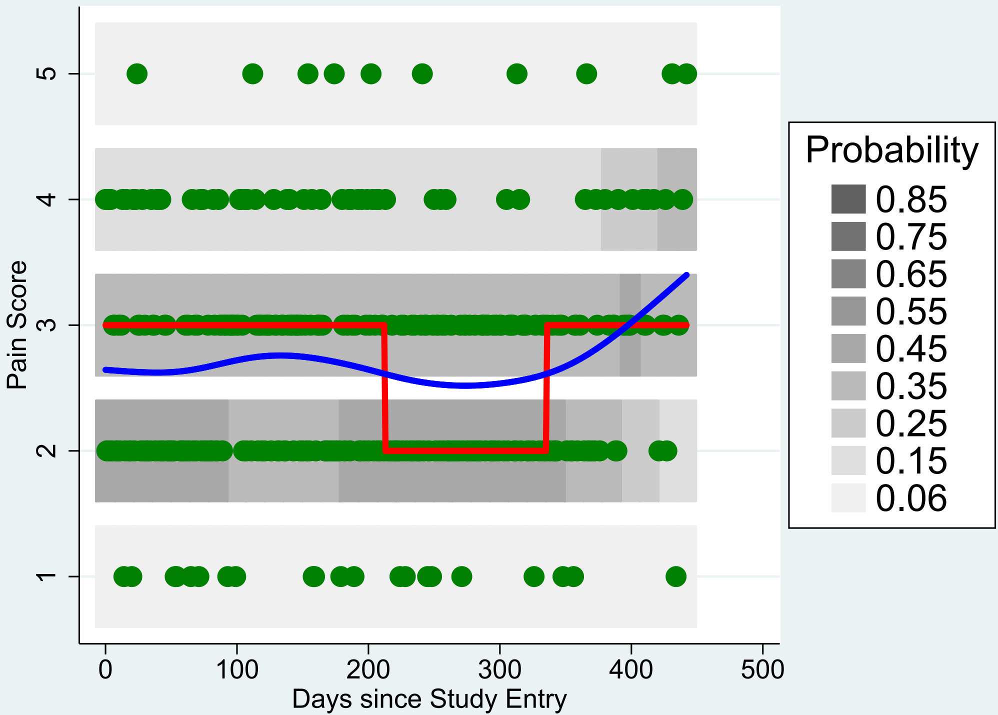

### nlin_plot_4.png

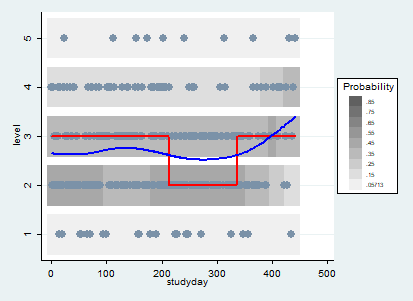

### nlin_plot_4_bw.png

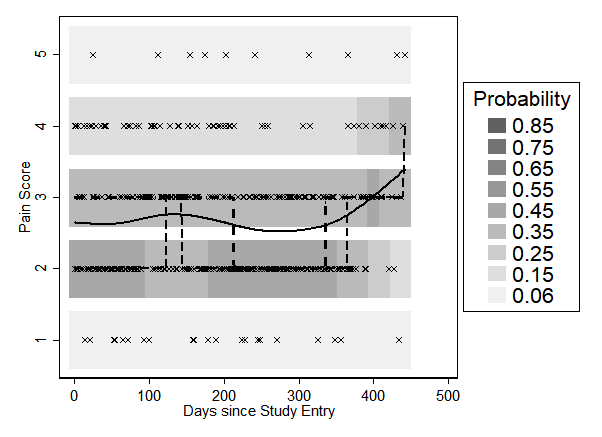

### nlin_plot_5.pdf

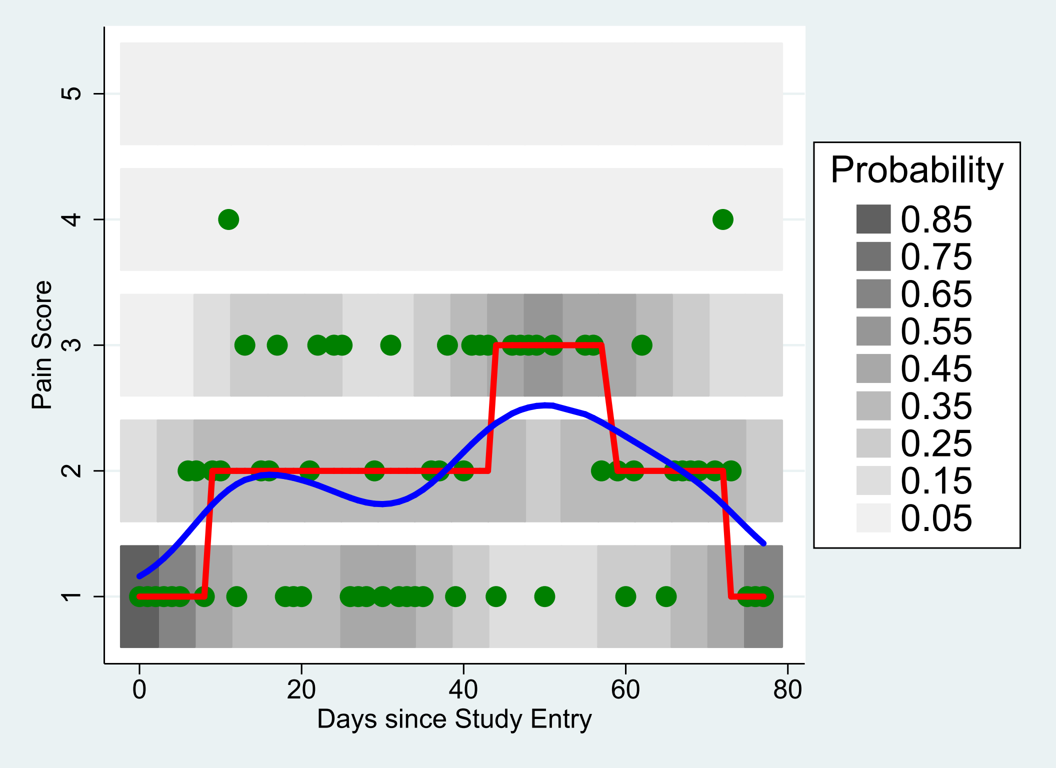

### nlin_plot_5.png

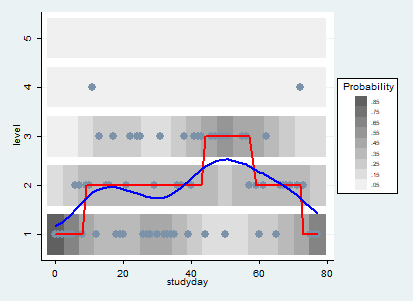

### nlin_plot_5_bw.png

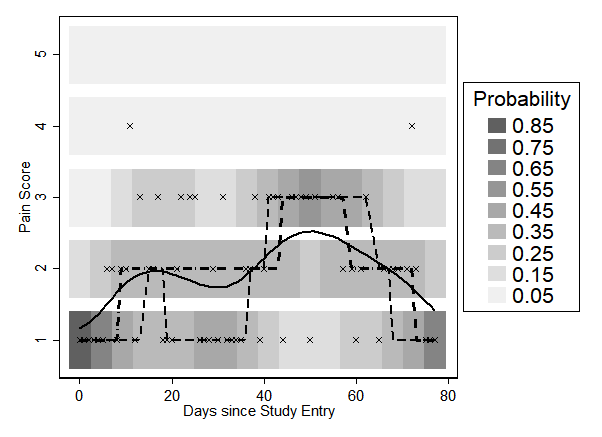

### nlin_plot_6.pdf

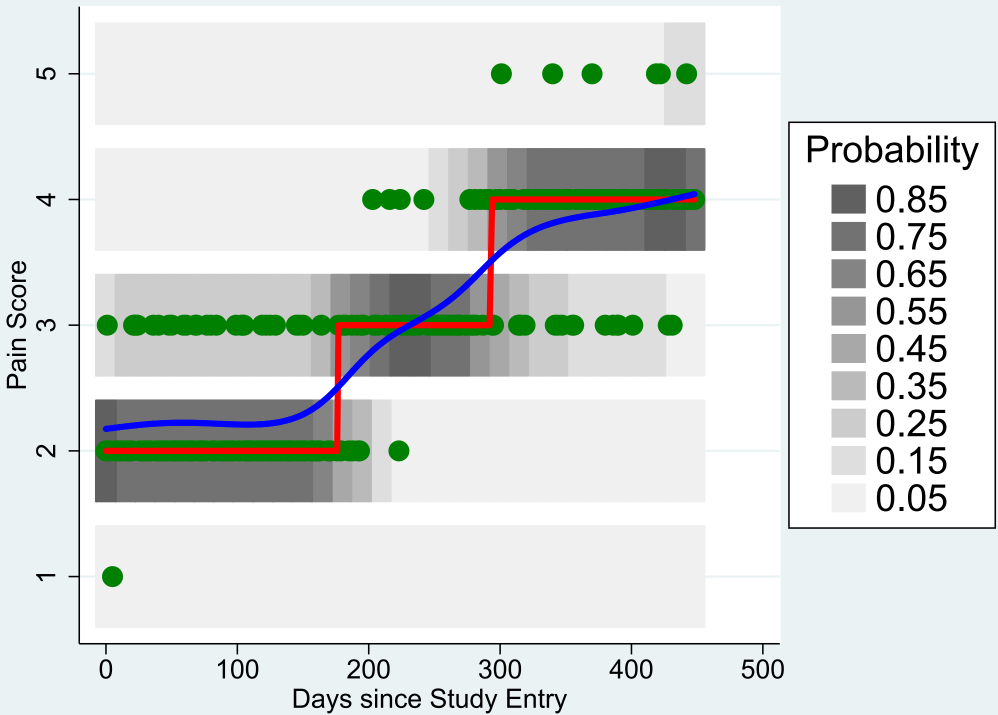

### nlin_plot_6.png

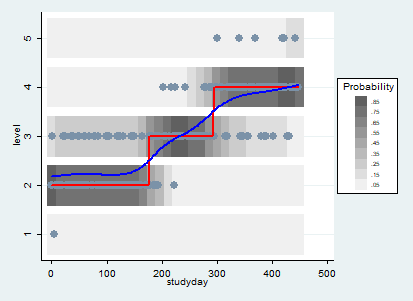

### nlin_plot_6_bw.png

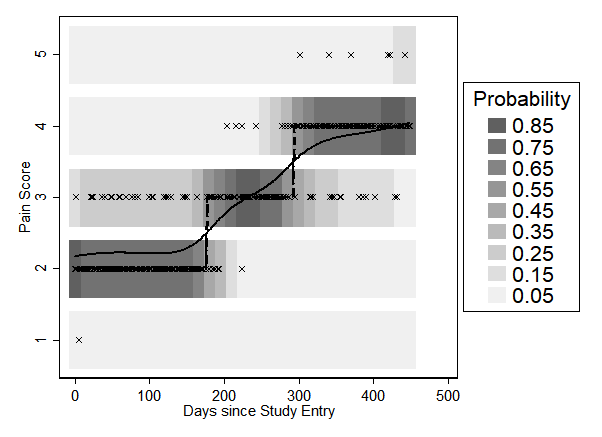

### nlin_plot_7.pdf

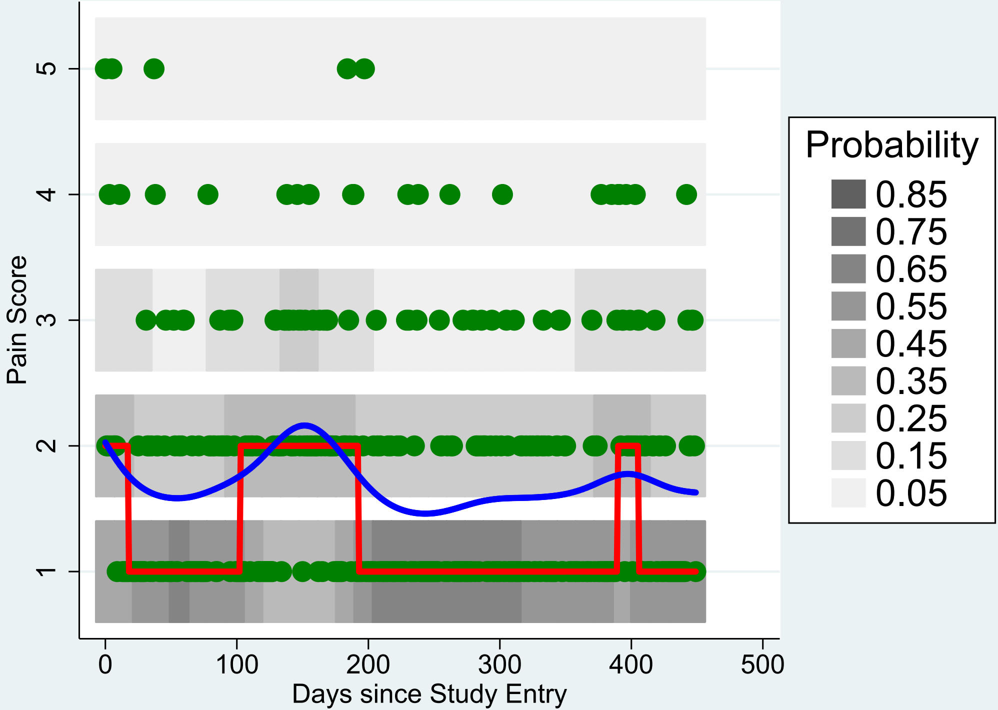

### nlin_plot_7.png

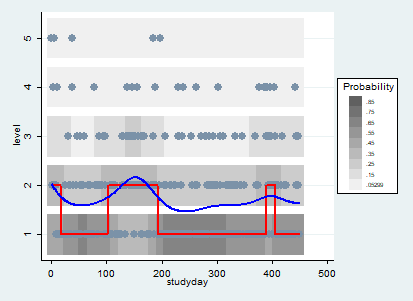

### nlin_plot_7_bw.png

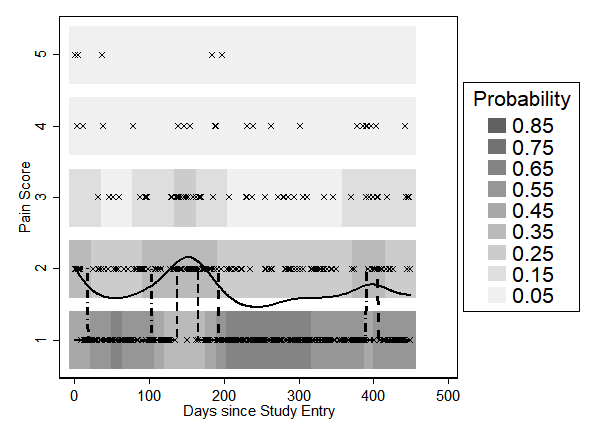

### nlin_plot_8.pdf

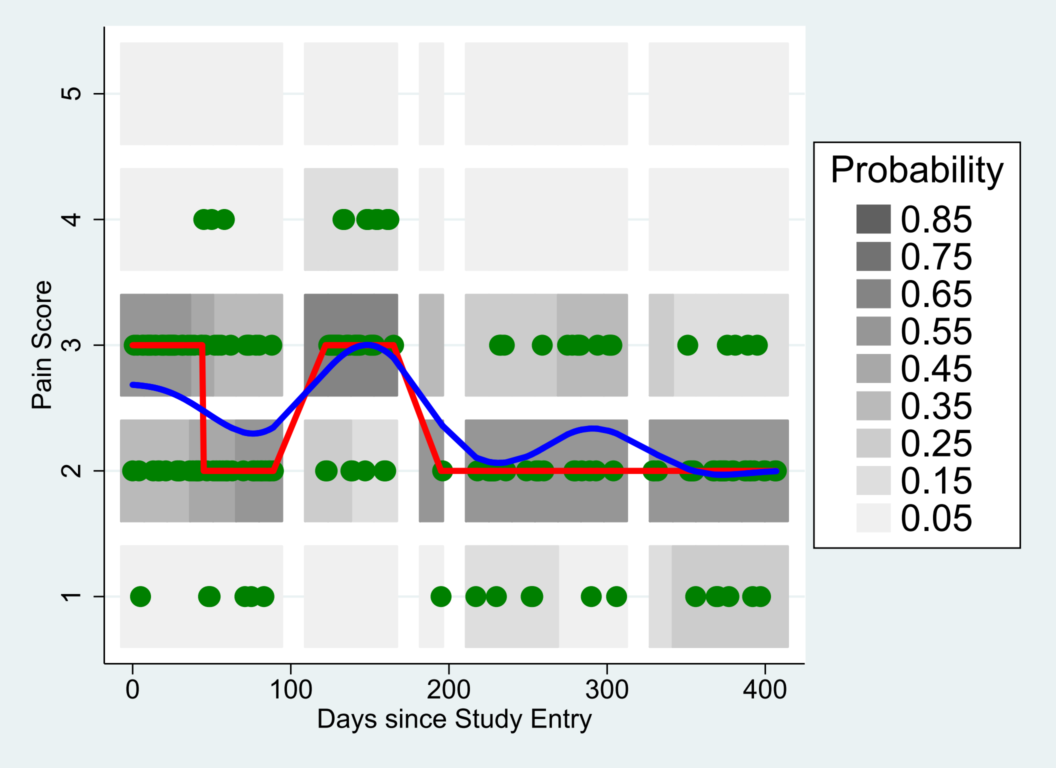

### nlin_plot_8.png

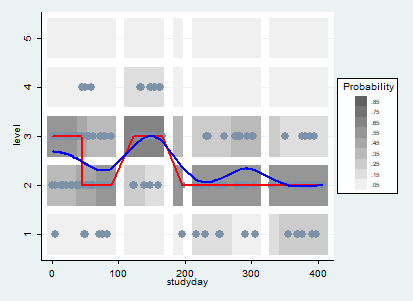

### nlin_plot_8_bw.png

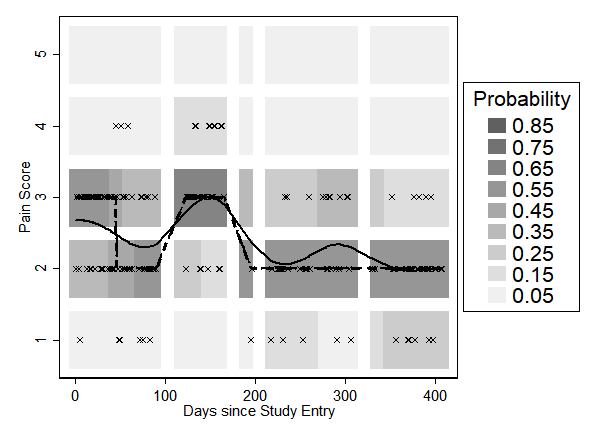

### nlin_plot_9.pdf

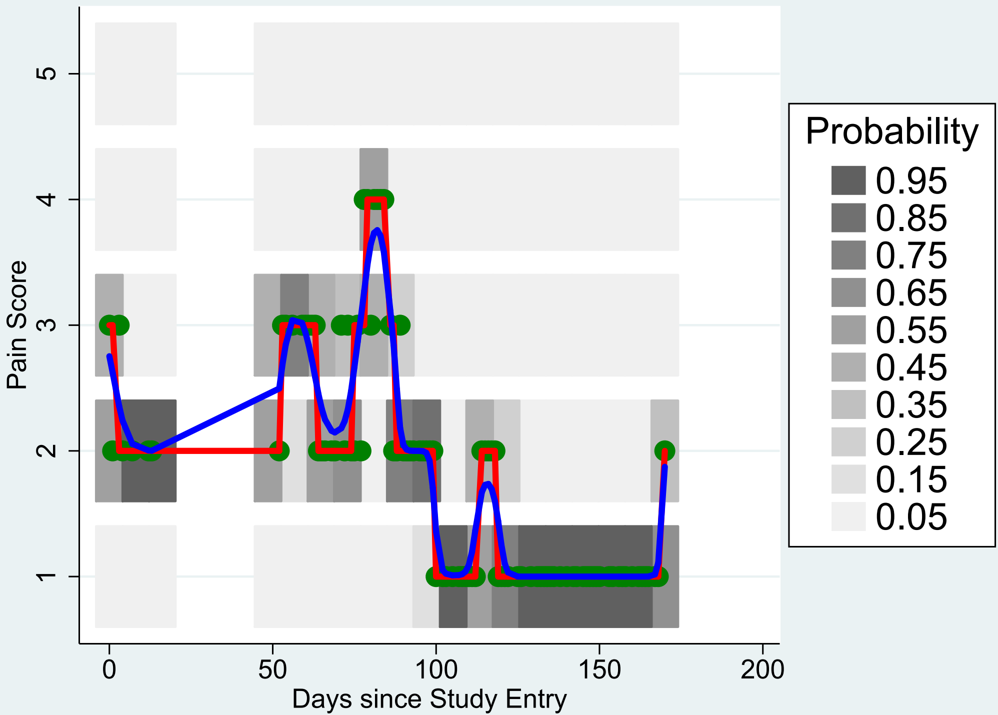

### nlin_plot_9.png

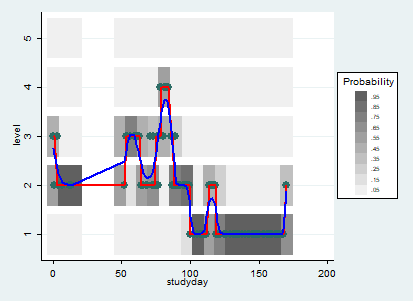

### nlin_plot_9_bw.png

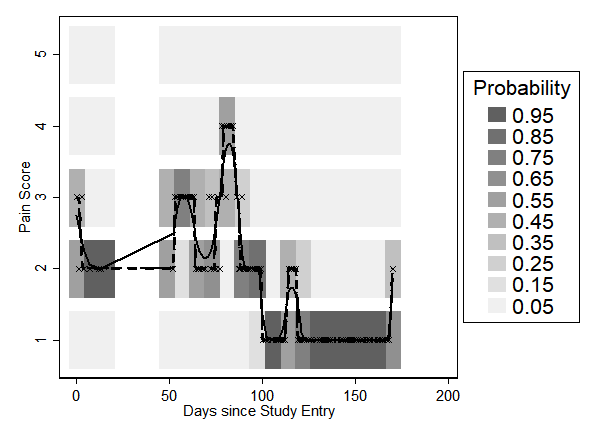

### nonsig_bars.png

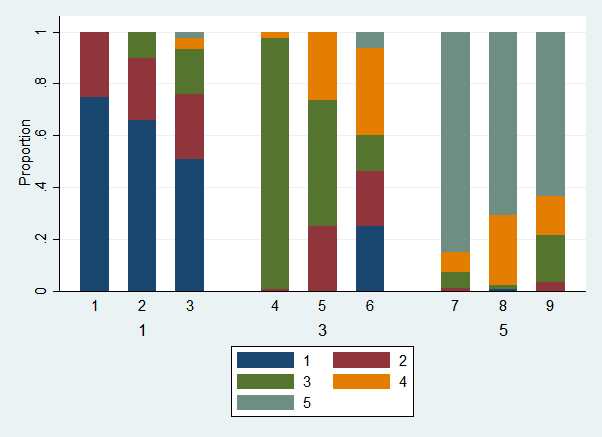

### nonsig_bars_bw.png

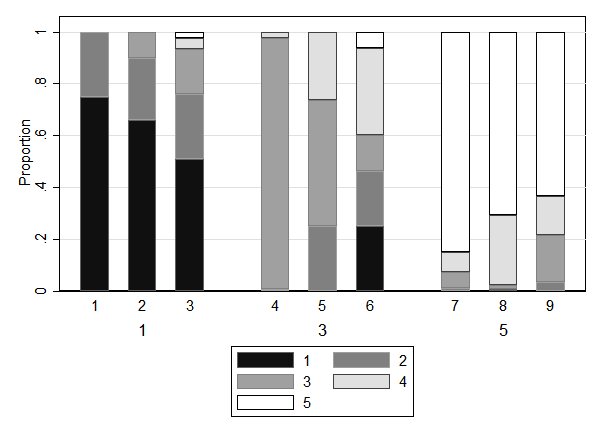

### obs_1.png

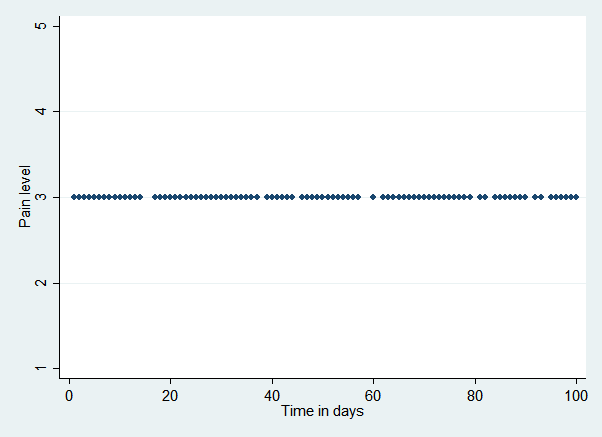

### obs_2.png

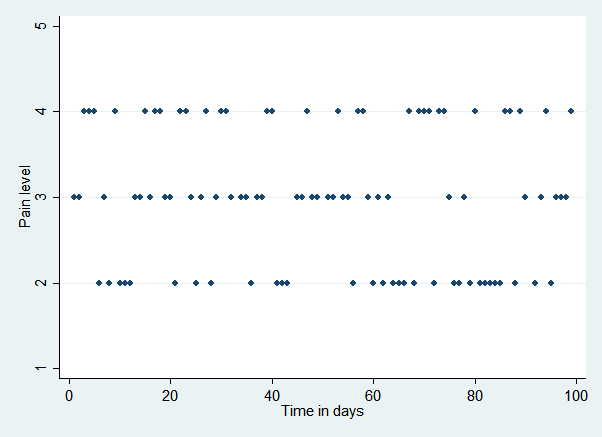

### obs_3.png

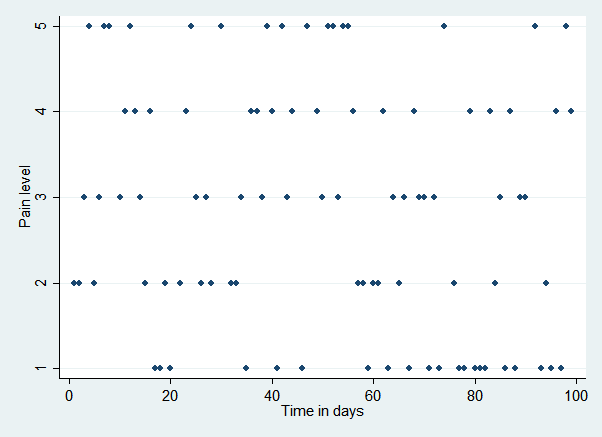

### obs_4.png

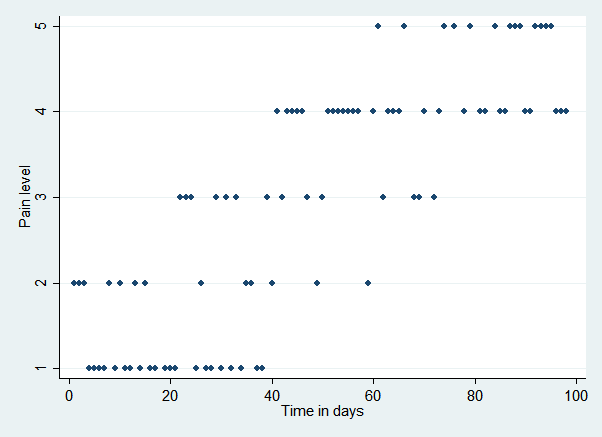

### obs_5.png

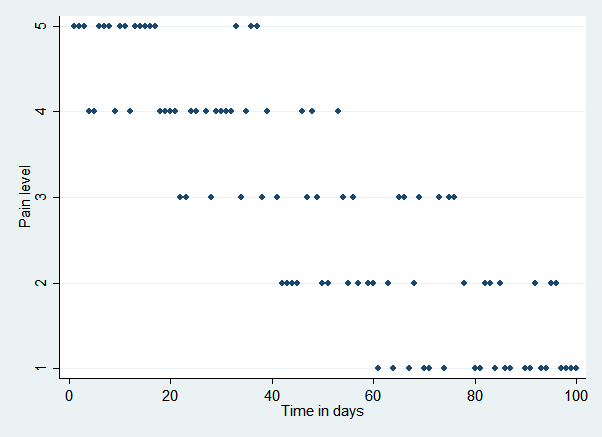

### obs_6.png

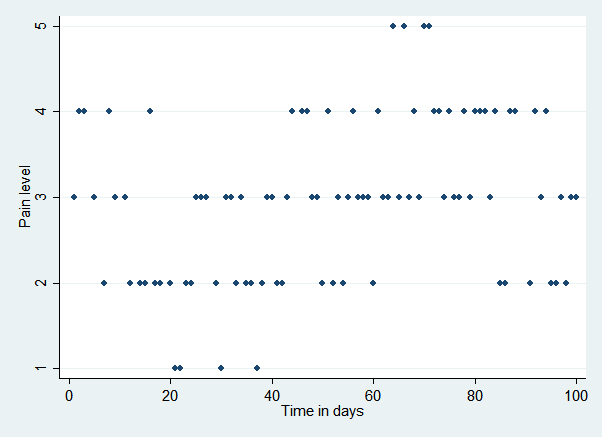

### obs_7.png

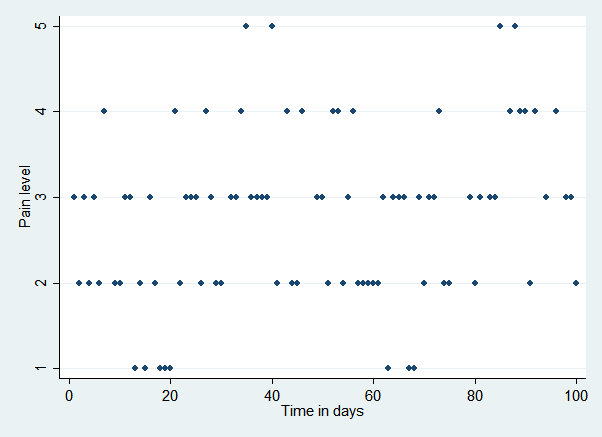

### obs_8.png

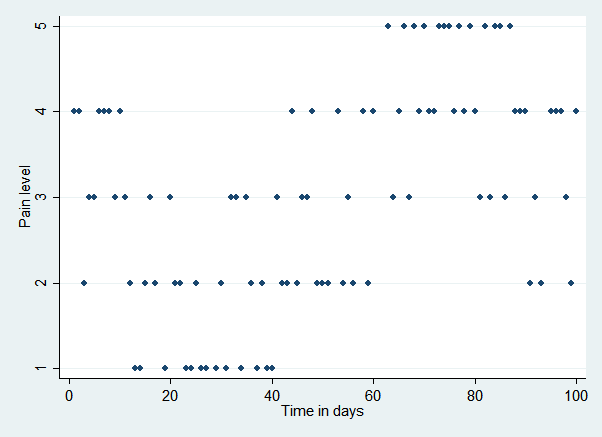

### sparsity_simulation_results.pdf

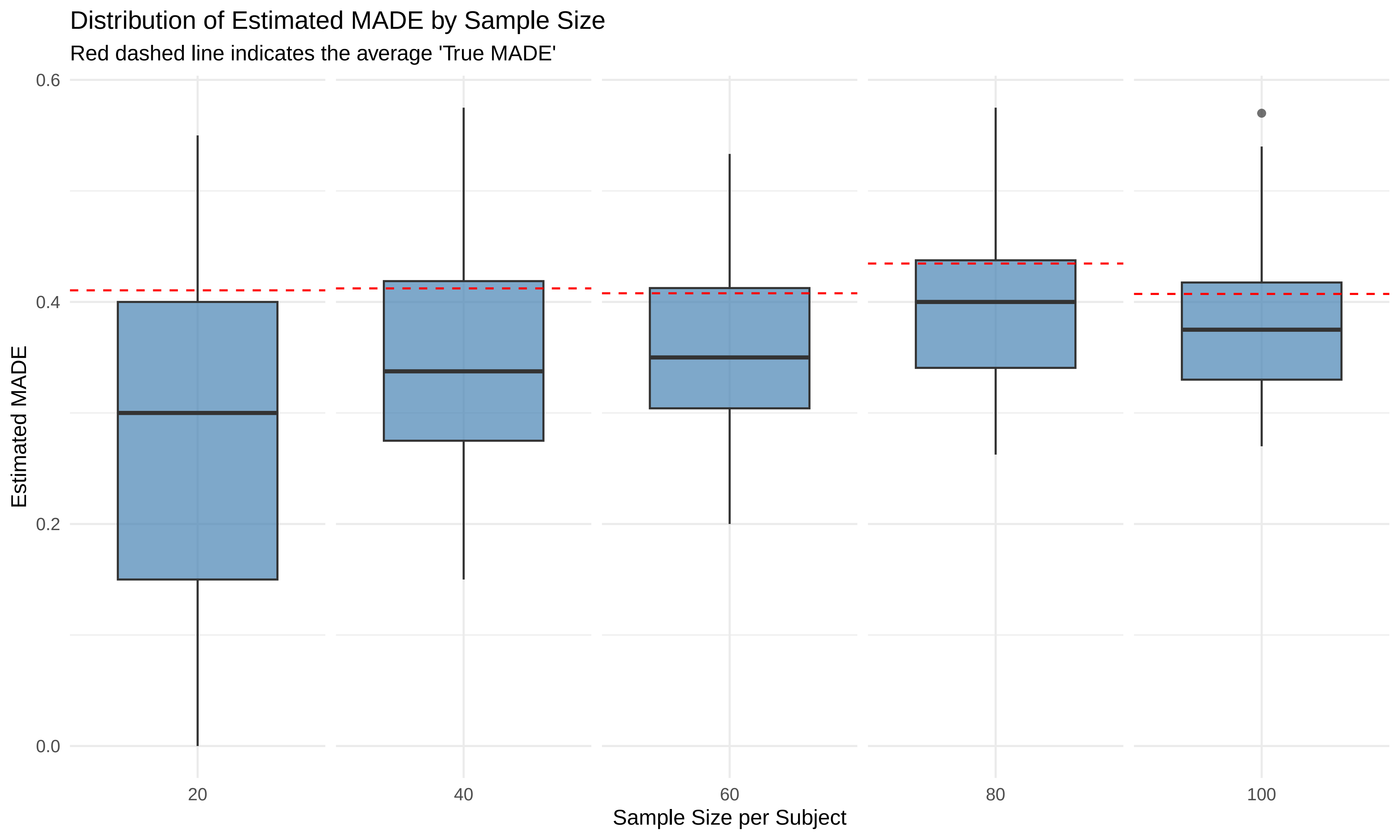

### sparsity_simulation_results_notitle.pdf

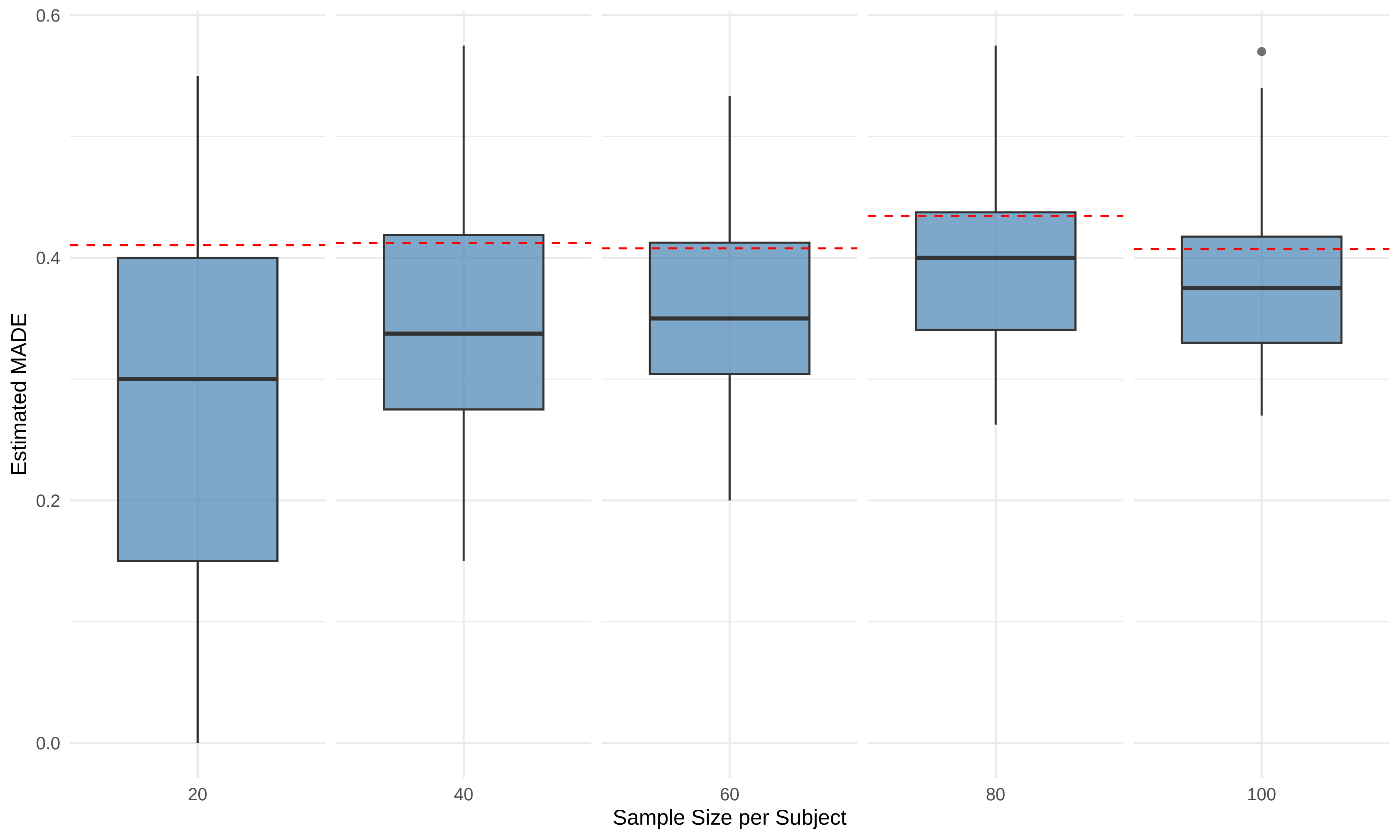

### stacked_bars.png

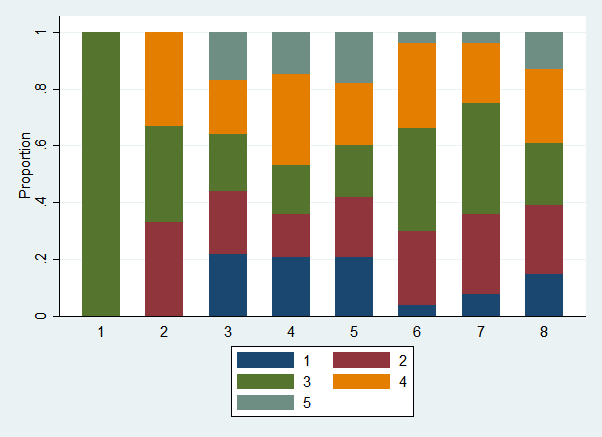

### stacked_bars_bw.png

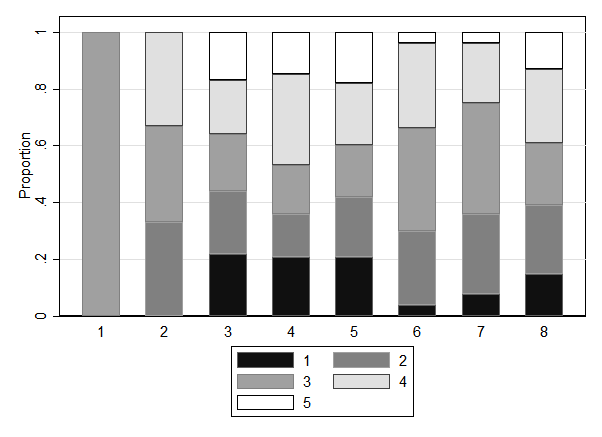
